# A scoping review of the format, content, effectiveness and acceptability of reproductive life planning tools

**DOI:** 10.1101/2024.05.31.24308272

**Authors:** Catherine Stewart, Hyun Noh, Neha Pathak, Jennifer Hall

## Abstract

**Purpose:** Screening for pregnancy intention and discussing Reproductive Life Planning (RLP) helps people reflect on their reproductive goals and how to achieve them. This scoping review maps the evidence on these approaches to inform their development and implementation in the UK.

**Methods:** We searched Medline, Embase, PsycINFO and Scopus, and Google for the grey literature. Data were extracted on audience, format, content, features, behaviour change theory, effectiveness and acceptability.

**Results:** 51 published and 21 grey literature sources were included. 25 RLPs were discussed in the published papers and 20 in the grey literature. Most came from the USA, most RLPs in published papers were asked verbally, while most grey literature RLPs were self-completed, there were six digital RLPs. Most ask whether an individual wants children, some then ask about number and timing of children, and contraception. RLPs are acceptable to people of reproductive age and healthcare professionals, and there is a desire for digital tools. While there is consistency in the questions asked, there is limited data on effectiveness and only three tools incorporated behaviour change theory.

**Conclusion:** A digital tool that builds on the evidence and integrates appropriate theory could enable more people to realise their reproductive goals.

## Introduction

“You’re either on contraception or you’re pregnant, and that middle step is missing”. So said one woman in our qualitative research(1), describing the gap that pregnancy intention screening (PIS) and reproductive life planning (RLP) is attempting to fill (see Box 1 for definitions used in this paper). Arguably the decision of whether or when to have children is one of the most important decisions a person will make in their lifetime, with far reaching impacts for health and wellbeing across the lifecourse. Despite this there is very little reliable and evidence-based support available for people to make these decisions, whether that is from health professionals, educational settings, digital sources, or from peers.

**Pregnancy intention screening** (PIS): a way of identifying a person’s desire for pregnancy, within a set timeframe, usually a single question. In a clinical context, this can be used to determine which reproductive health services to offer or could precede a reproductive life planning discussion.

**Reproductive life plan** (RLP): a set of longer-term personal goals about having or not having children and how to achieve those goals, based on the person’s own values, goals, and resources.

Box 1: Definitions of pregnancy intention screening and reproductive life plans

Reproductive life planning is a concept developed in the USA that has been championed by the Centers for Disease Control and Prevention (CDC)(2). It is a set of questions that health care providers were encouraged to use to prompt their patients to reflect on their reproductive goals and the actions that they need to take to achieve them (Box 2). This could lead to advice on how to improve health before pregnancy (preconception care) to increase the chances of pregnancy(3) and reduce adverse birth outcomes(4, 5), counselling on contraceptive choices, or support for those who are unsure or ambivalent about a future pregnancy, recognising that pregnancy planning is not a salient concept for some(6).

• Do you plan to have any(more) children?

• How many children do you hope to have?

• How long do you want to wait until you become pregnant (again)?

• How much space do you plan to have between your future pregnancies?

• What do you plan to do to avoid pregnancy (until you are ready to become pregnant)?

• What can I do to help you achieve your plan?

Box 2: Questions in the RLP developed by Moos (2006) and adopted and adapted by the CDC(7).

As the implementation of reproductive life planning has been developed it has increasingly been emphasised that it should be used in a way that supports reproductive justice and patient autonomy, is not judgemental, and is patient-centred i.e. focusing on the patient’s priorities and not what the healthcare professional thinks those should be, particularly when it comes to contraceptive choice(6). Consequently, five key attributes of reproductive life planning have been defined: goal-orientated; personalised; collaborative but person-centred; fluid; and health promotion-focused(8). Pregnancy intention screening is an antecedent to reproductive life planning(8), often a single question in a clinical encounter which seeks to establish a person’s current attitude towards pregnancy, and which may begin the process of reproductive life planning, resulting in the formulation of a reproductive life plan. Throughout this paper we use RLP to refer to both the *process* and the *output* of reproductive life planning.

To date there has been no attempt to implement pregnancy intention screening or reproductive life planning in the United Kingdom. However, in 2023 it was recommended as the entry point for holistic care across the reproductive life course in primary care in England(1). There are a range of potential tools available globally for asking about pregnancy preferences and it is not clear which would be best in any given setting. Furthermore, women and healthcare professionals have expressed preferences for a digital approach, or one that integrates digital and in-person modalities(1, 9, 10), and the evidence for this needs to be explored.

This scoping review of the published literature, and review of RLPs available in the grey literature, aims to map out what is currently available and describe the content and format of existing RLPs, as well as synthesise any information on their effectiveness or acceptability, with a particular focus on digital tools, to inform their development and implementation in the UK.

## Methods

We conducted a scoping review of the published and grey literature(11). The search protocol was pre-registered with OSF (DOI 10.17605/OSF.IO/S26CV). We searched four databases: Medline; Embase; PsycINFO; and Scopus, accessed via the Ovid and Scopus interfaces, and used Google to search the grey literature. After extensive testing we created a parsimonious search strategy of keywords, as there were no suitably focused MeSH terms, of “reproductive life plan$” OR “reproductive plans$” OR “pregnancy intention screening”. The search was limited to peer-reviewed studies, in humans, published from 1 January 2015, when the first digital RLPs began to appear, to 24th October 2023. Observational and intervention studies were included. The full electronic search strategies are included in S1. We searched Google on 15/11/2023 using the search strategy “reproductive life plan$” OR “reproductive plan$ tool$”, extracting the first 50 results. We re-ran all searches on 28 January 2026. Additional inclusion criteria were that the study focused on people of reproductive age, or health professionals serving them, and was about any reproductive life planning or pregnancy intention screening tool. Studies were excluded if they were not in humans, not available in English or not a primary study. The same criteria were applied to papers and grey literature.

Database search results were extracted and uploaded to Rayyan where duplicates were removed, and title/abstract screening was managed. The list of papers selected for full text review was uploaded to Endnote. To minimise bias and increase objectivity, reviewers worked in parallel for all screening rounds, blind to each other’s decisions. Any discrepancies were resolved by a third reviewer. Reviews were excluded after references were checked for additional studies meeting the inclusion criteria. References and citations of included studies were screened to identify additional studies. Data items extracted into an Excel spreadsheet designed for this review included meta data about the studies, target audience, format, content, including any associated behaviour change theory, features, effectiveness and acceptability of the PIS/RLP. Findings were synthesised within each of these categories with differences between tools, settings and formats considered.

We contacted authors of any papers that did not include the wording of the PIS or RLP content in the paper. Of the three authors contacted (12–14) only one replied (14).

## Results

The initial database searches in 2023 yielded a total of 432 publications: 135 were duplicates; 235 were removed on title and abstract review; and 62 full-text papers were retrieved. Thirty-one of these were excluded, mostly because they were not about PIS/RLP. Thirteen additional publications were identified from forward and backward citation searching. The updated database searches undertaken in 2026 yielded 559 records. Following de-duplication, 131 new publications remained for screening. Title and abstract review resulted in the exclusion of 104 records, with 27 articles progressing to full-text assessment. Twenty were excluded for not being related to PIS/RLP.

From the grey literature search, 18 results were retained in 2023, with an additional three results in 2026; most results were excluded as they were a research paper that had been identified in the database search.

The PRISMA flowchart is shown in Figure 1.sss

**Figure 1:**
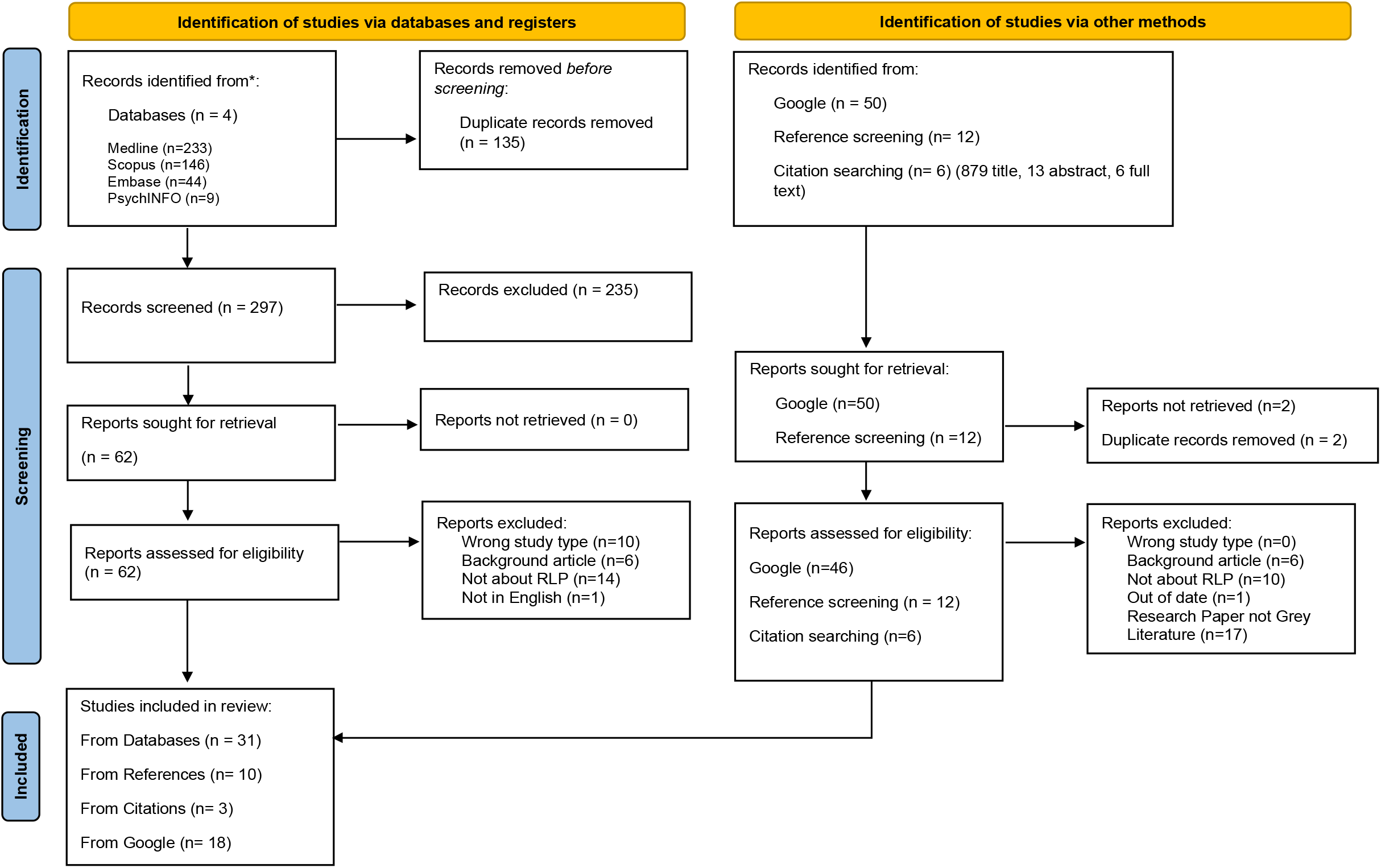
Flowchart of study identification

### Included studies

Ultimately, 51 papers were included in our review and are described in Table 1(6, 9, 12–60). Although published between 2015-2026, data were collected between 2012-2023. The majority of papers came from the USA (n=37). Qualitative and cross-sectional studies were the most common (n=10), followed by pilot studies (n=9) and randomised controlled trials (n=8). Most studies took place in clinical settings: either primary/community health services (n=24) or secondary clinic/specialist clinic/hospital settings (n=19). Within the grey literature sources most were published by health care services in the USA and are described in Table 2 (61–81). All grey literature links were working at time of extraction, however six of the 18 grey literature links from 2023 were broken at time of second extraction in 2026 (see Table 2 for details).

**Table 1:**
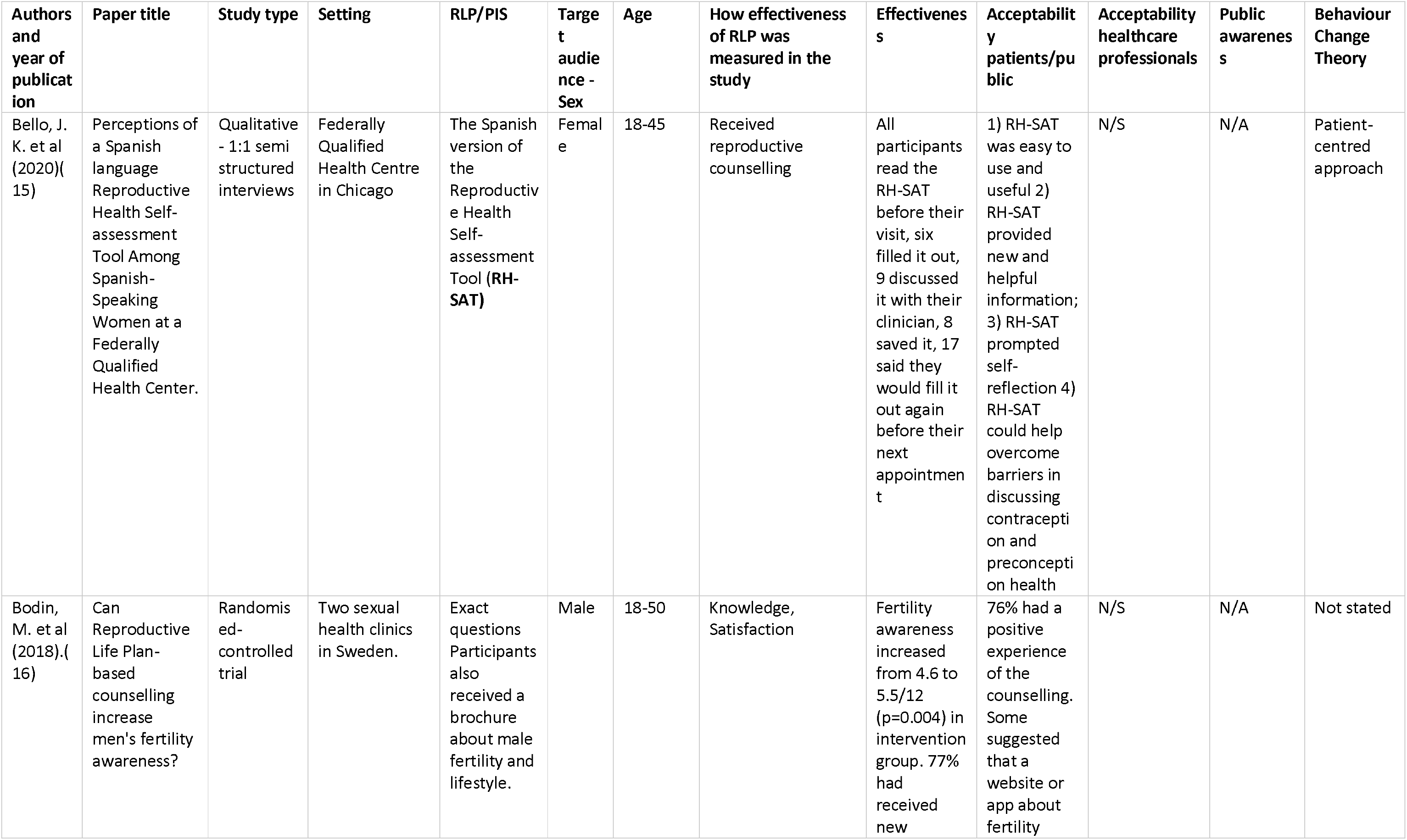
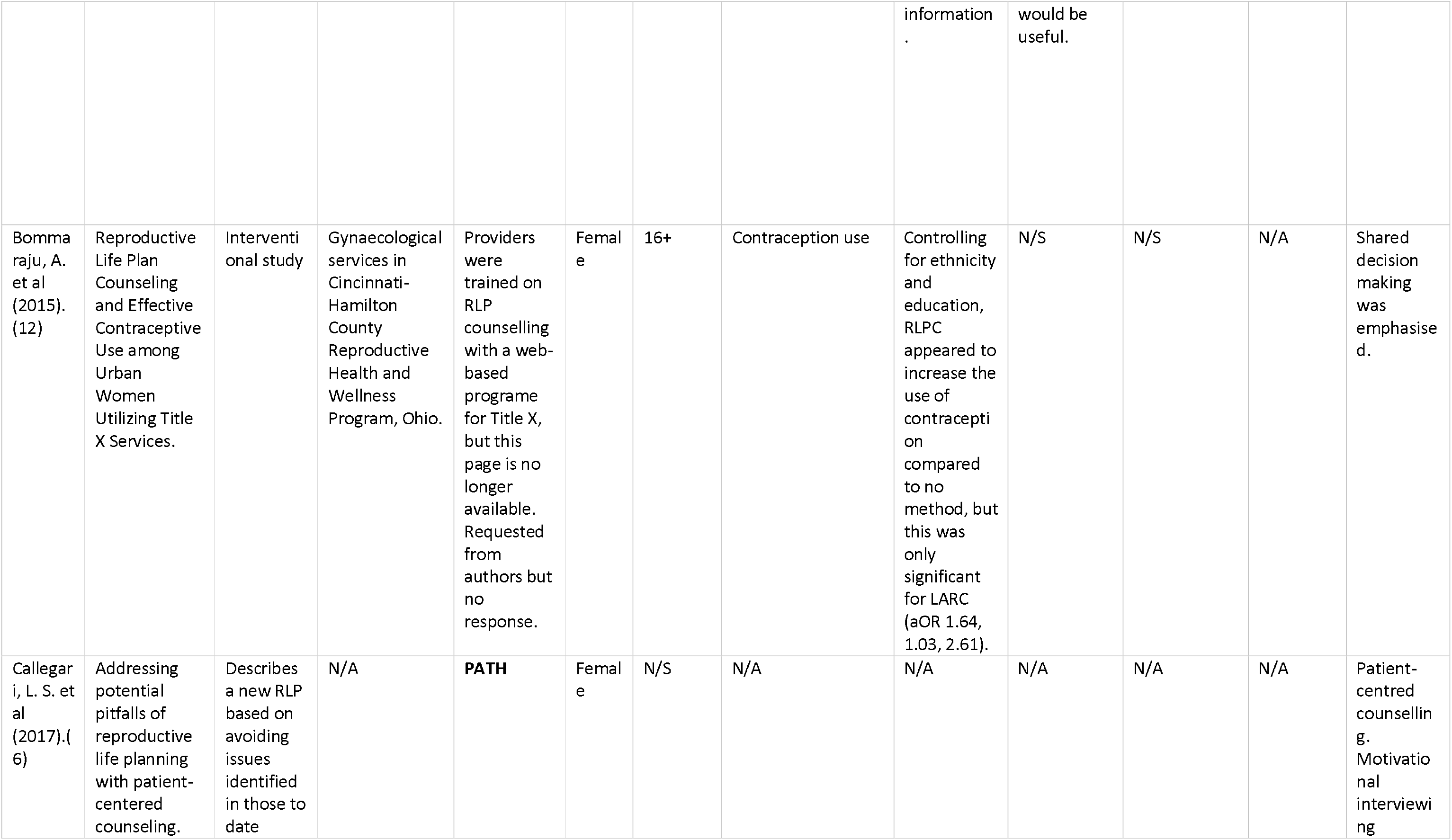
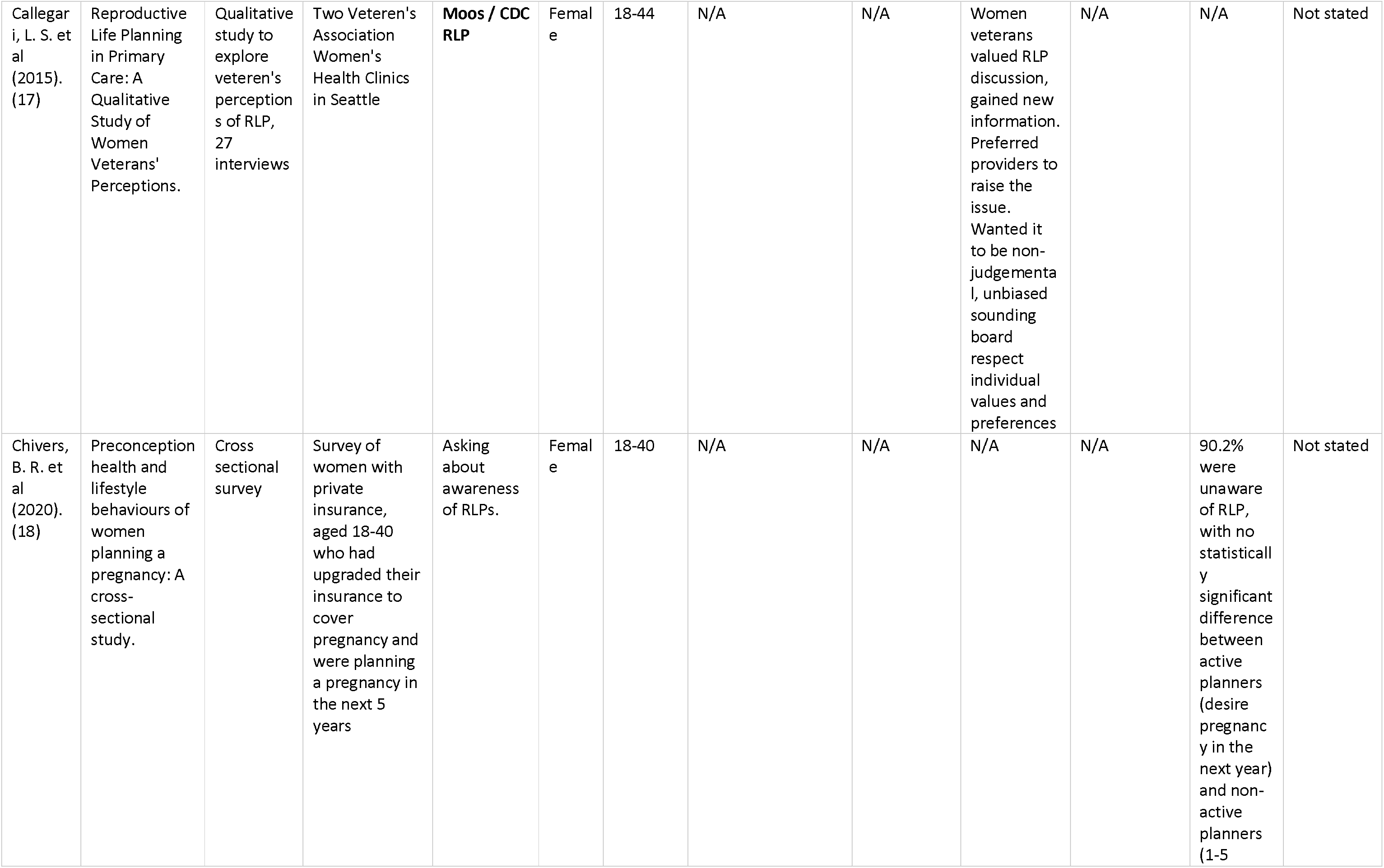
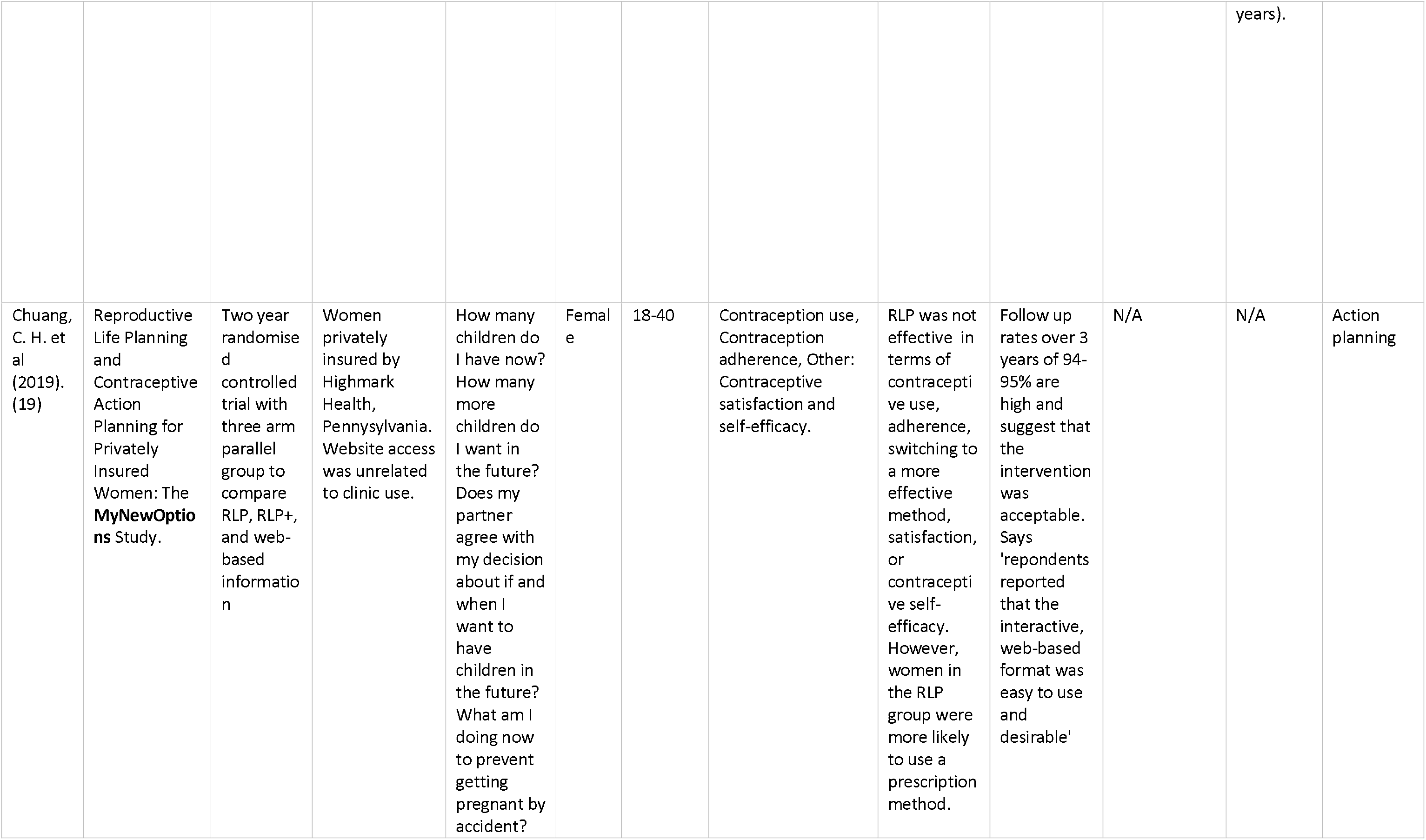
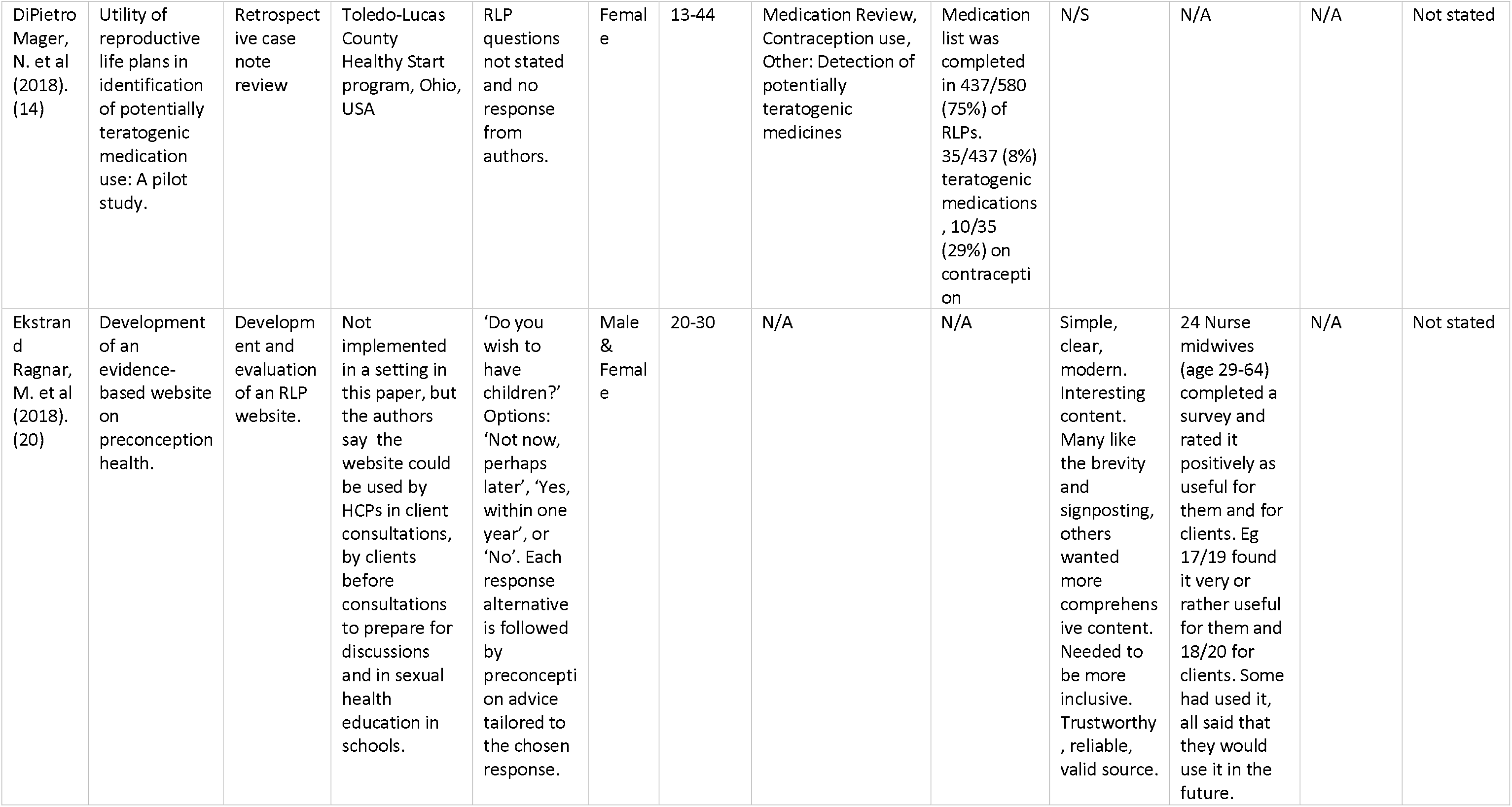
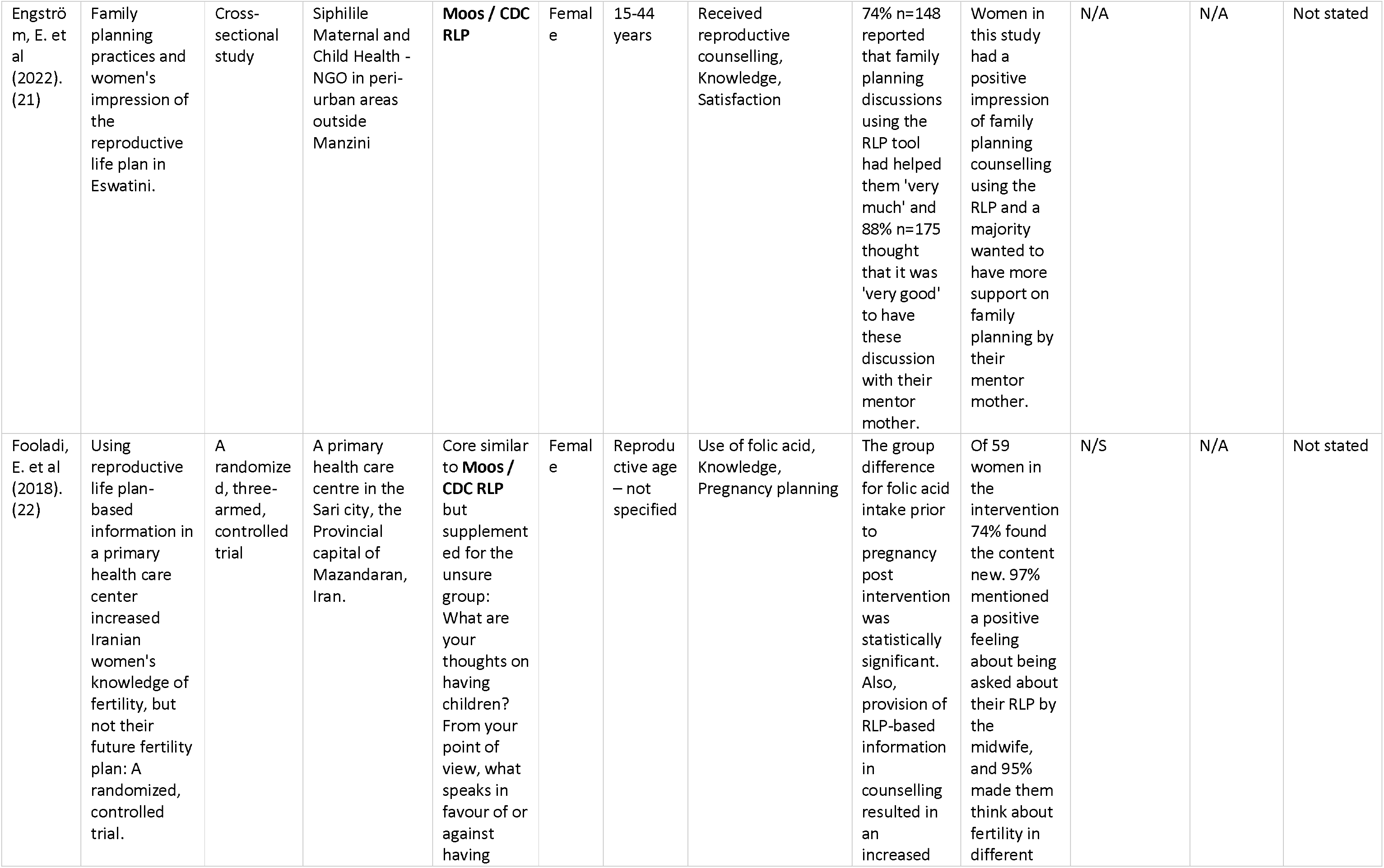
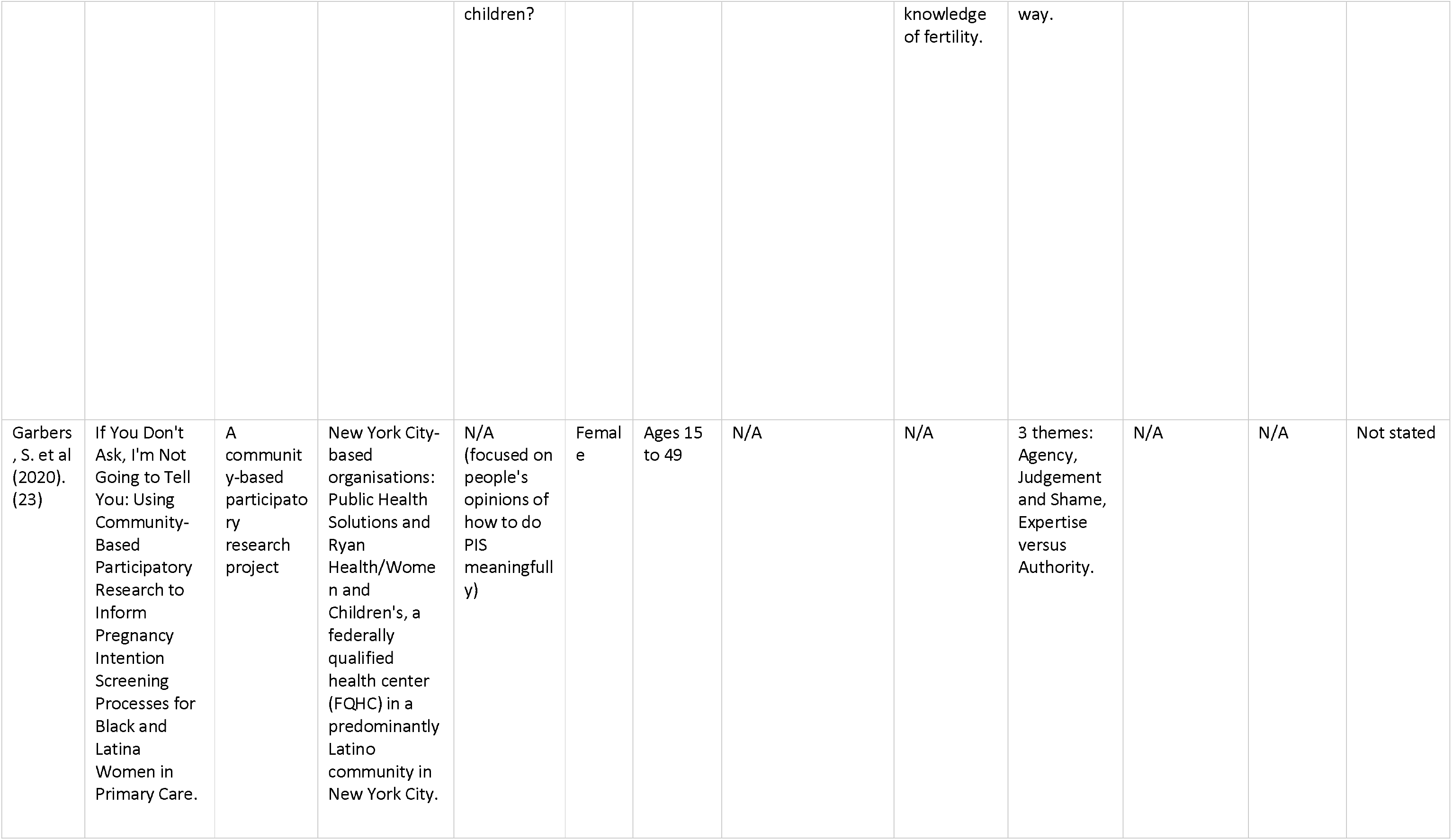
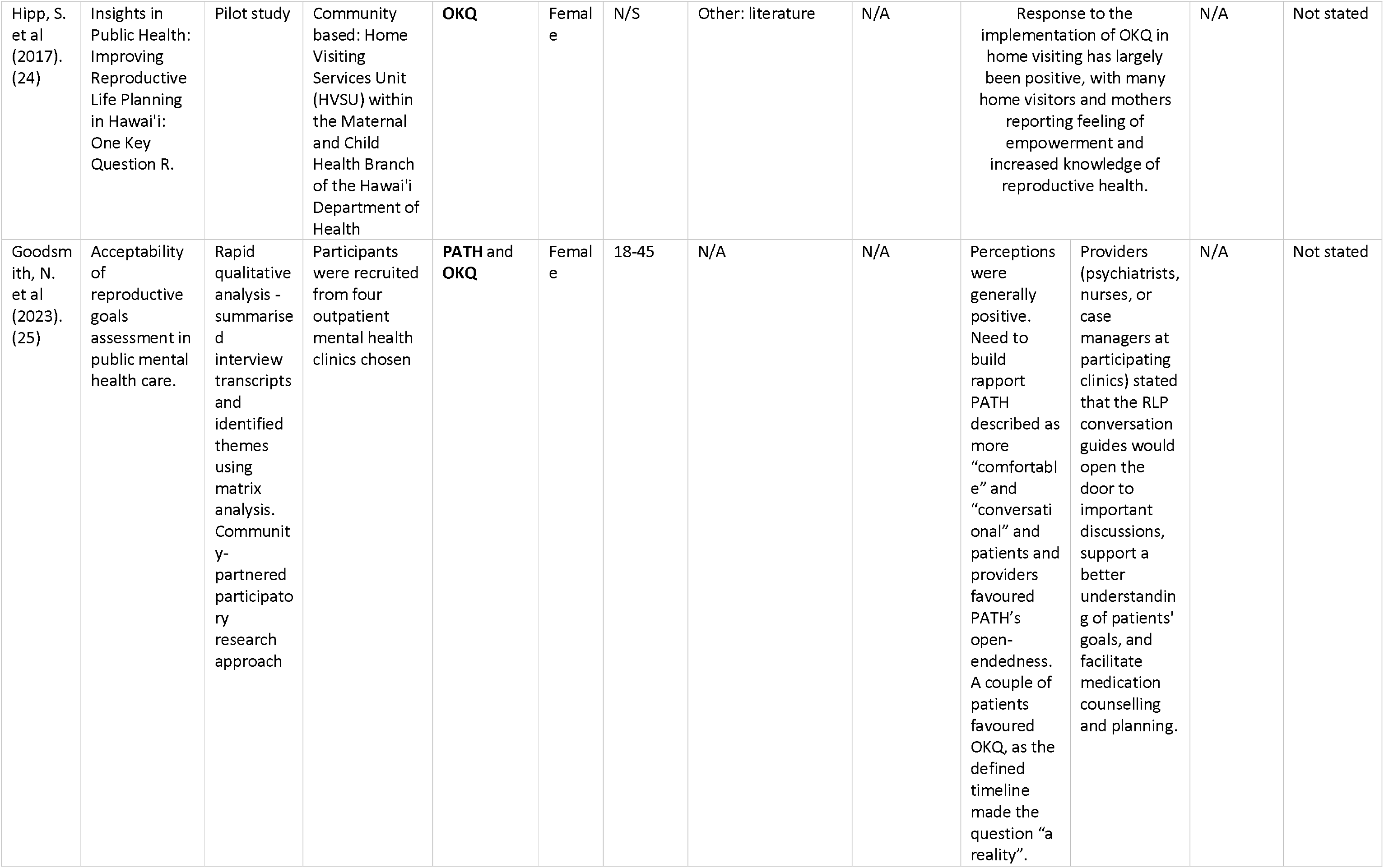
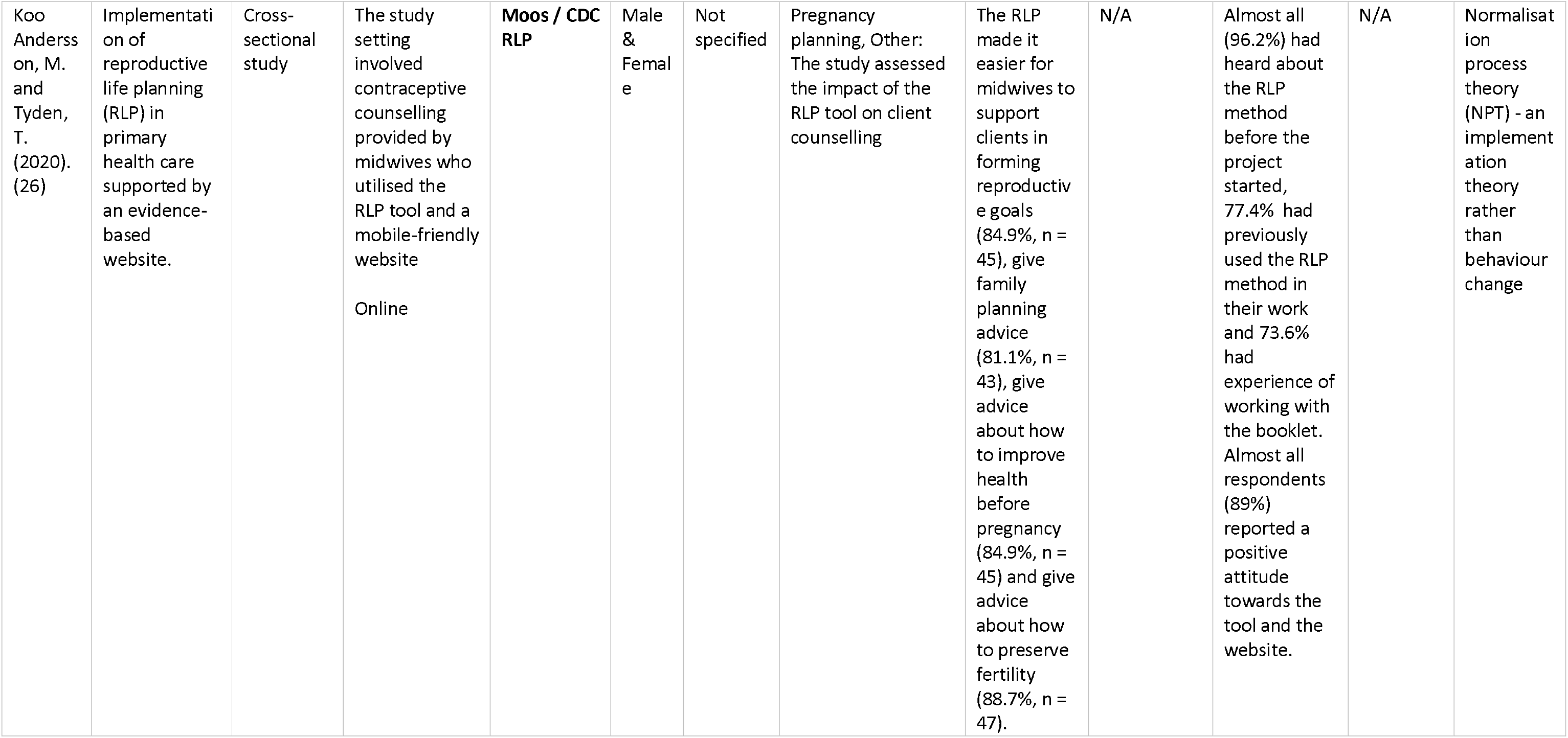
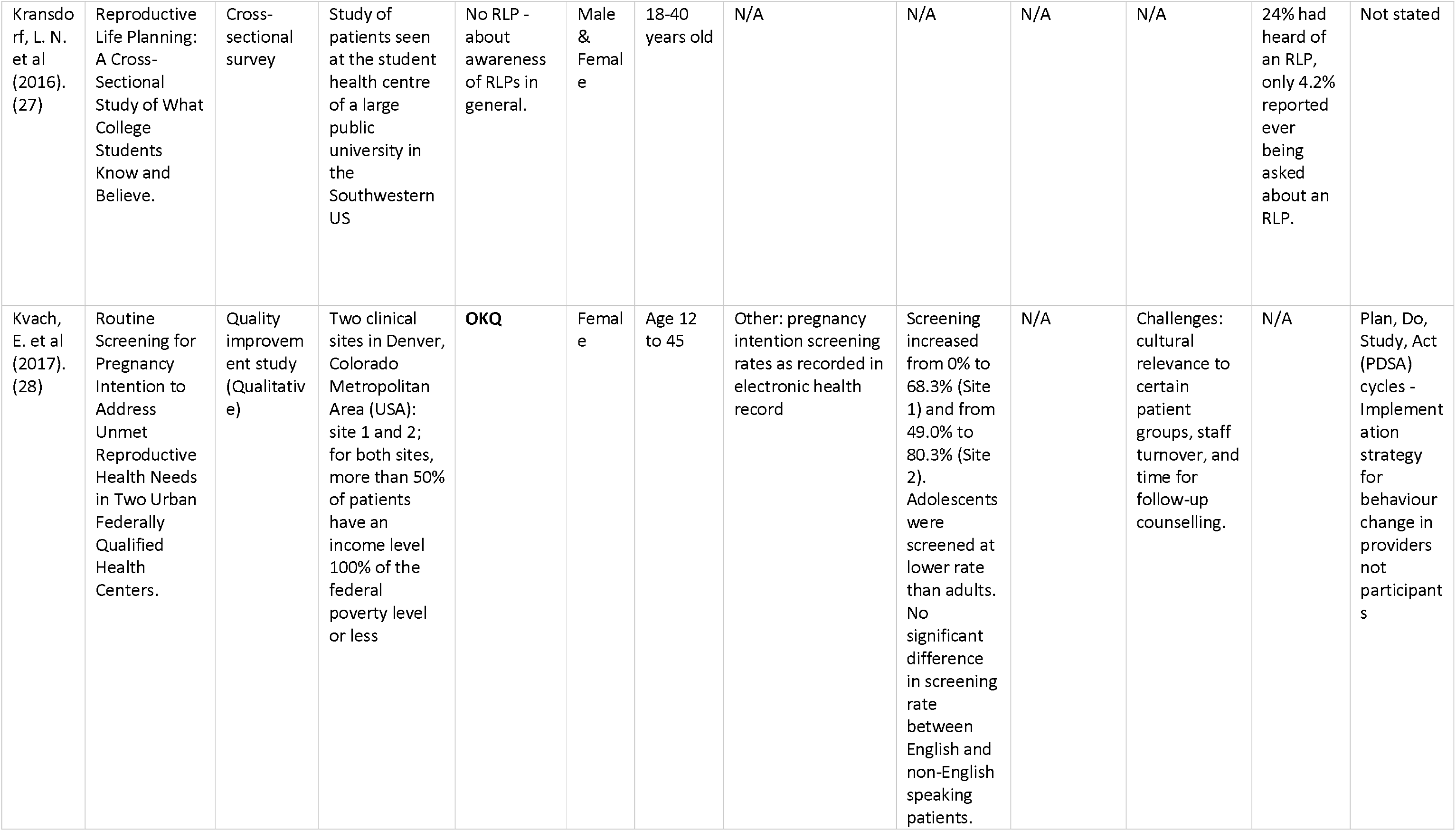
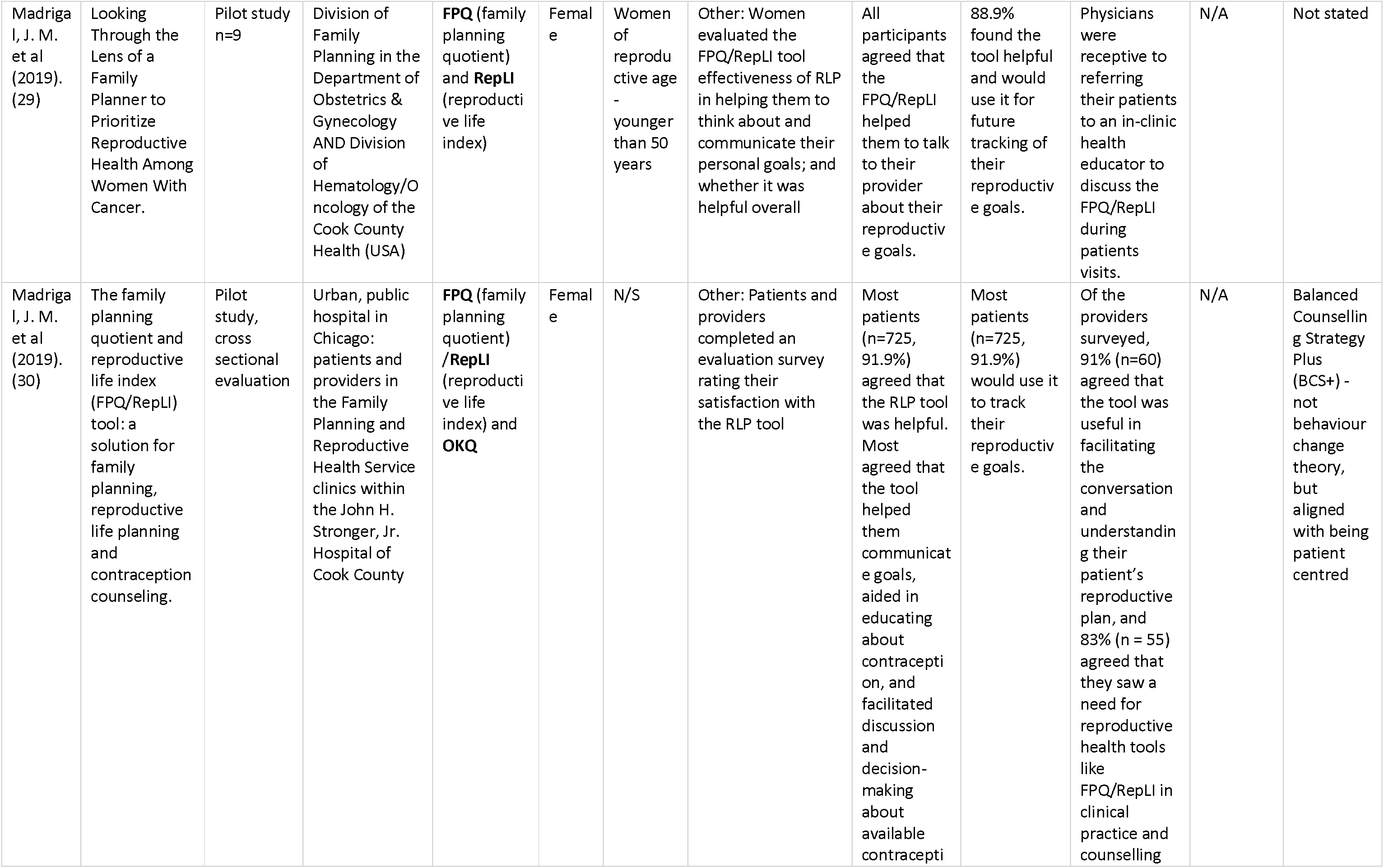
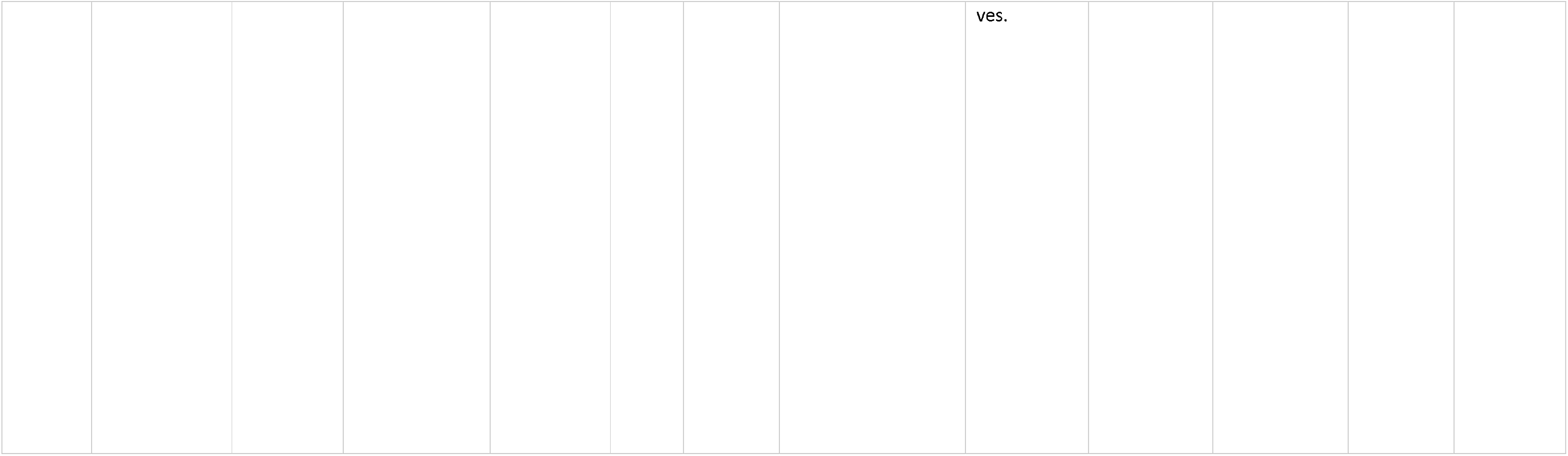
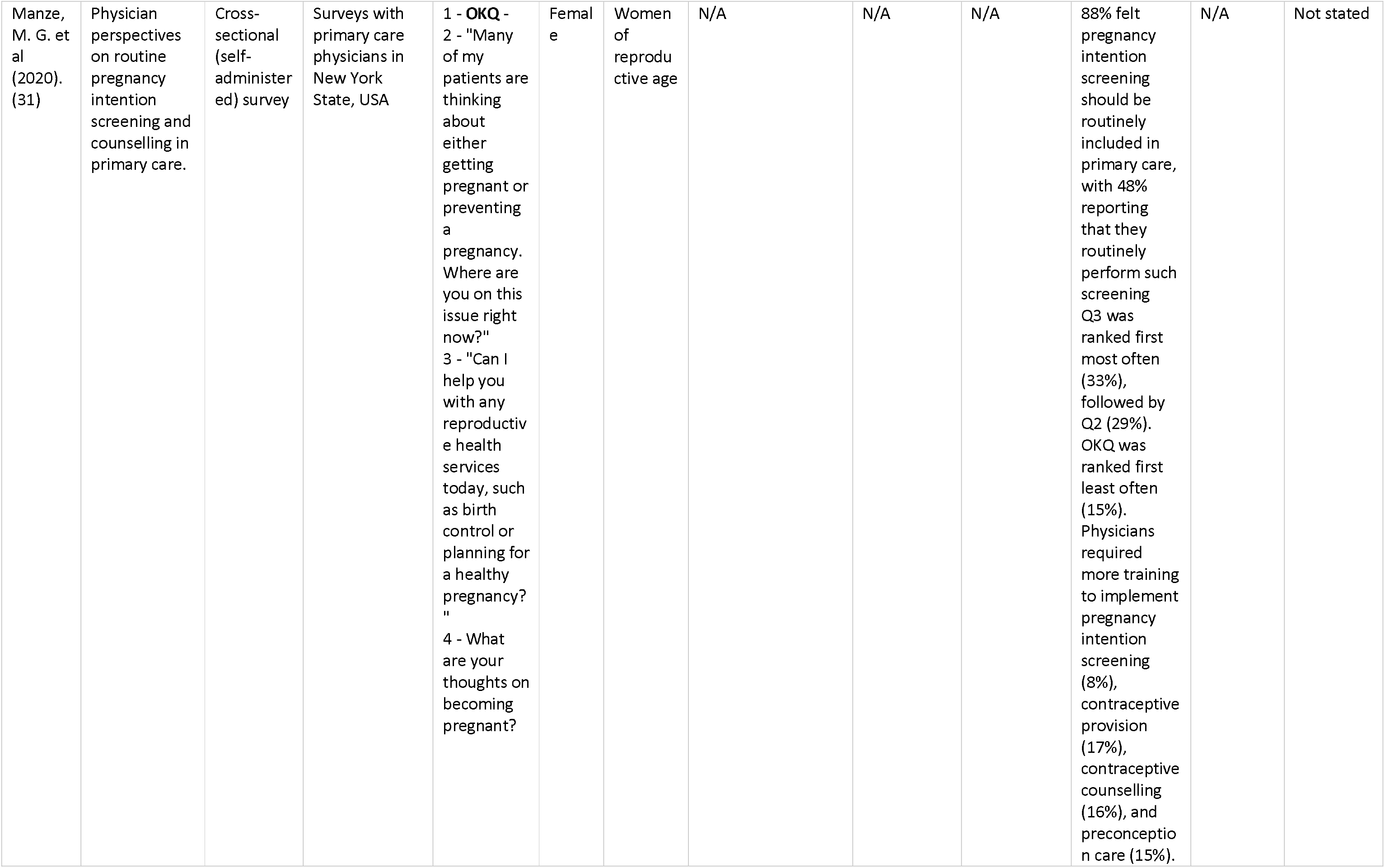
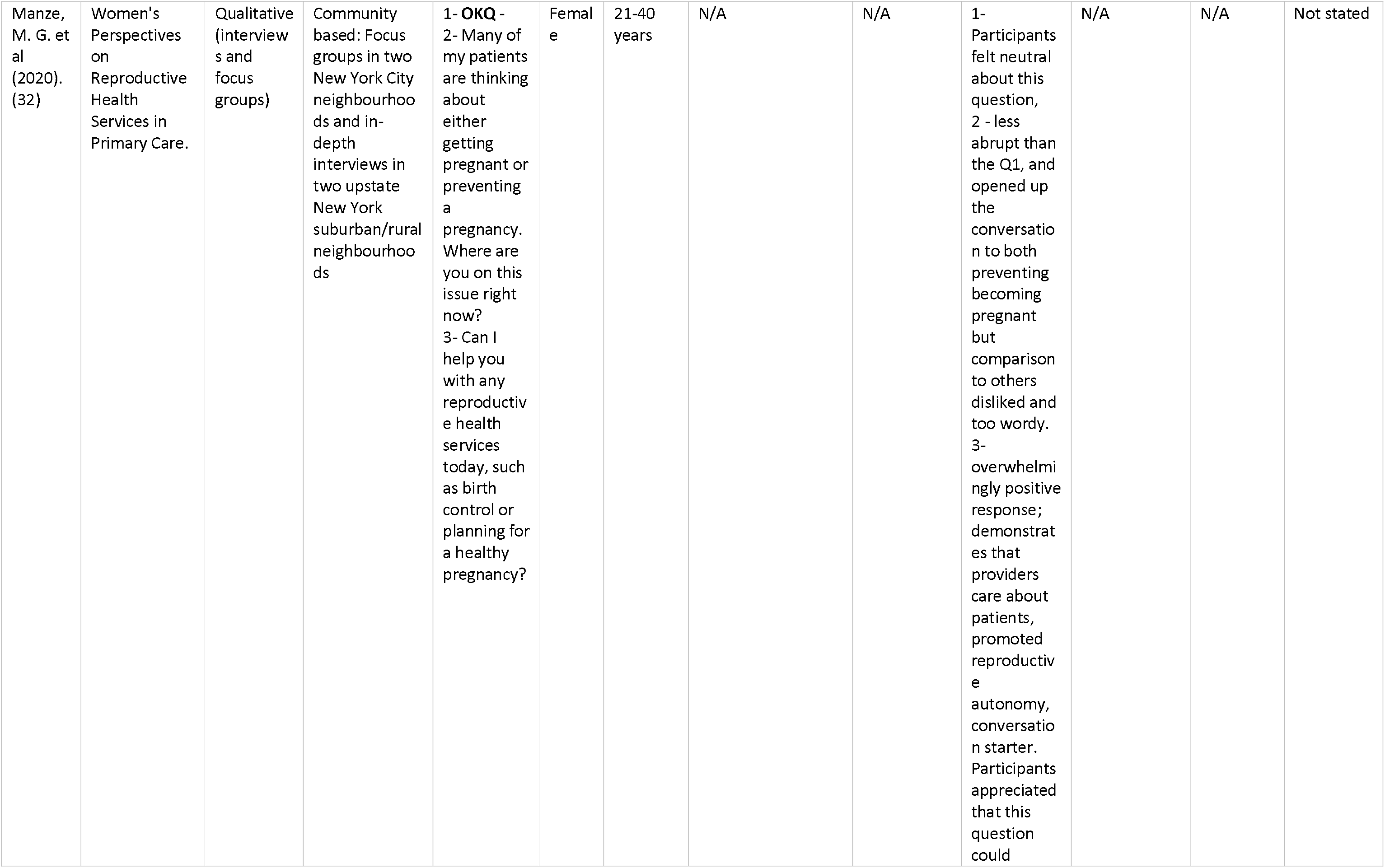
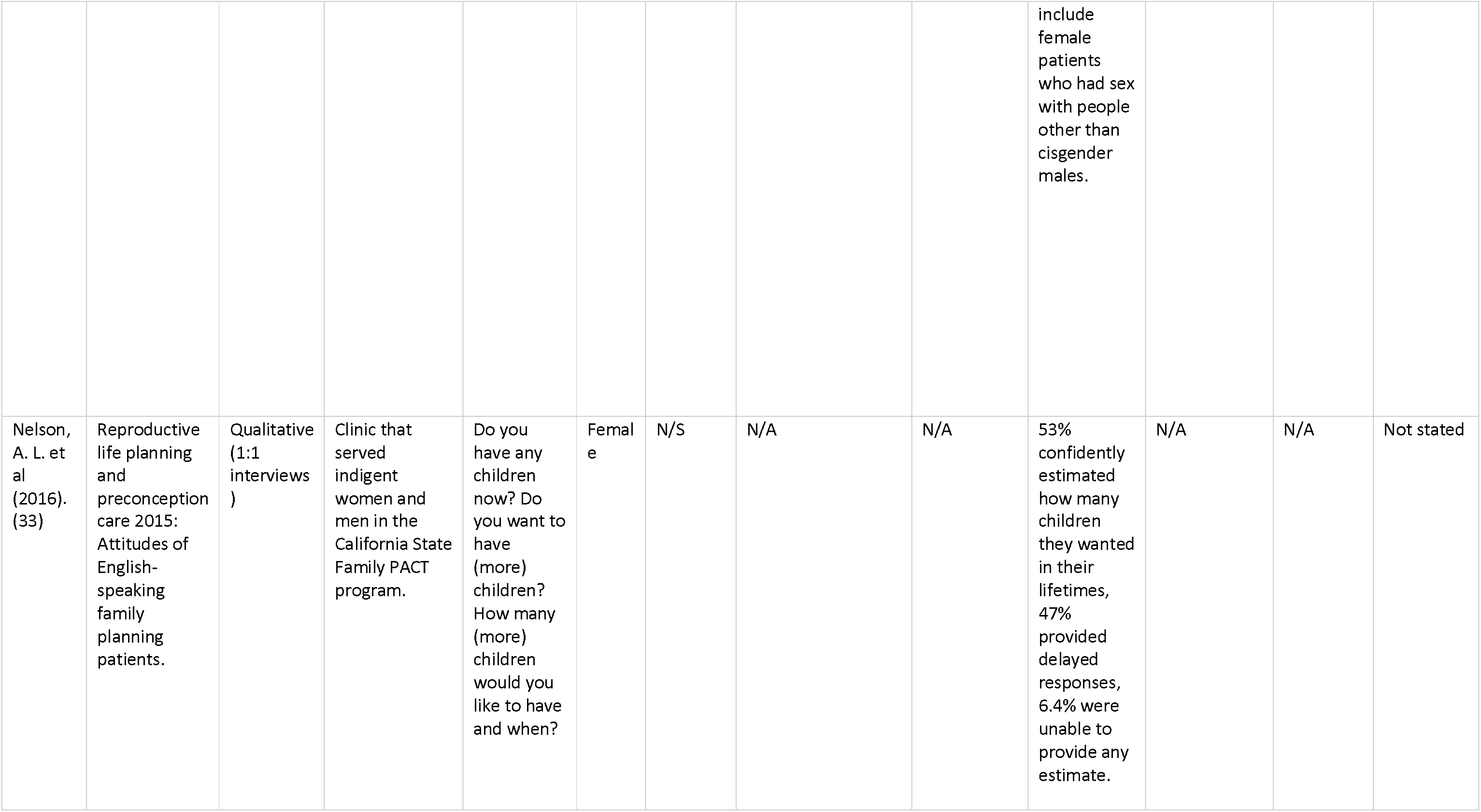
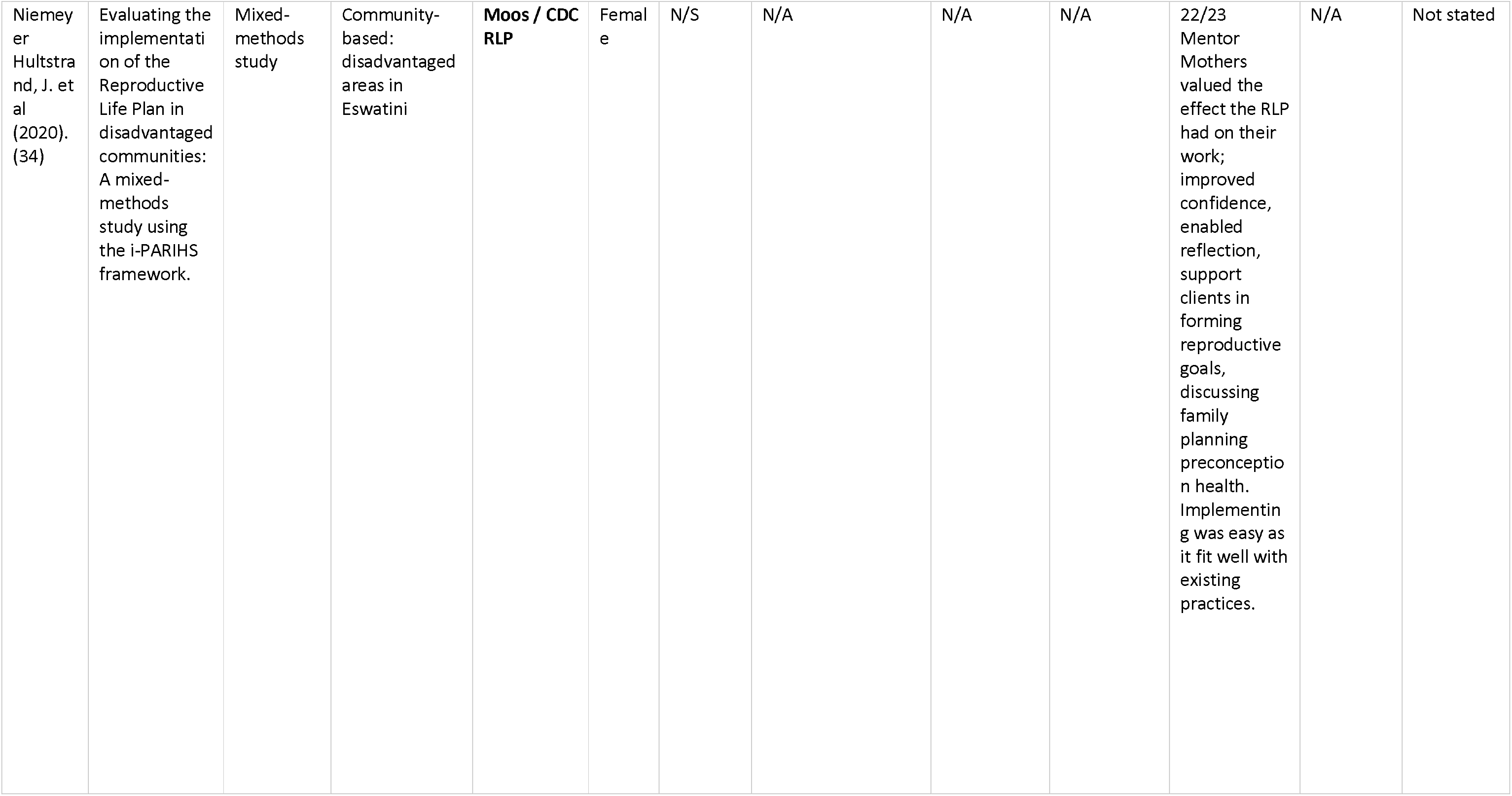
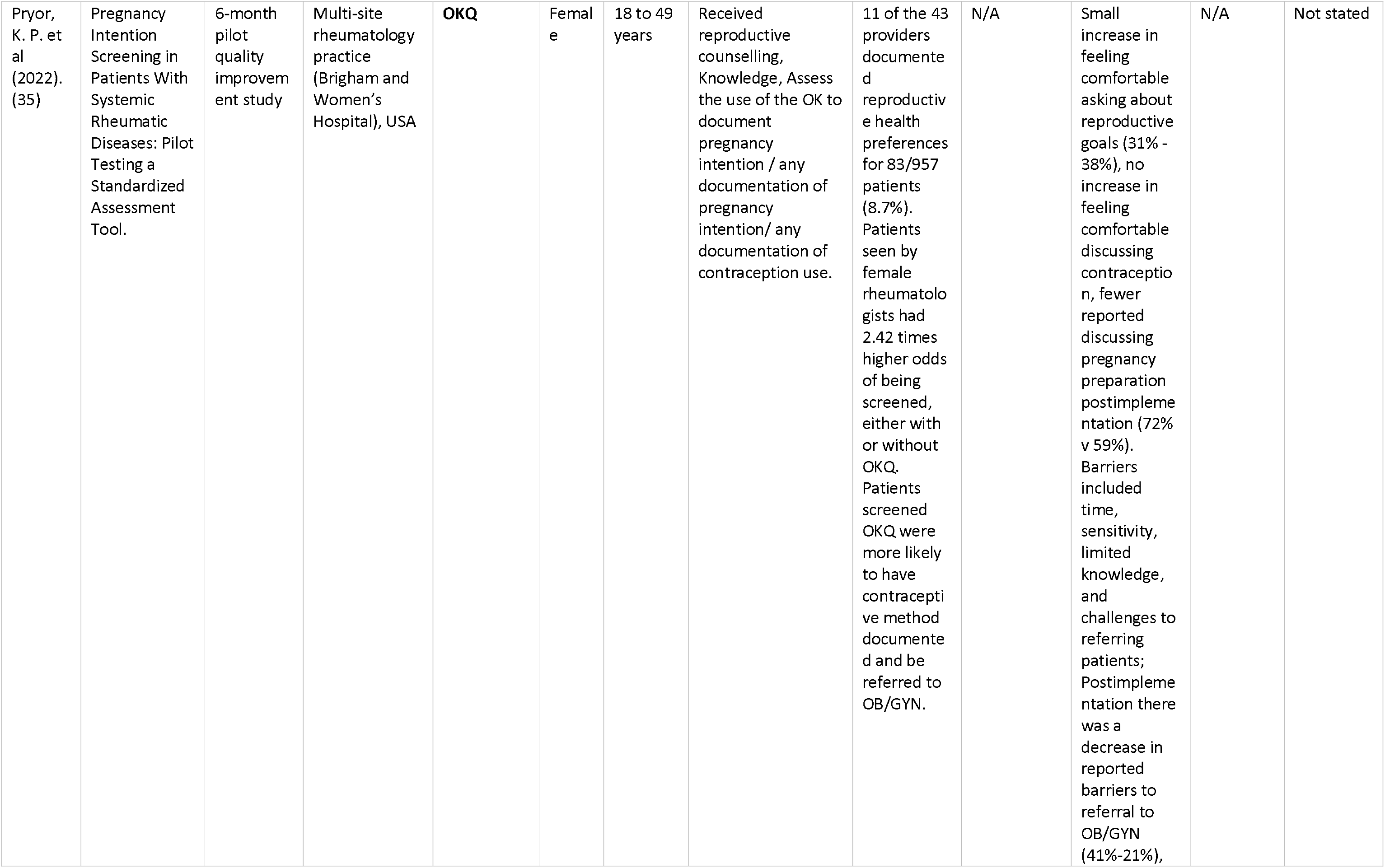
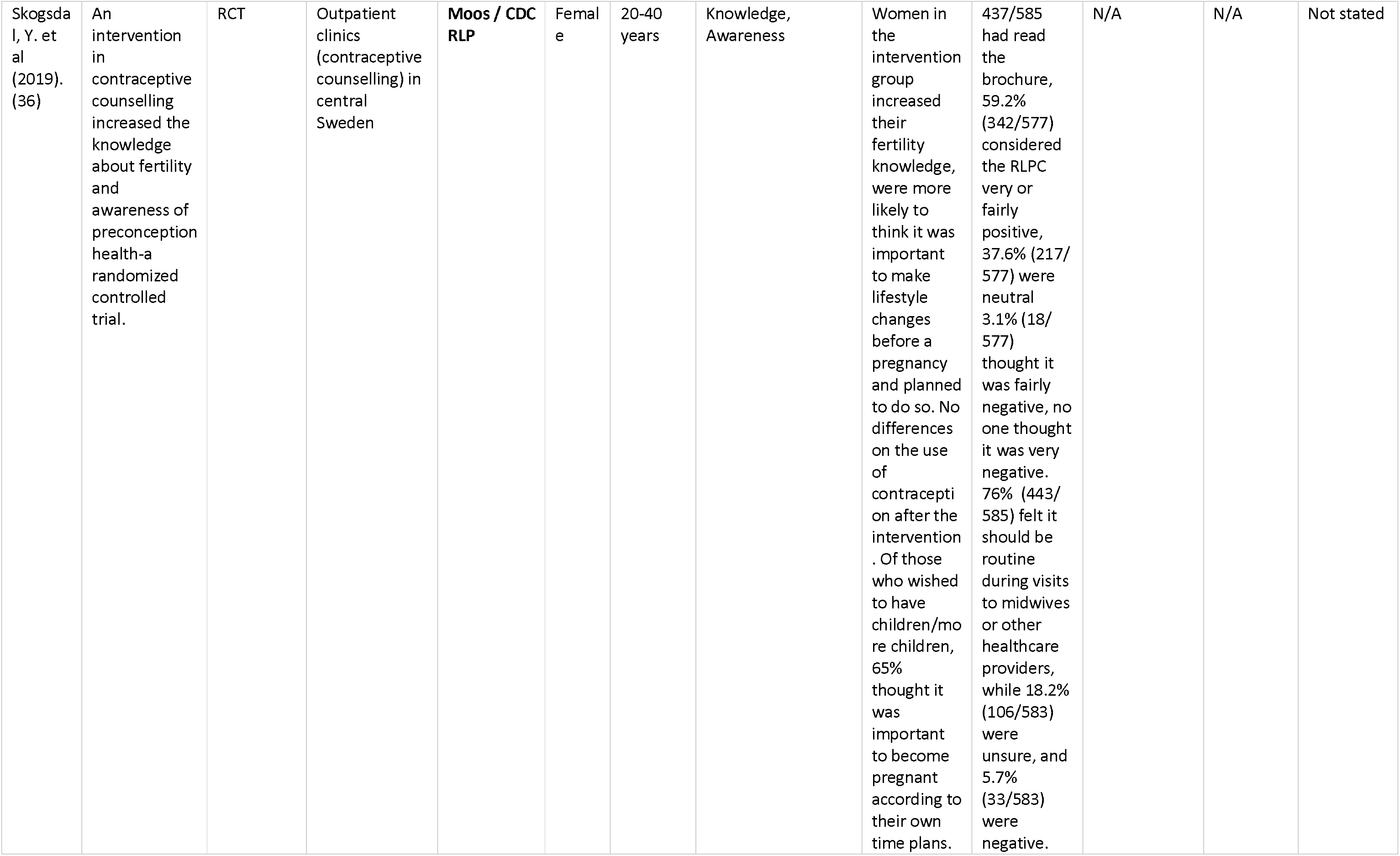
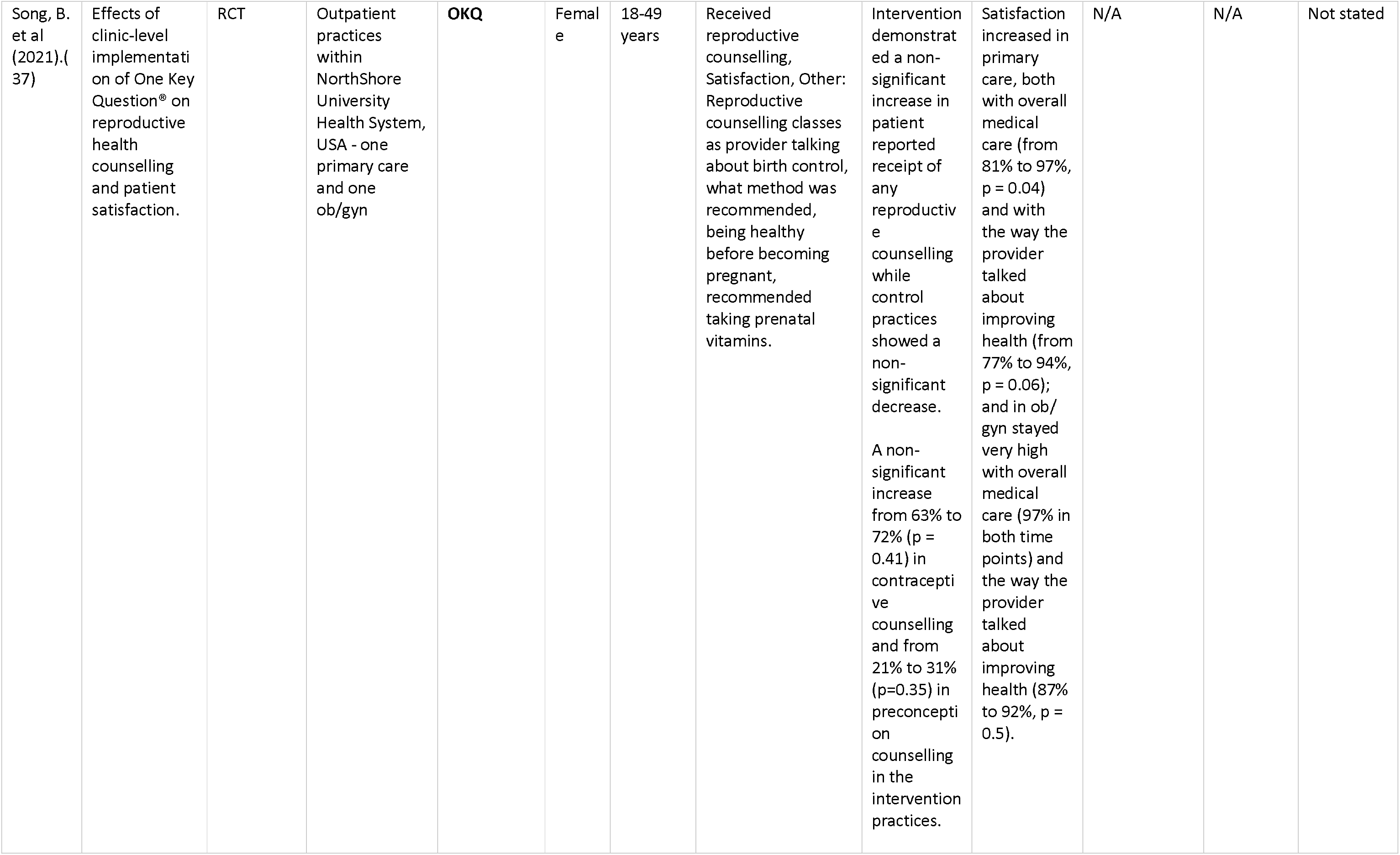
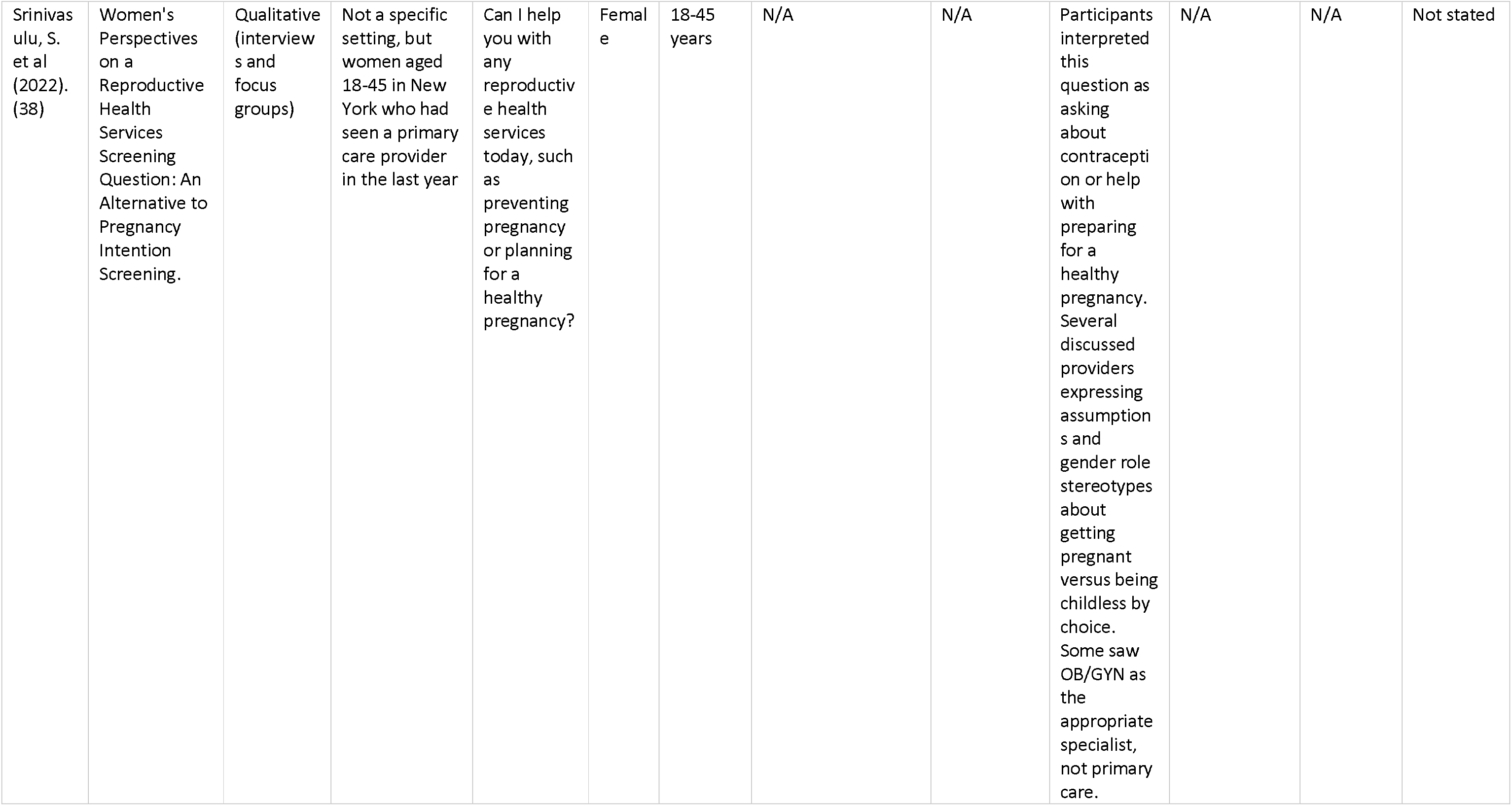
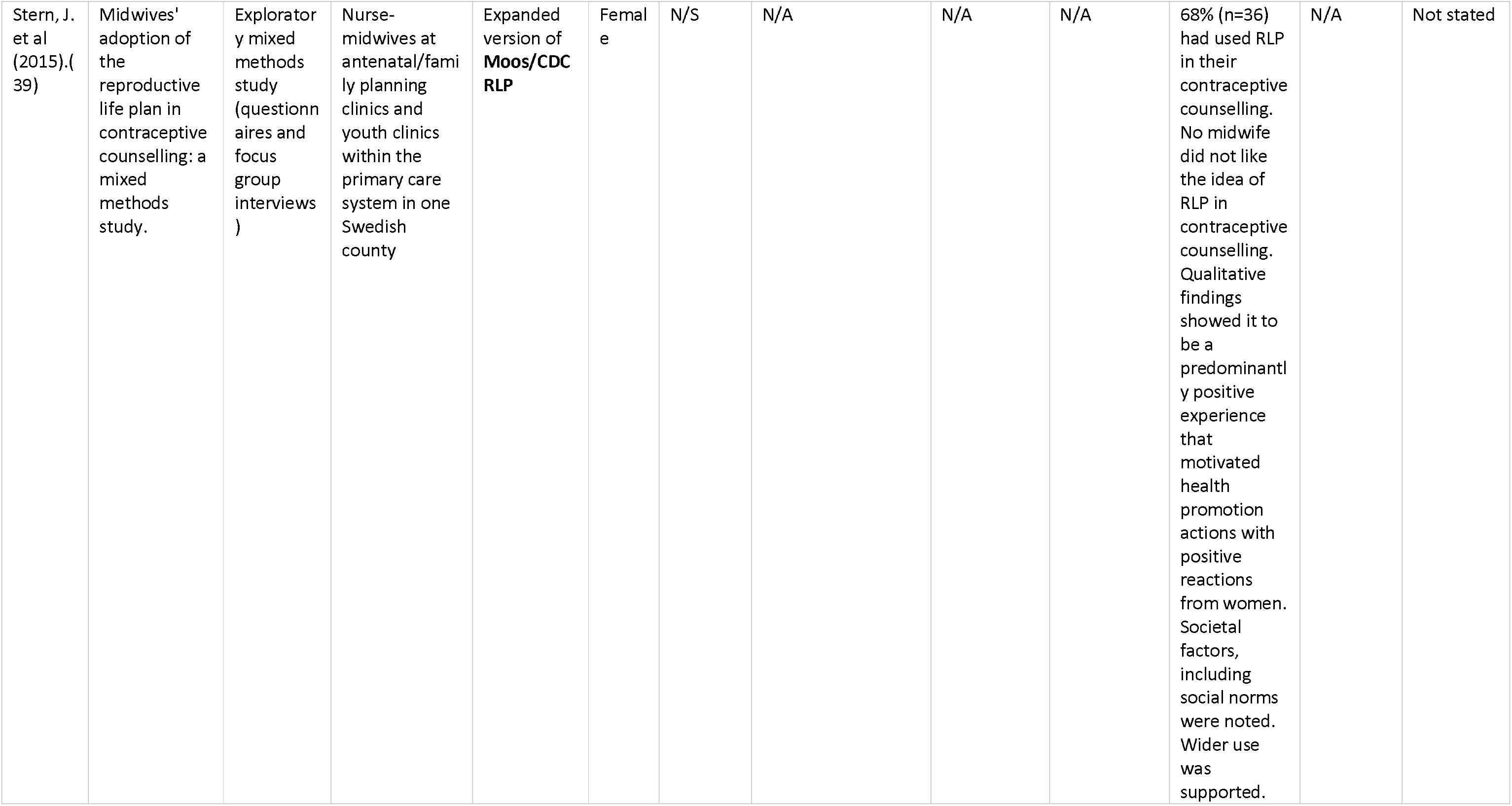
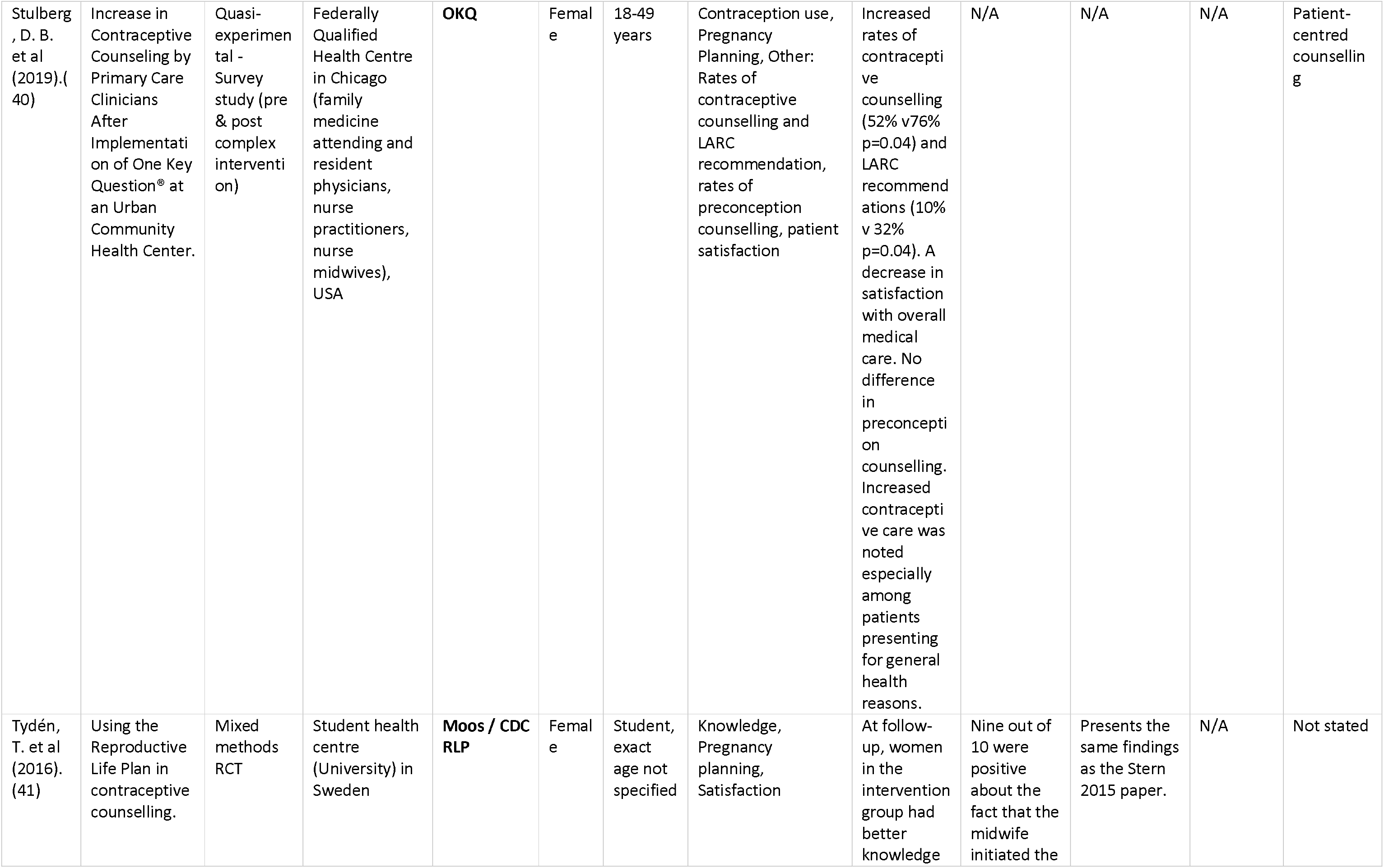
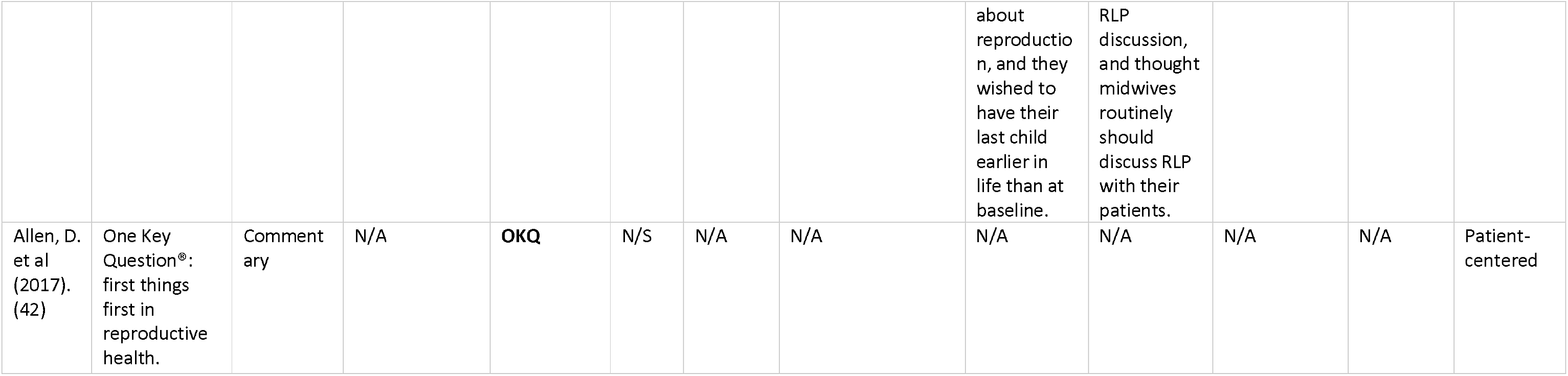
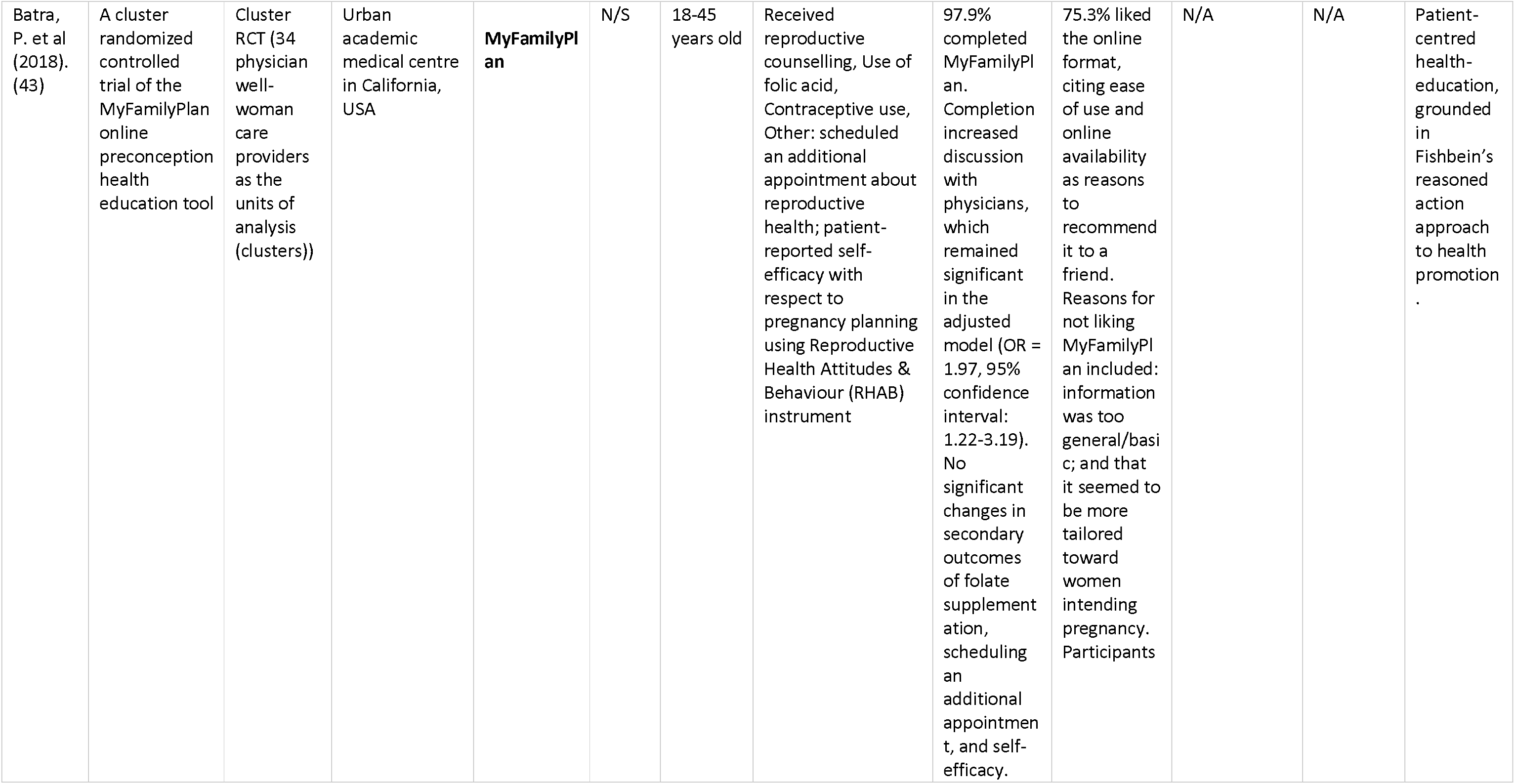
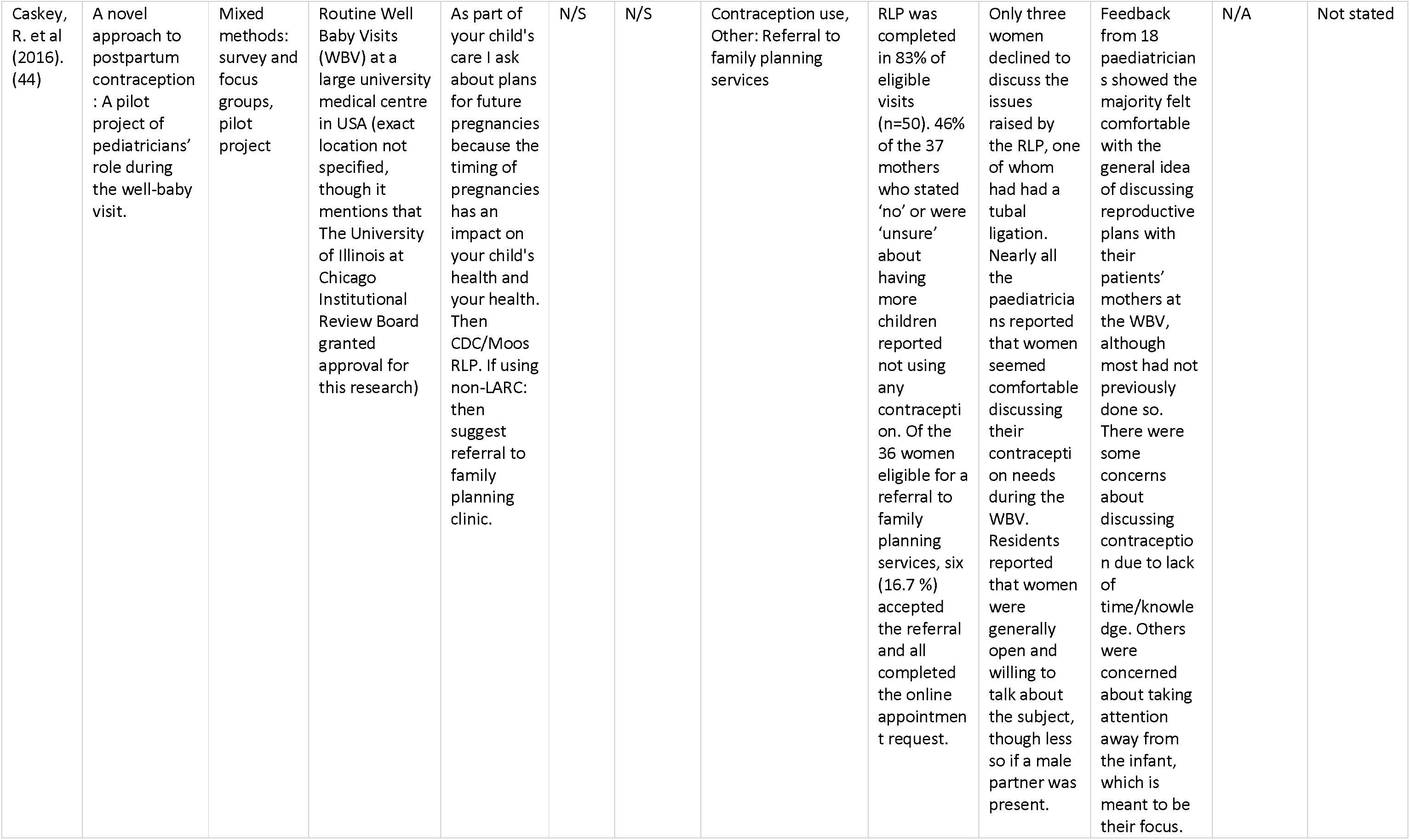
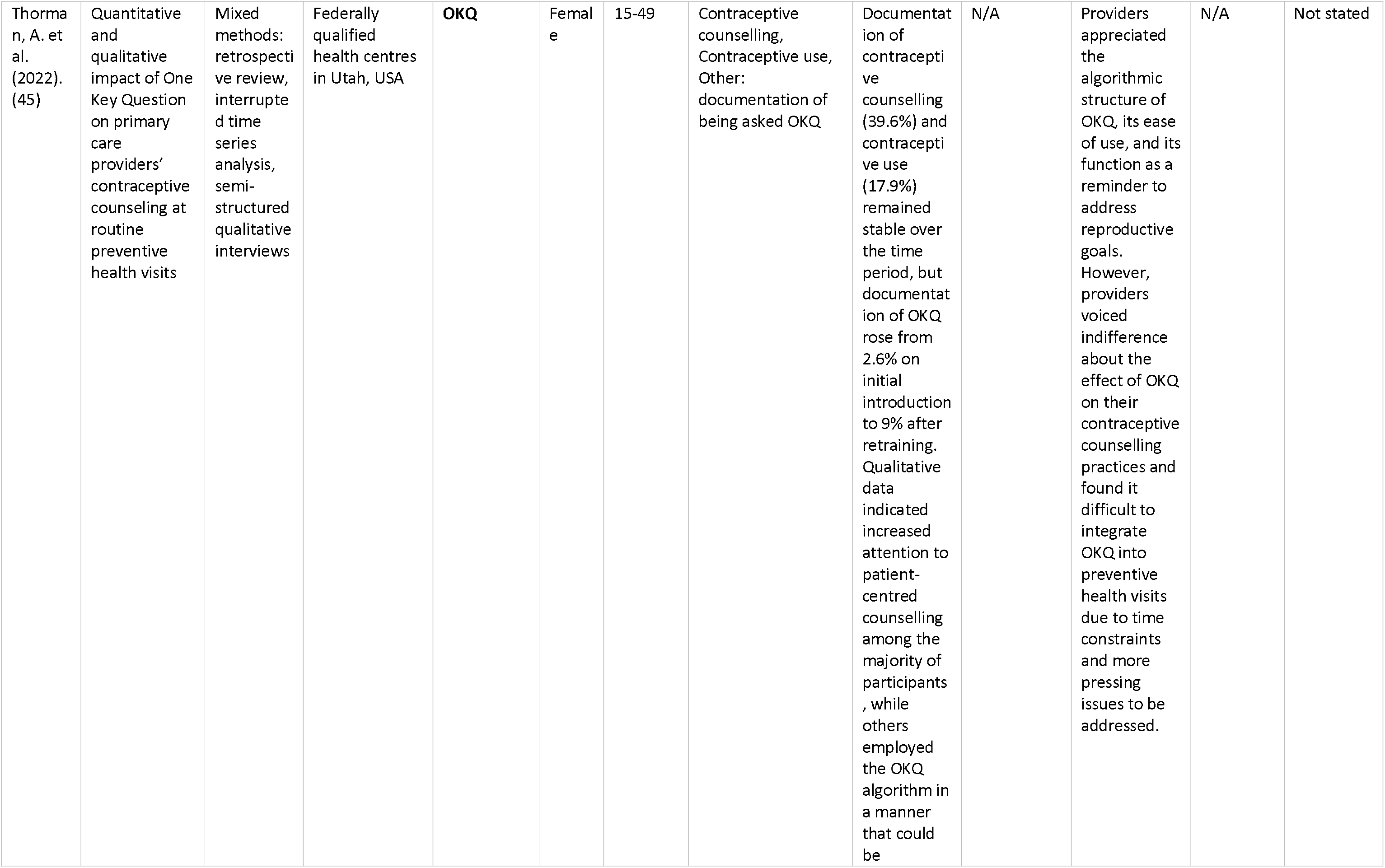
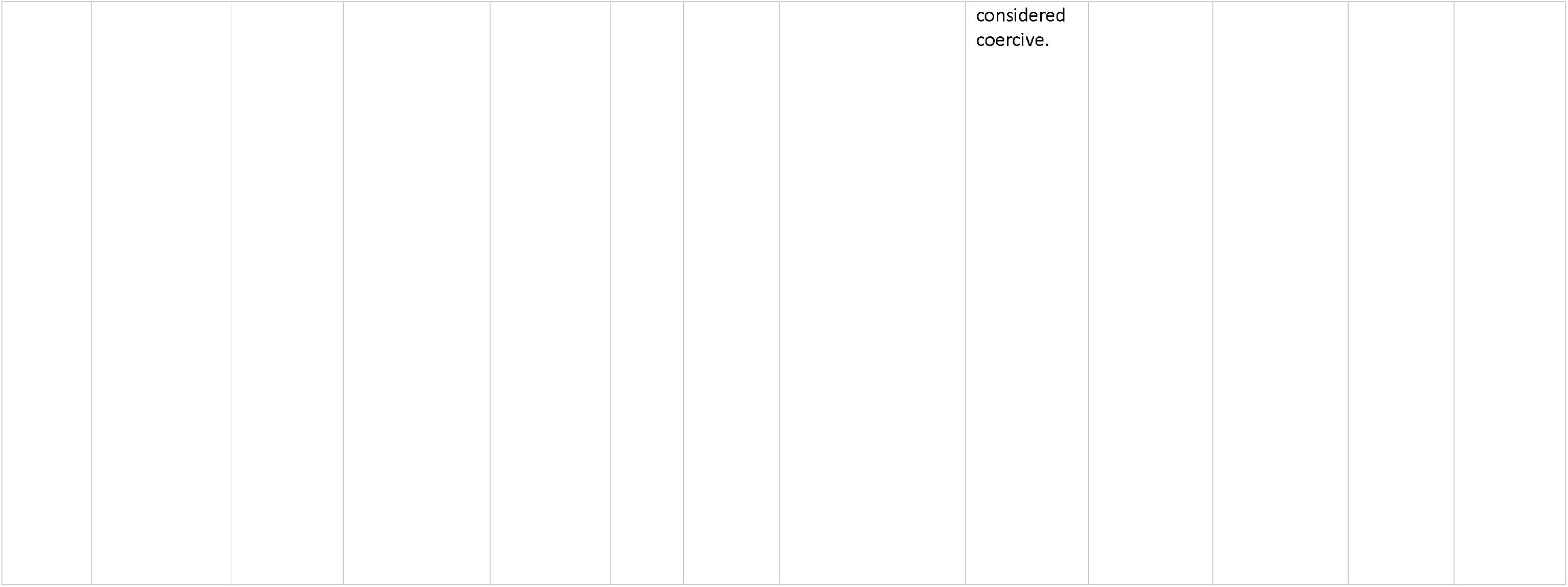
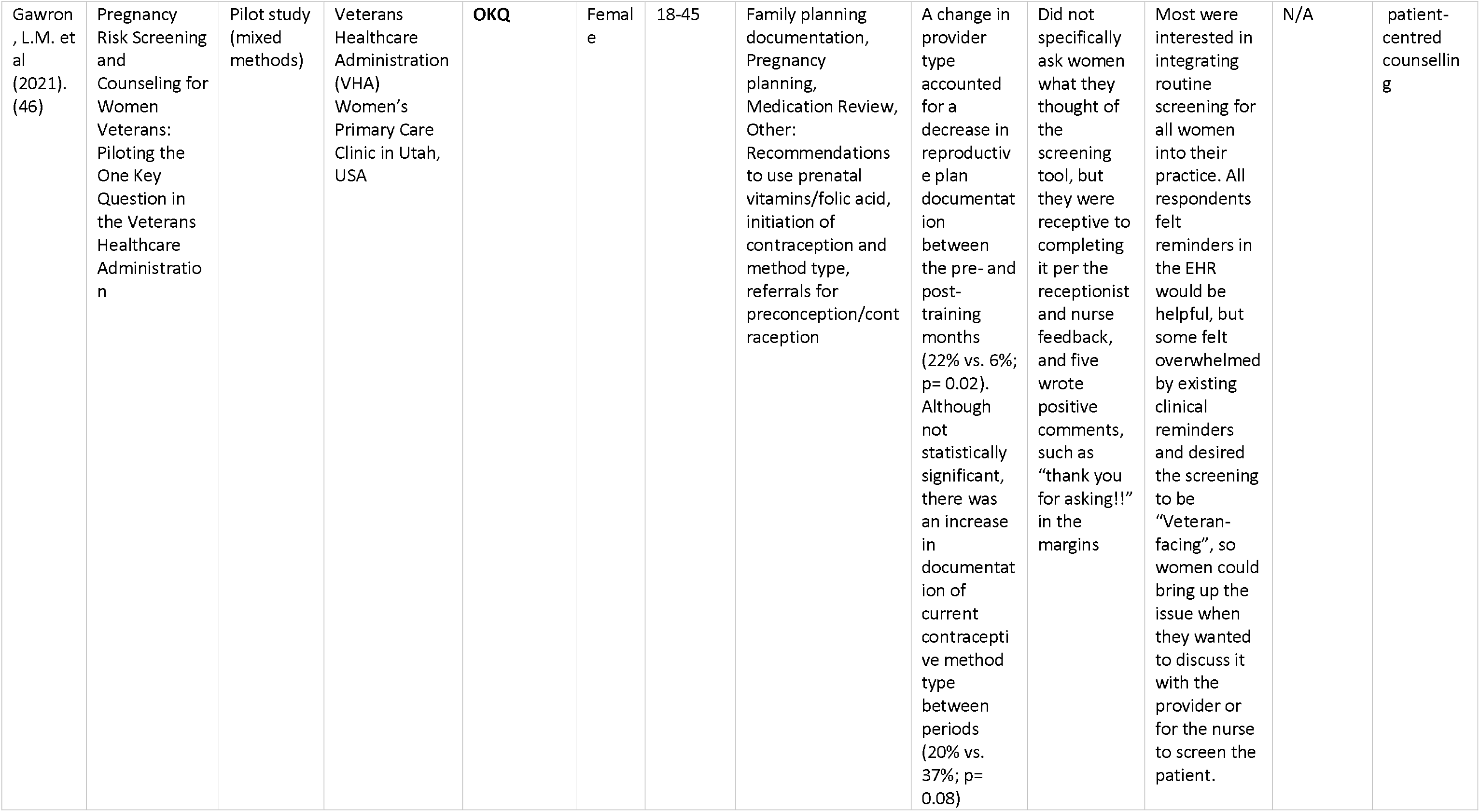
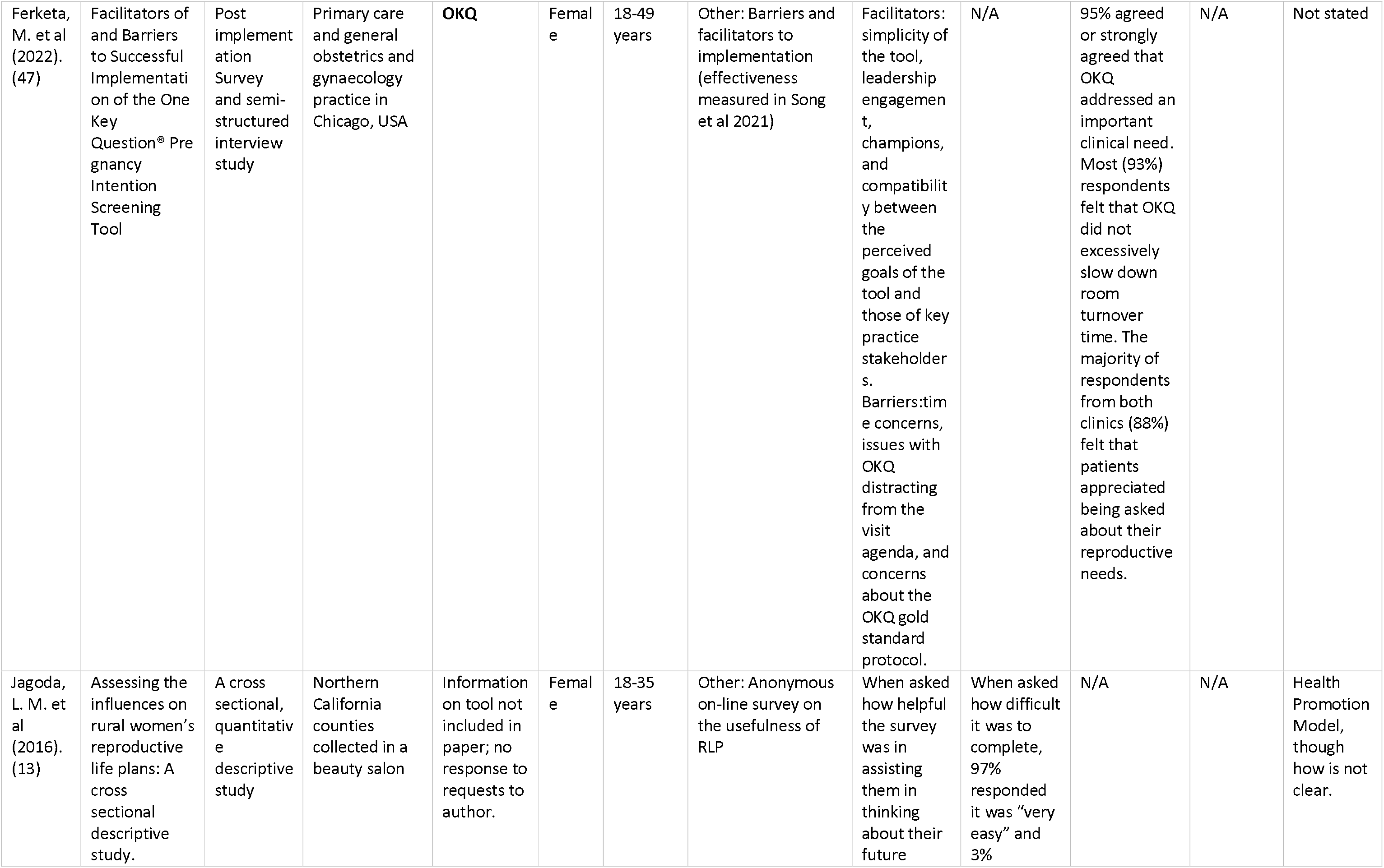
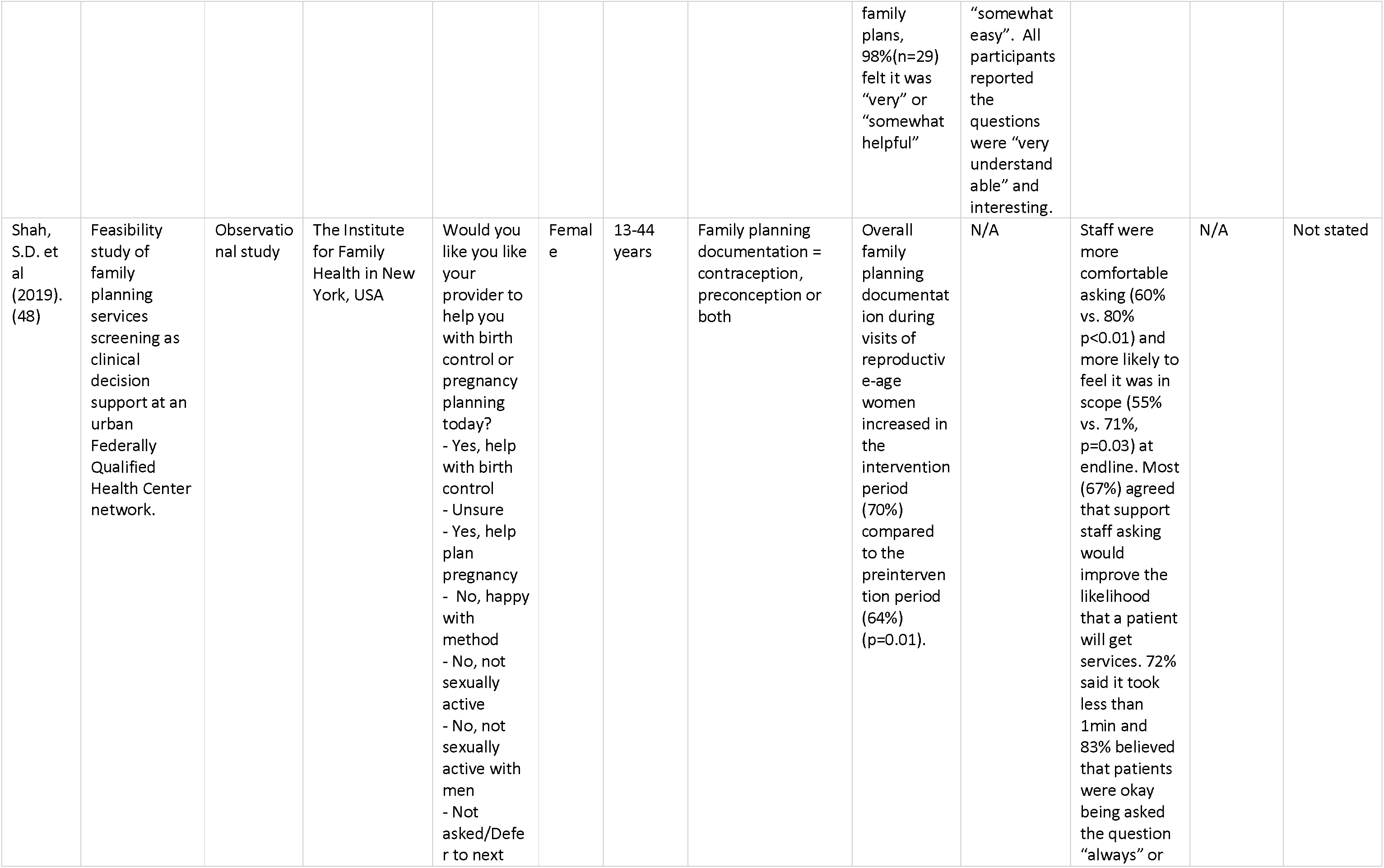
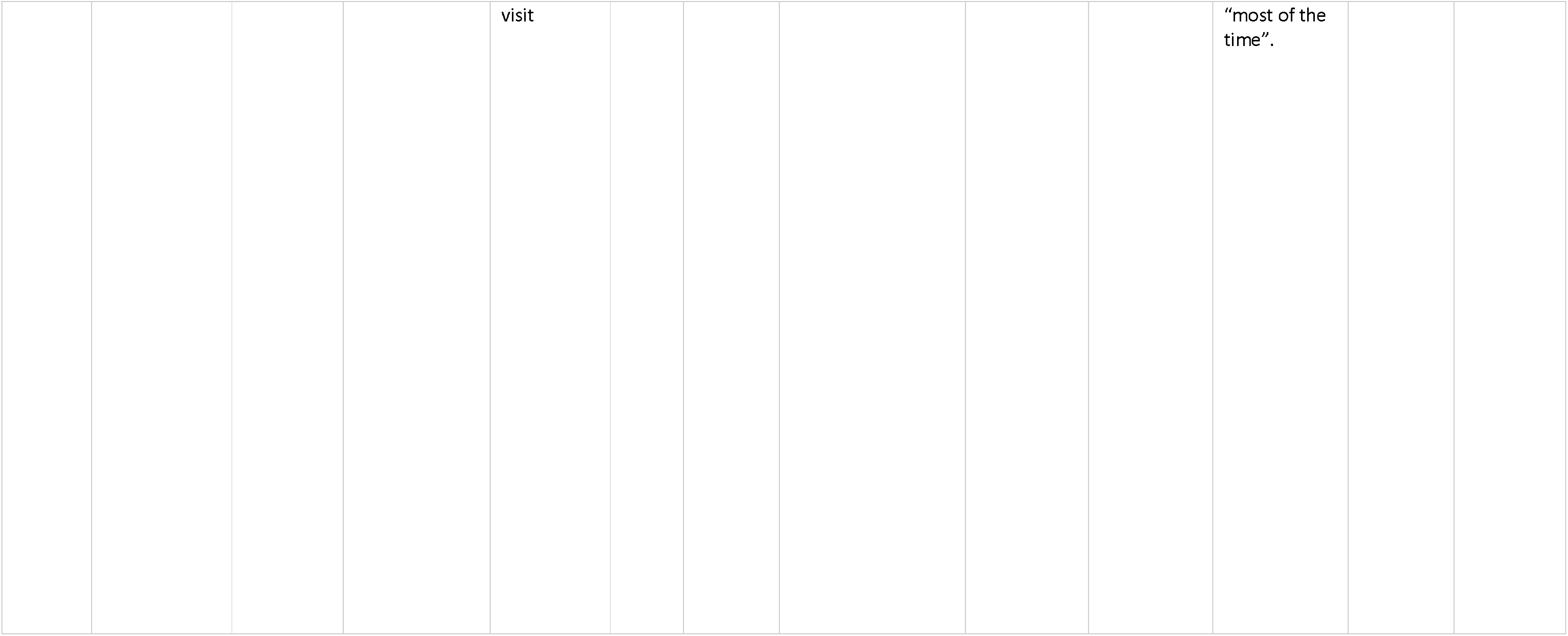
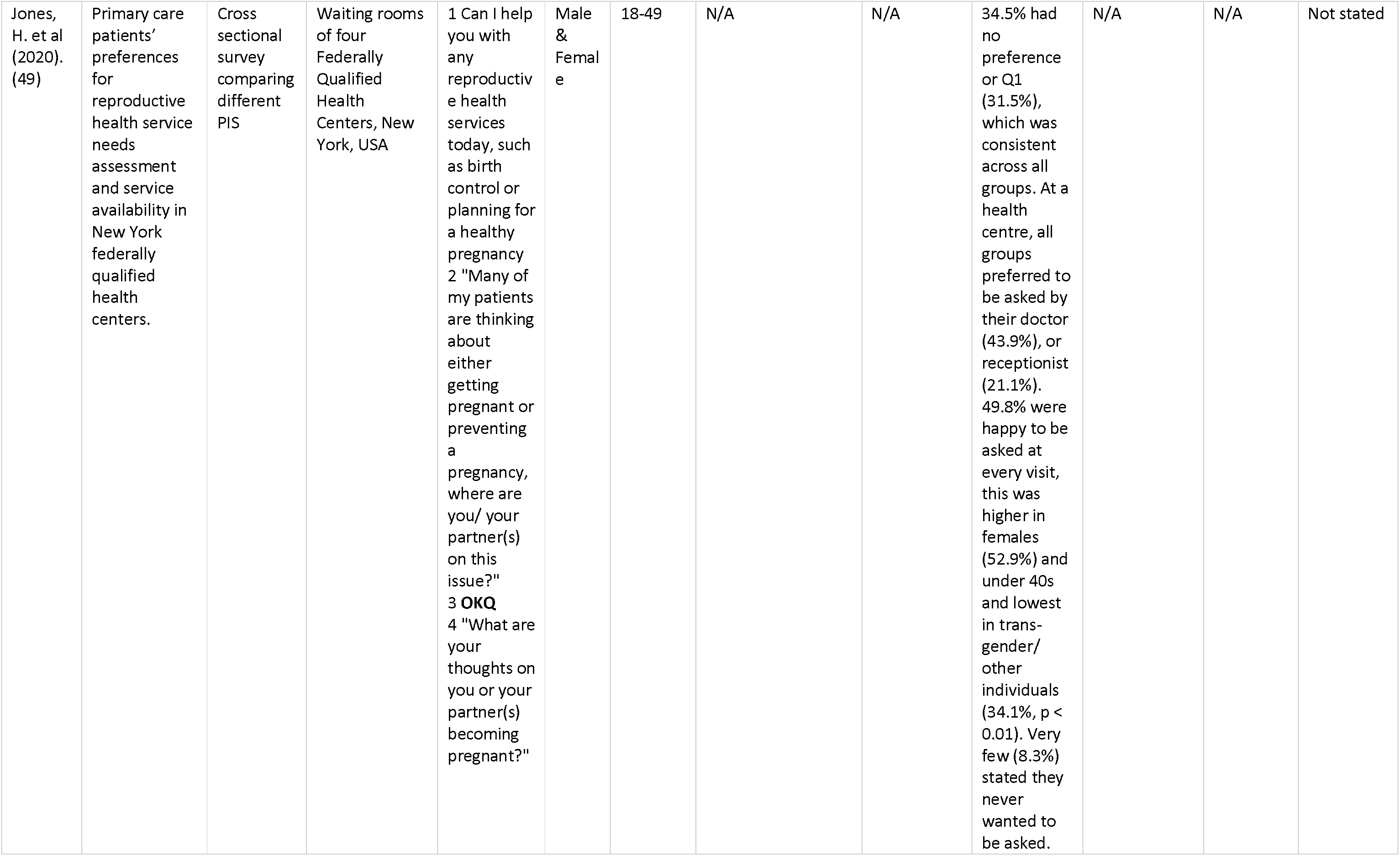
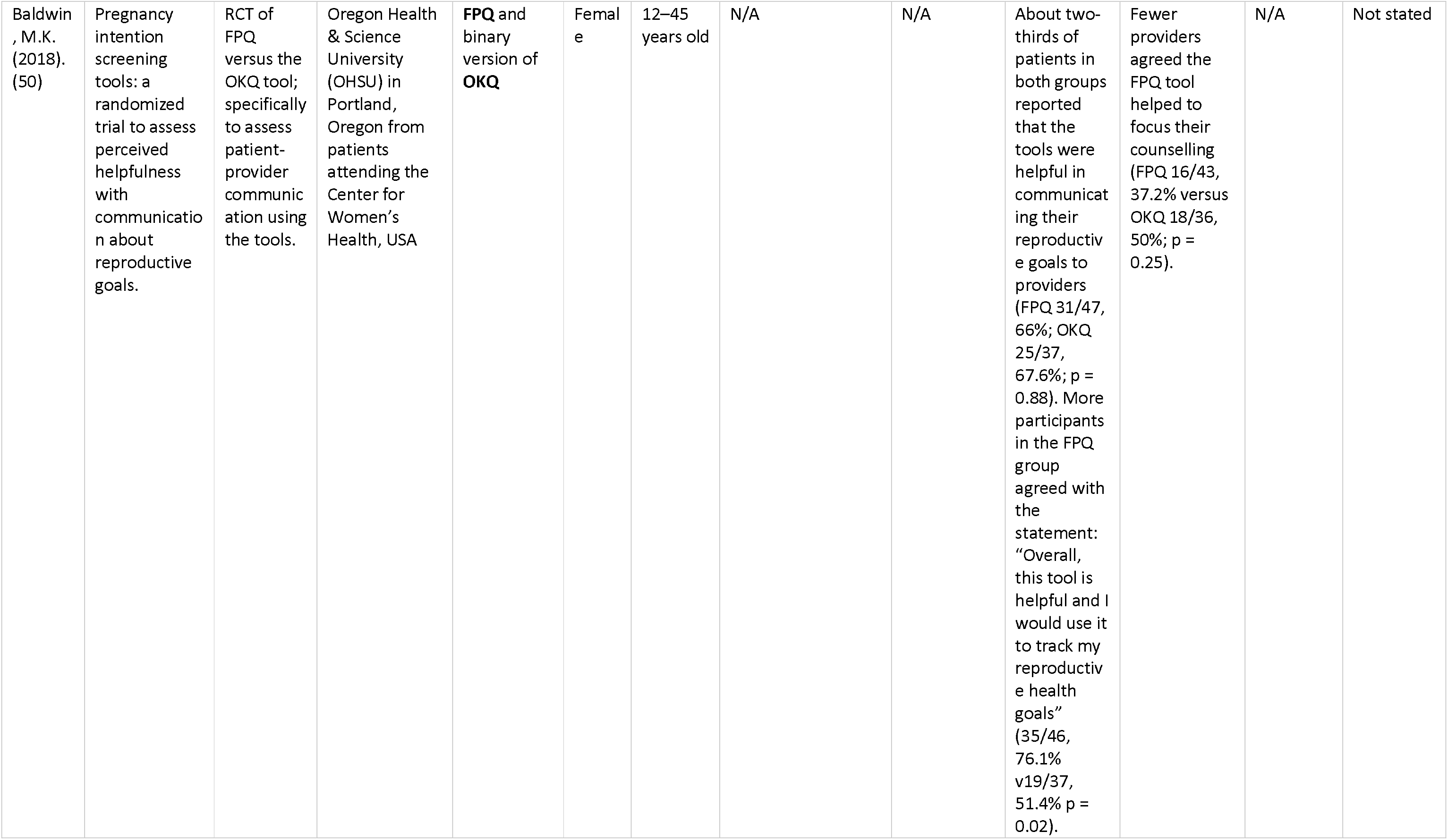
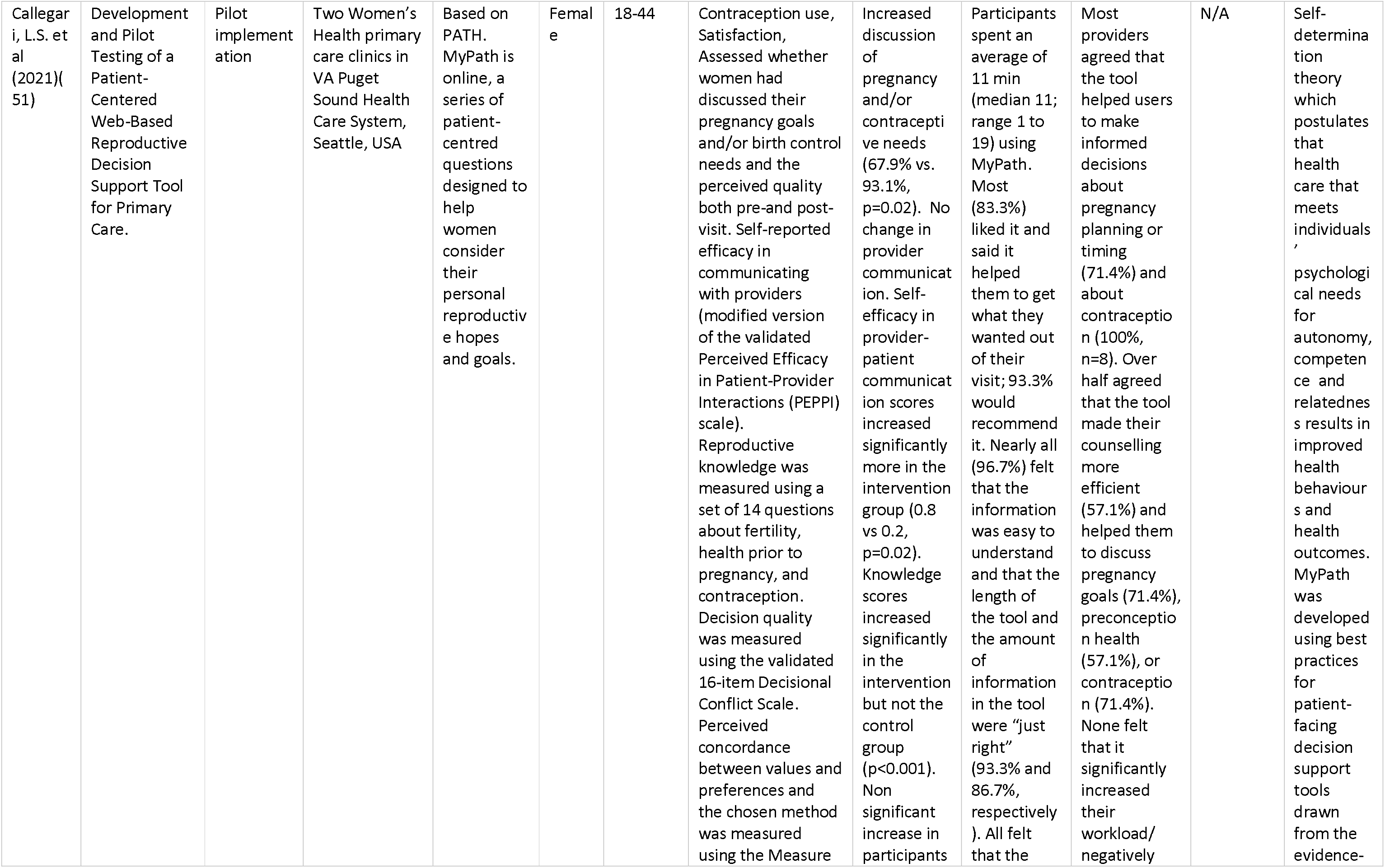
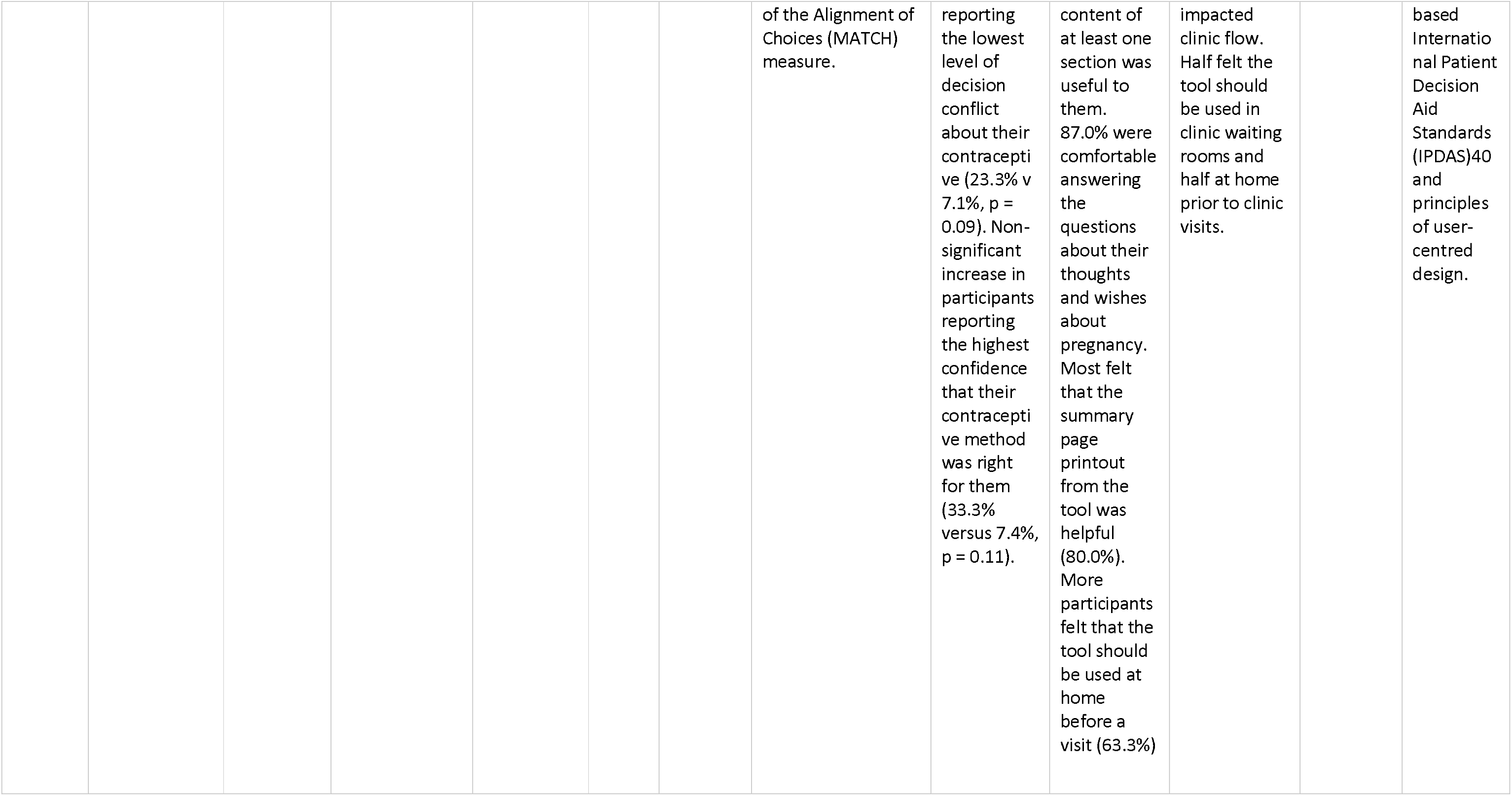
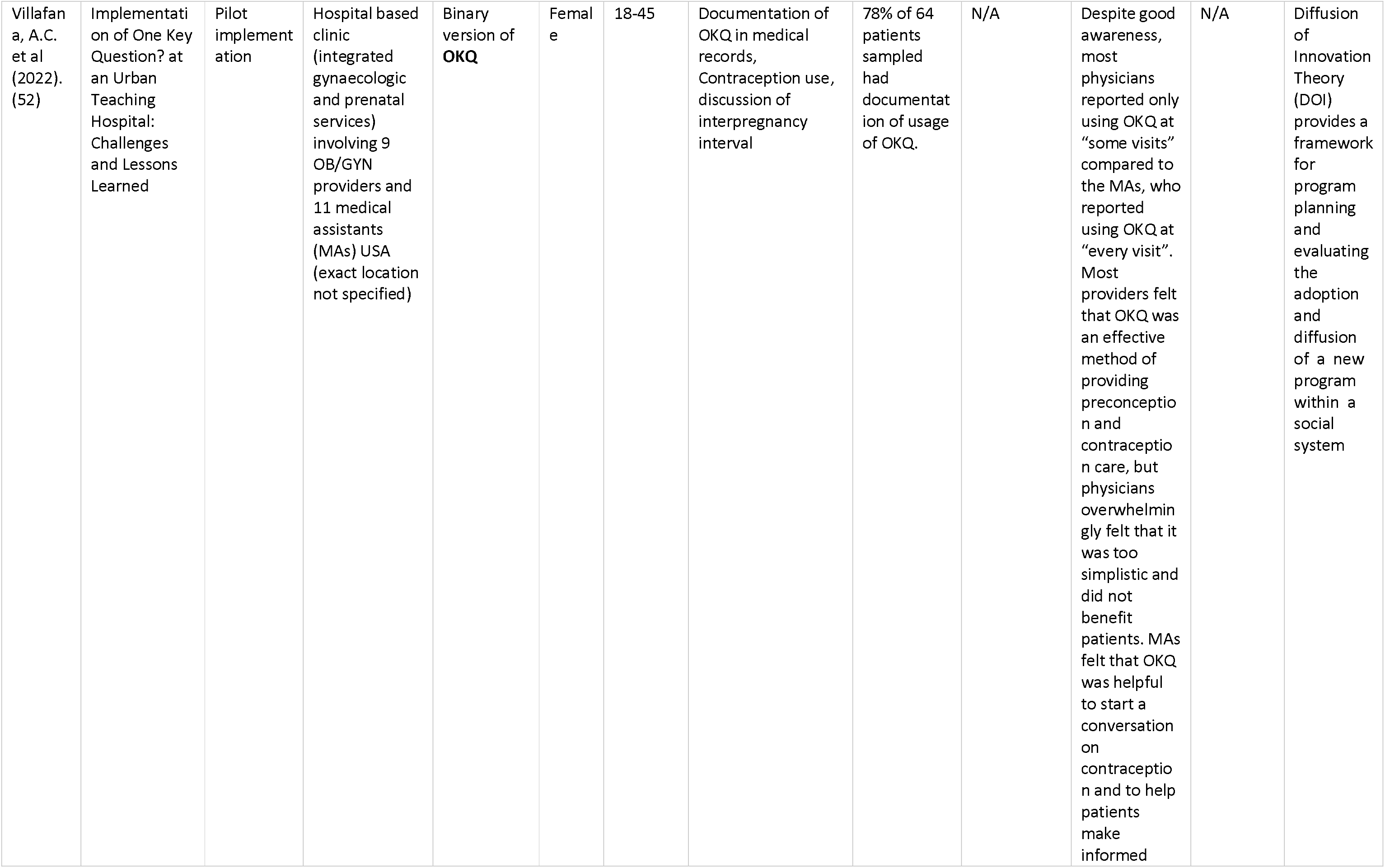
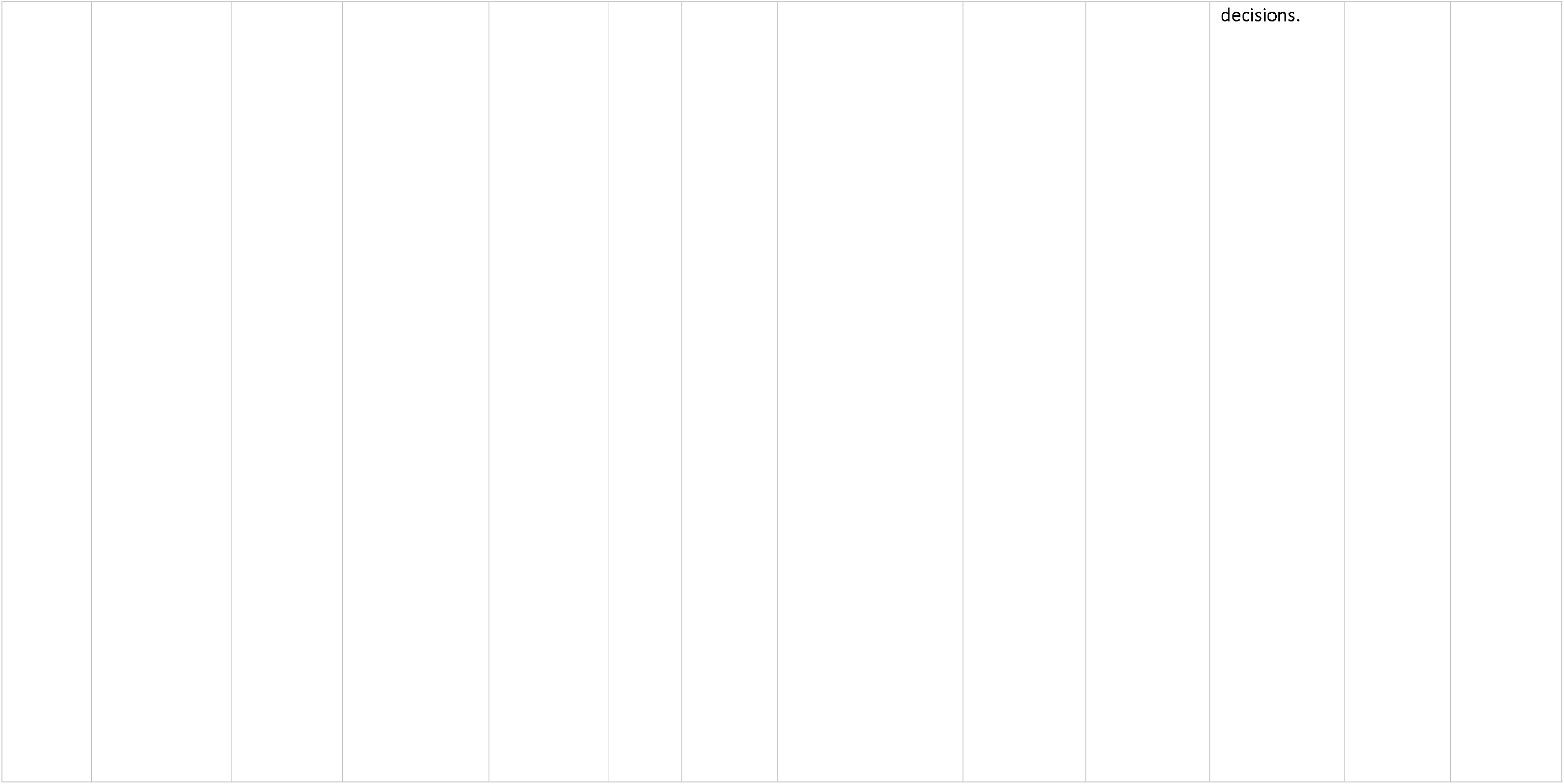
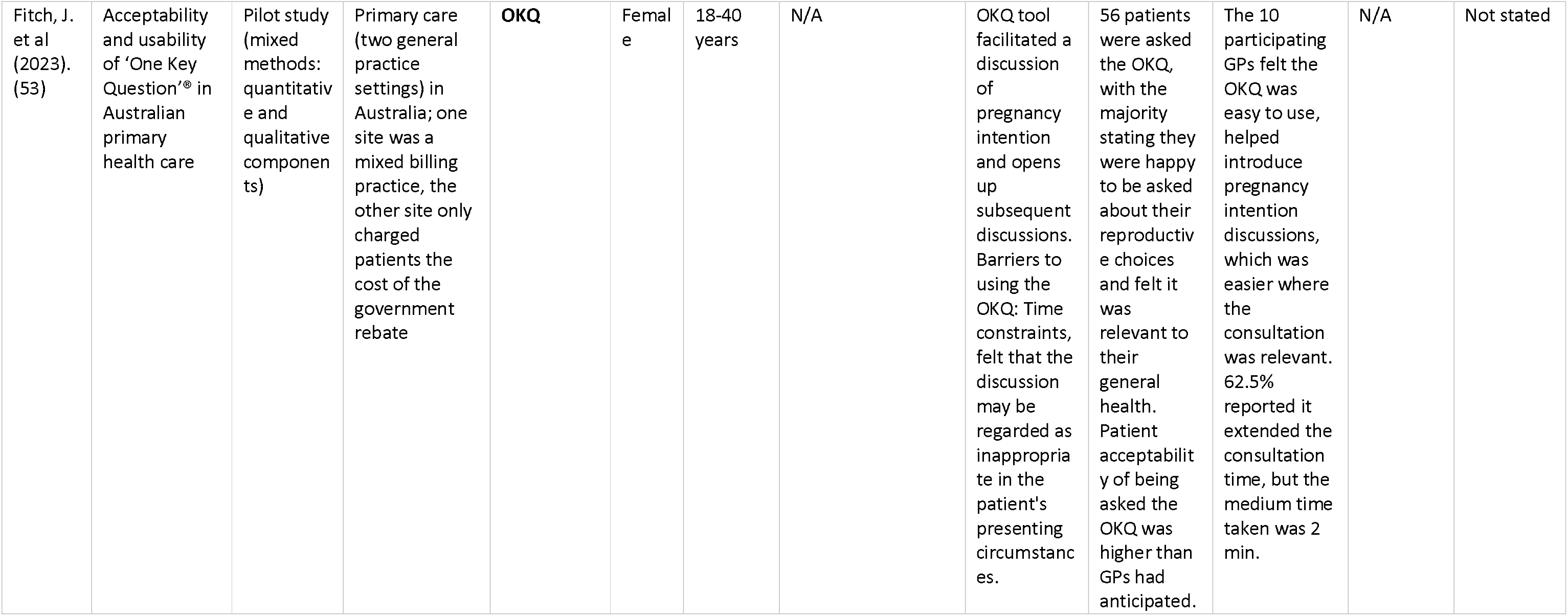
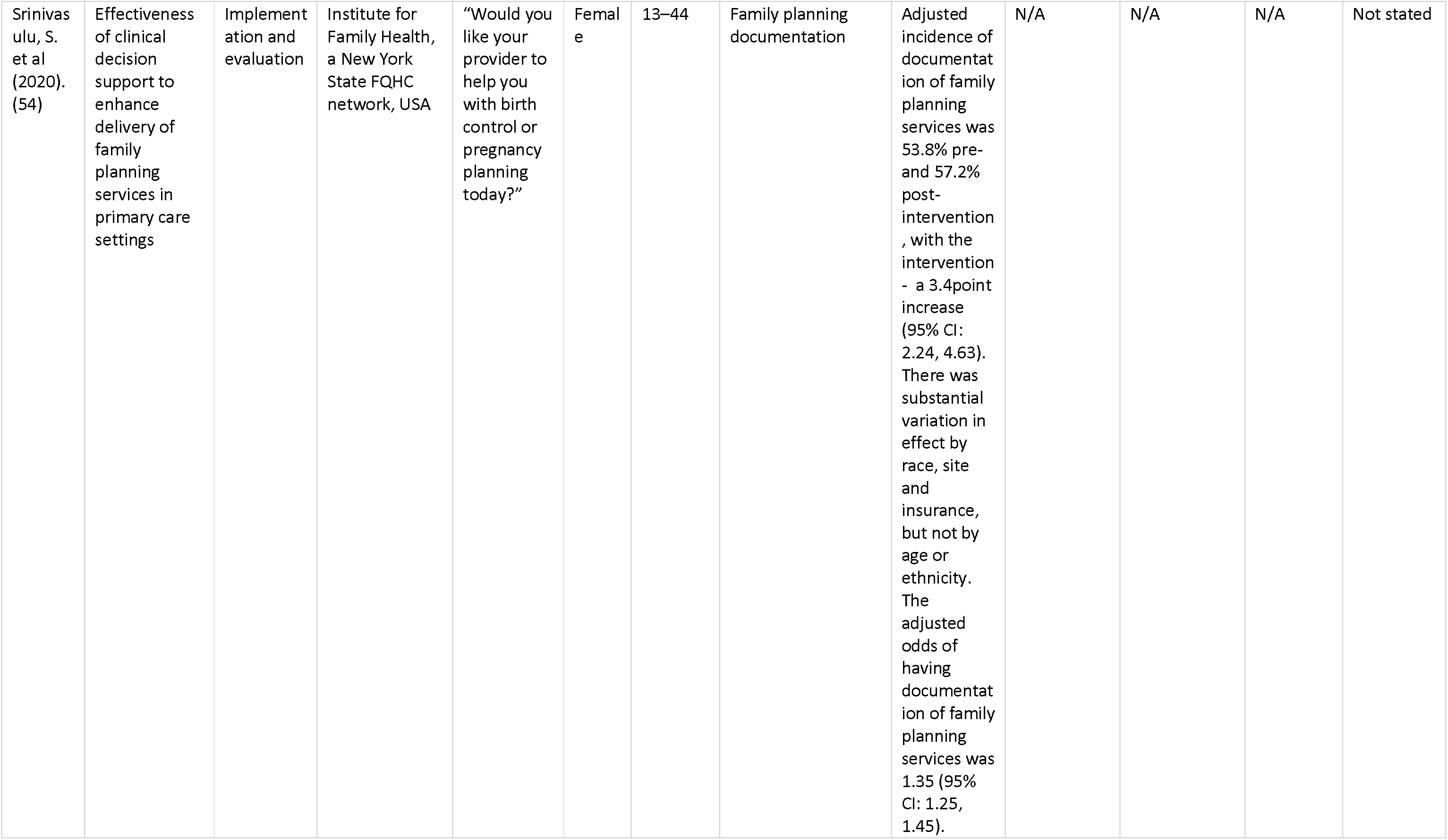
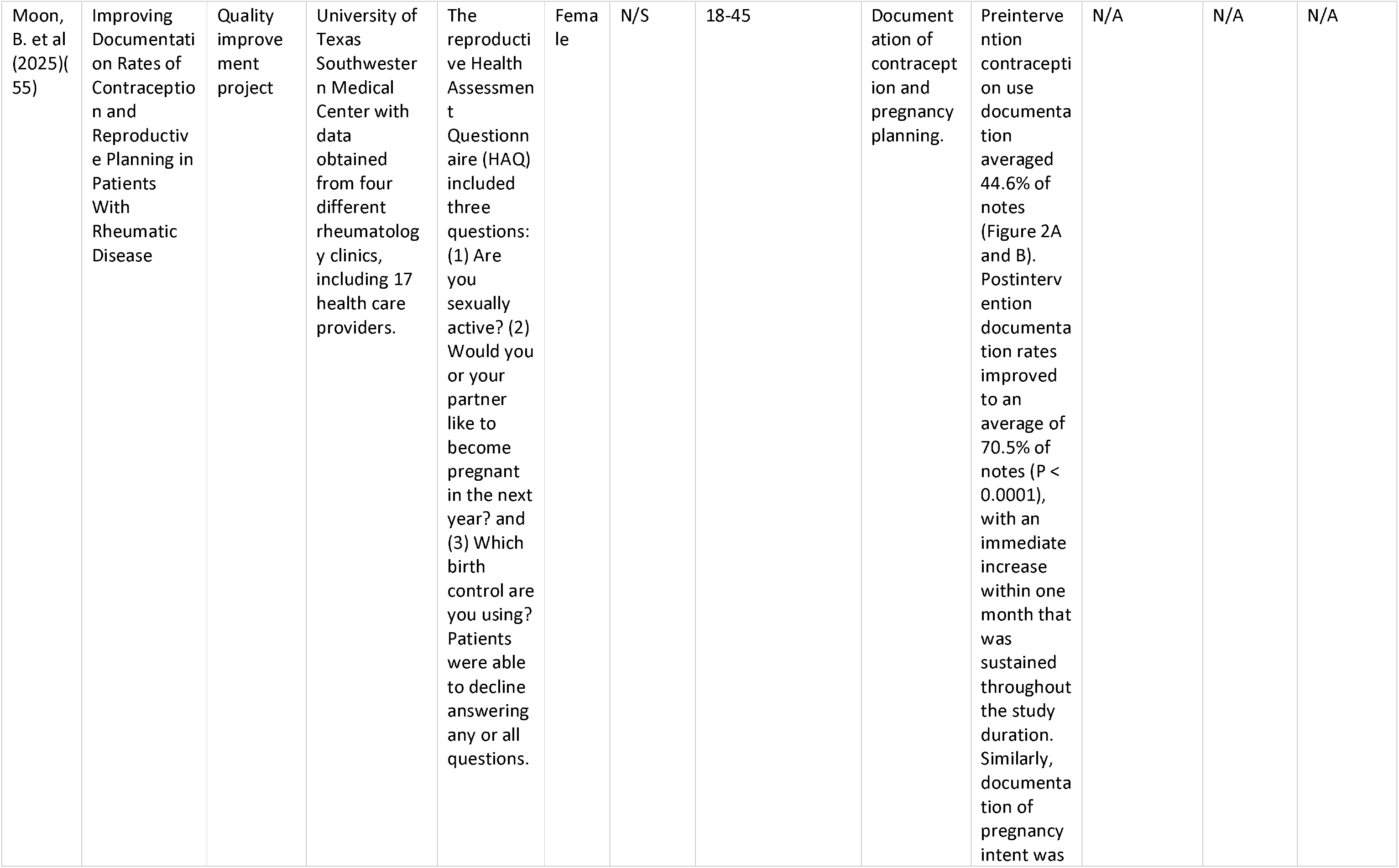
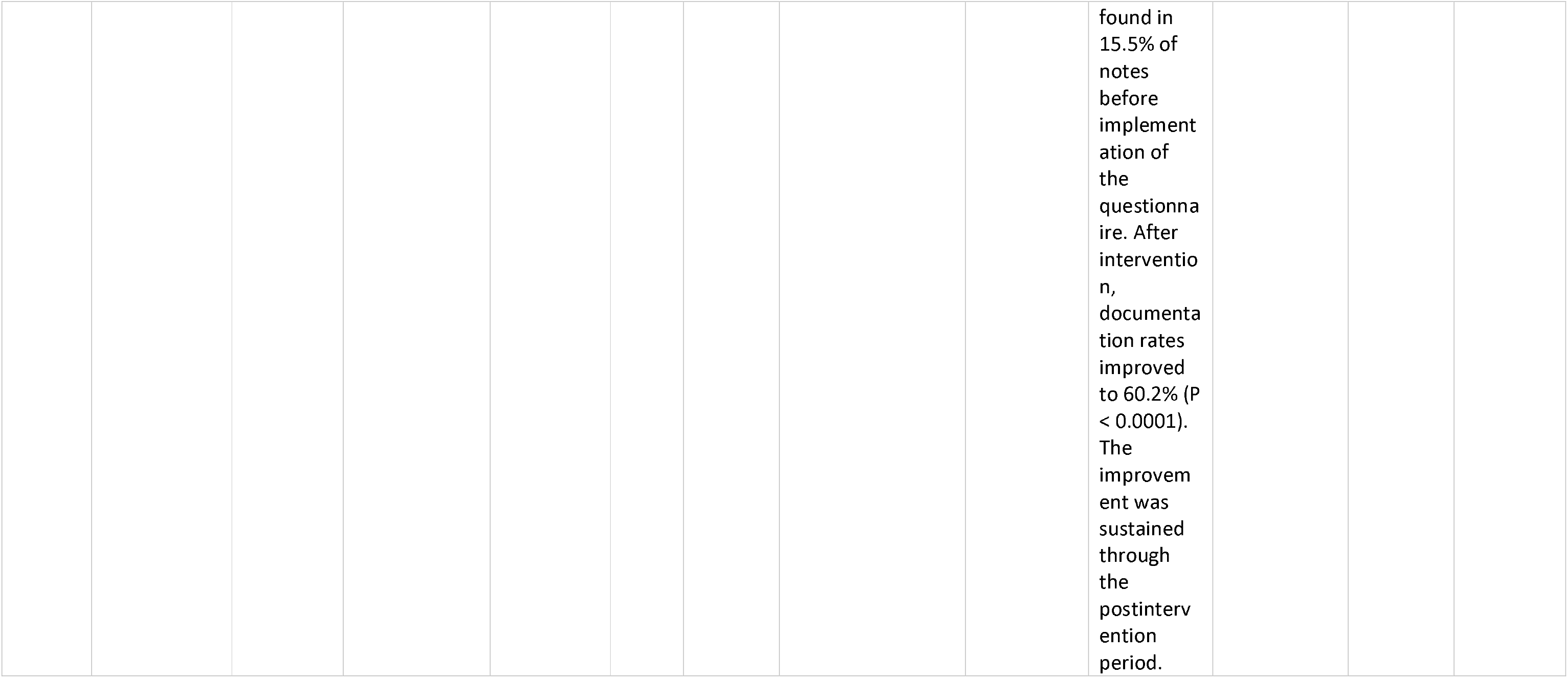
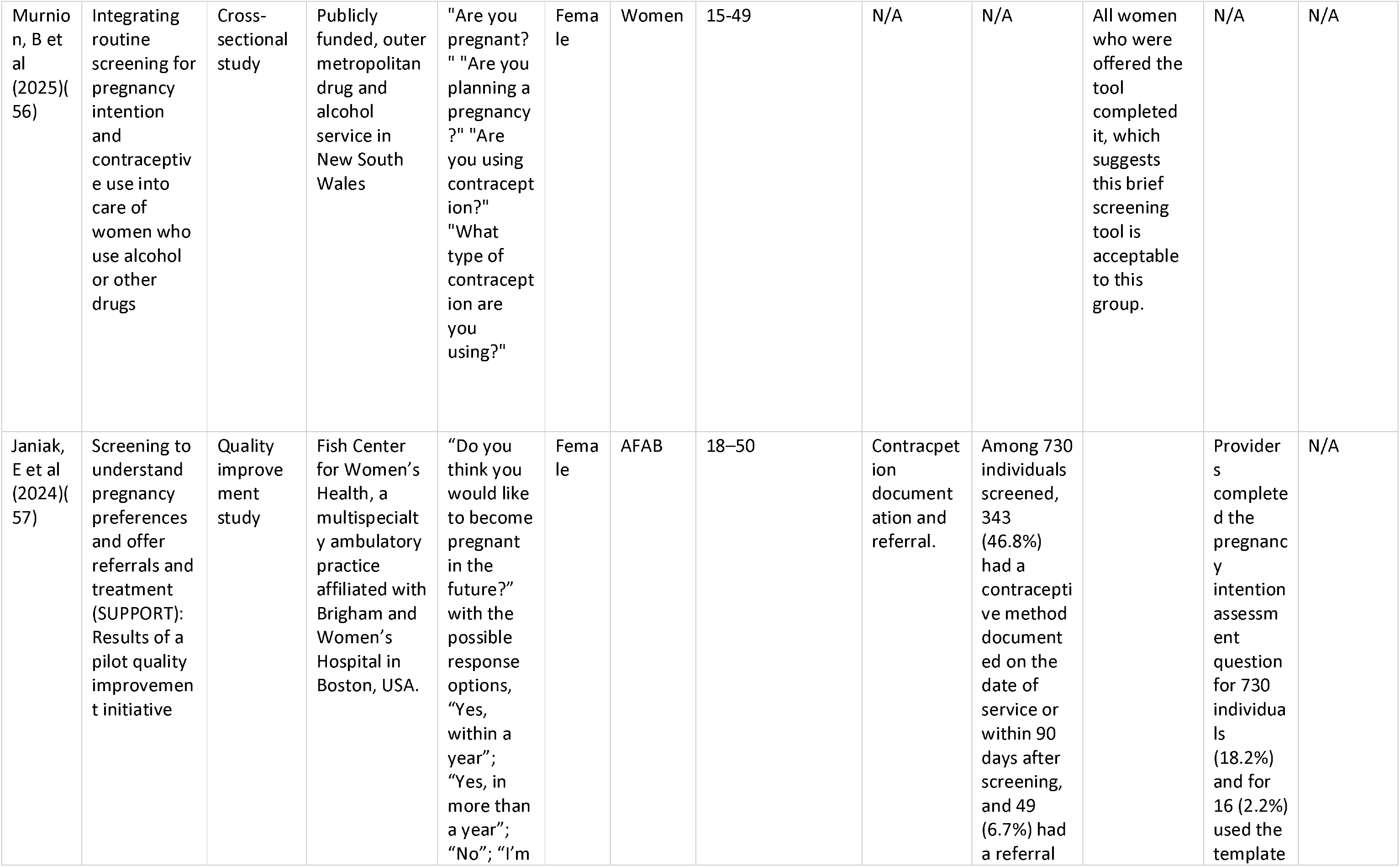
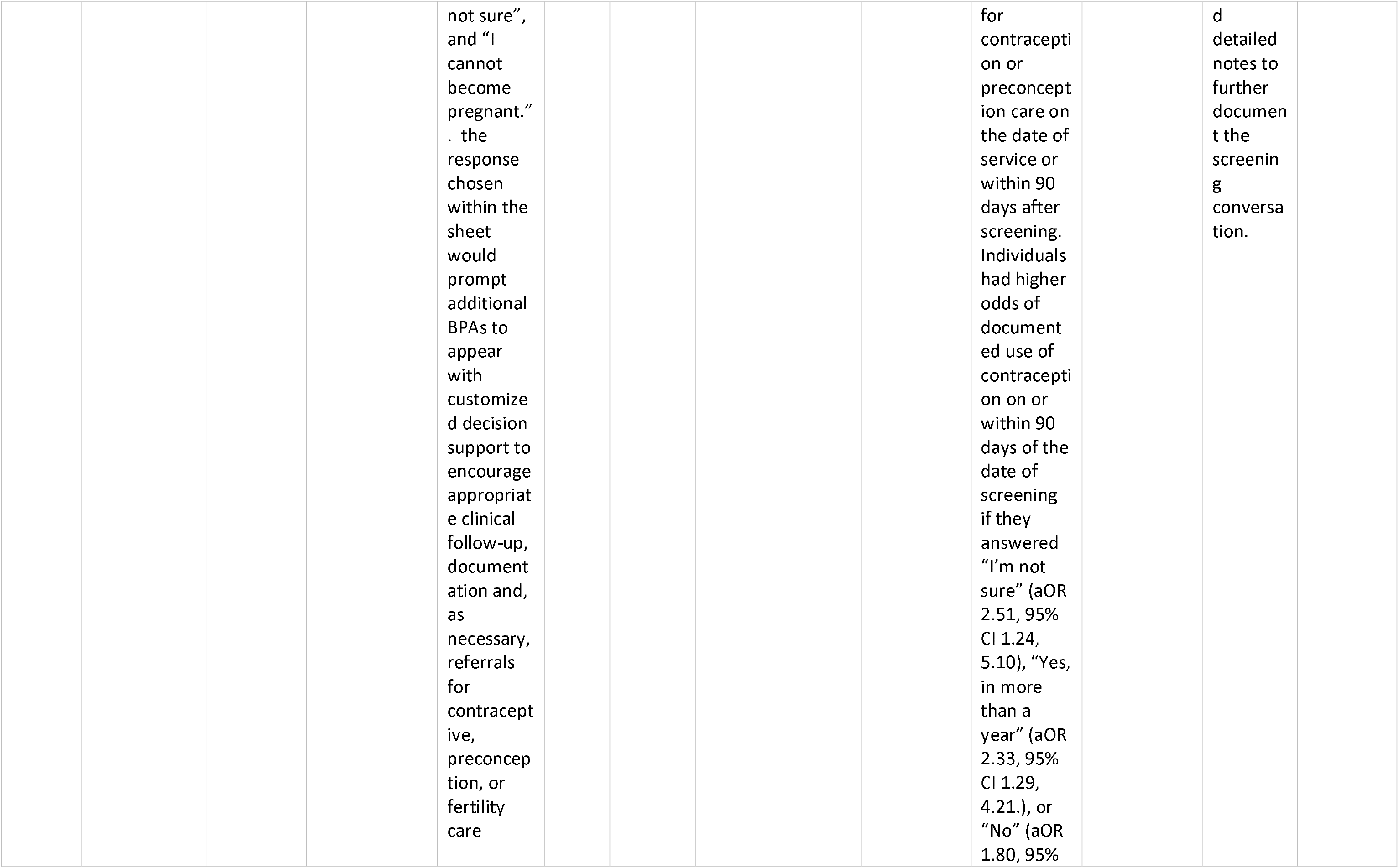
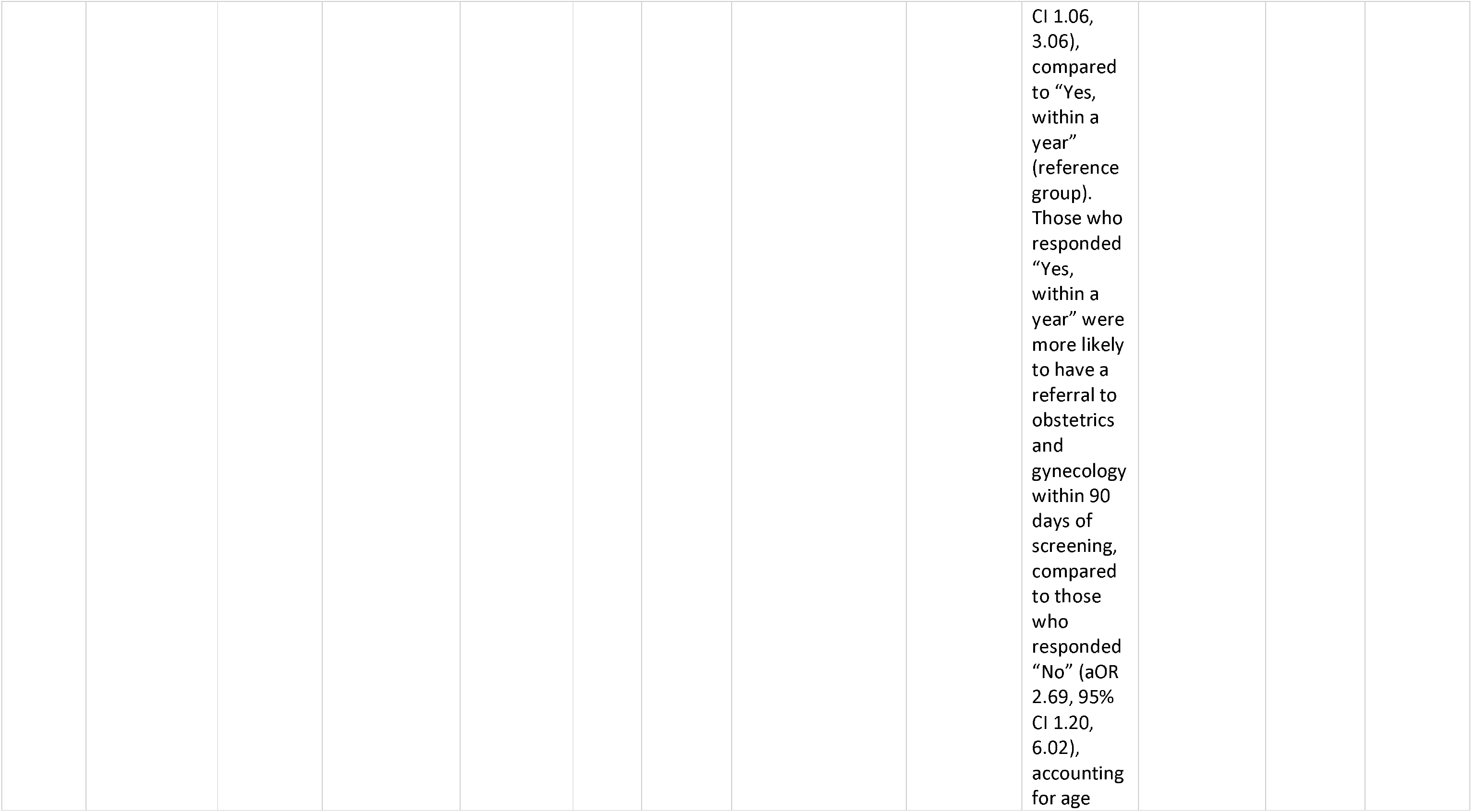
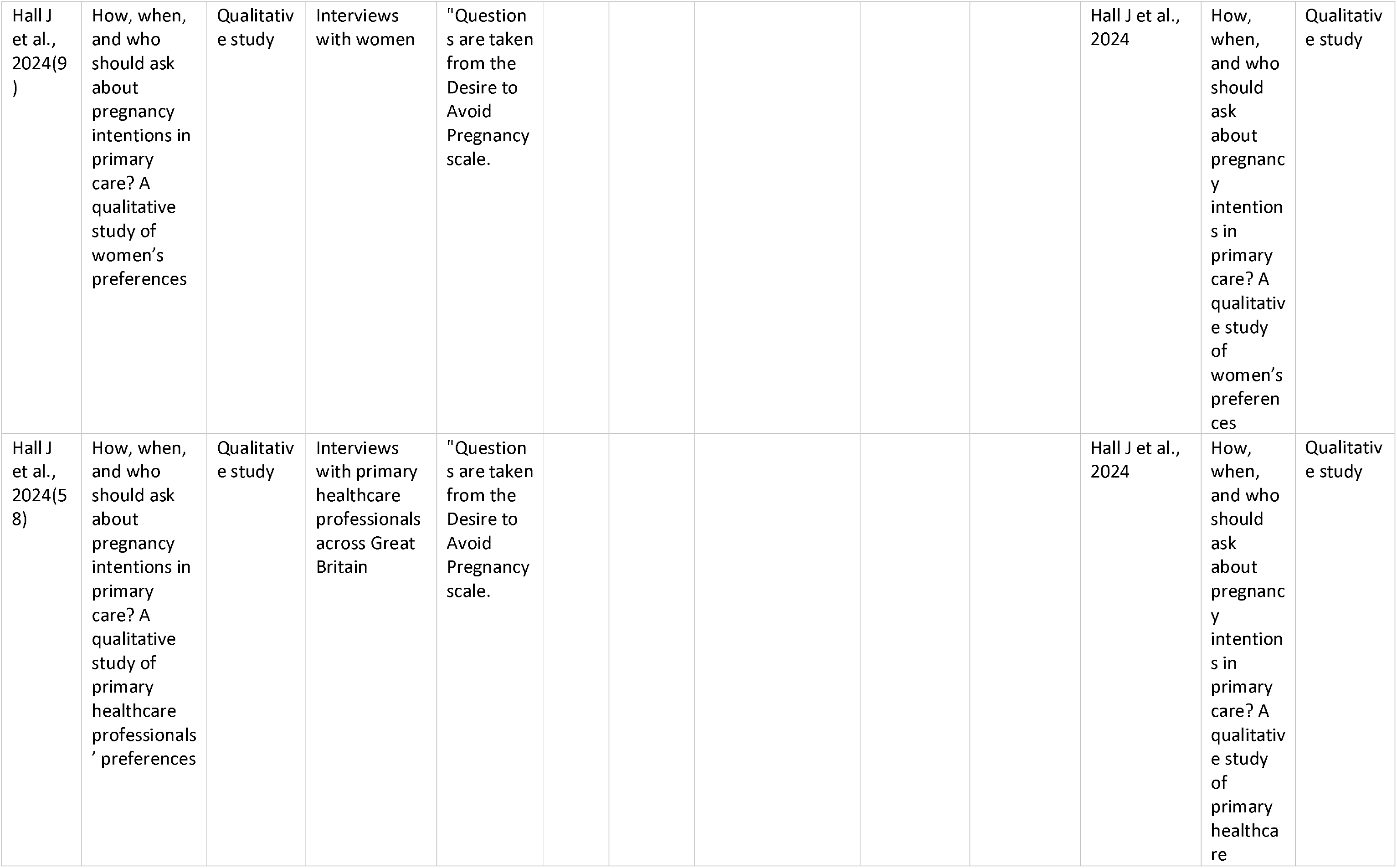
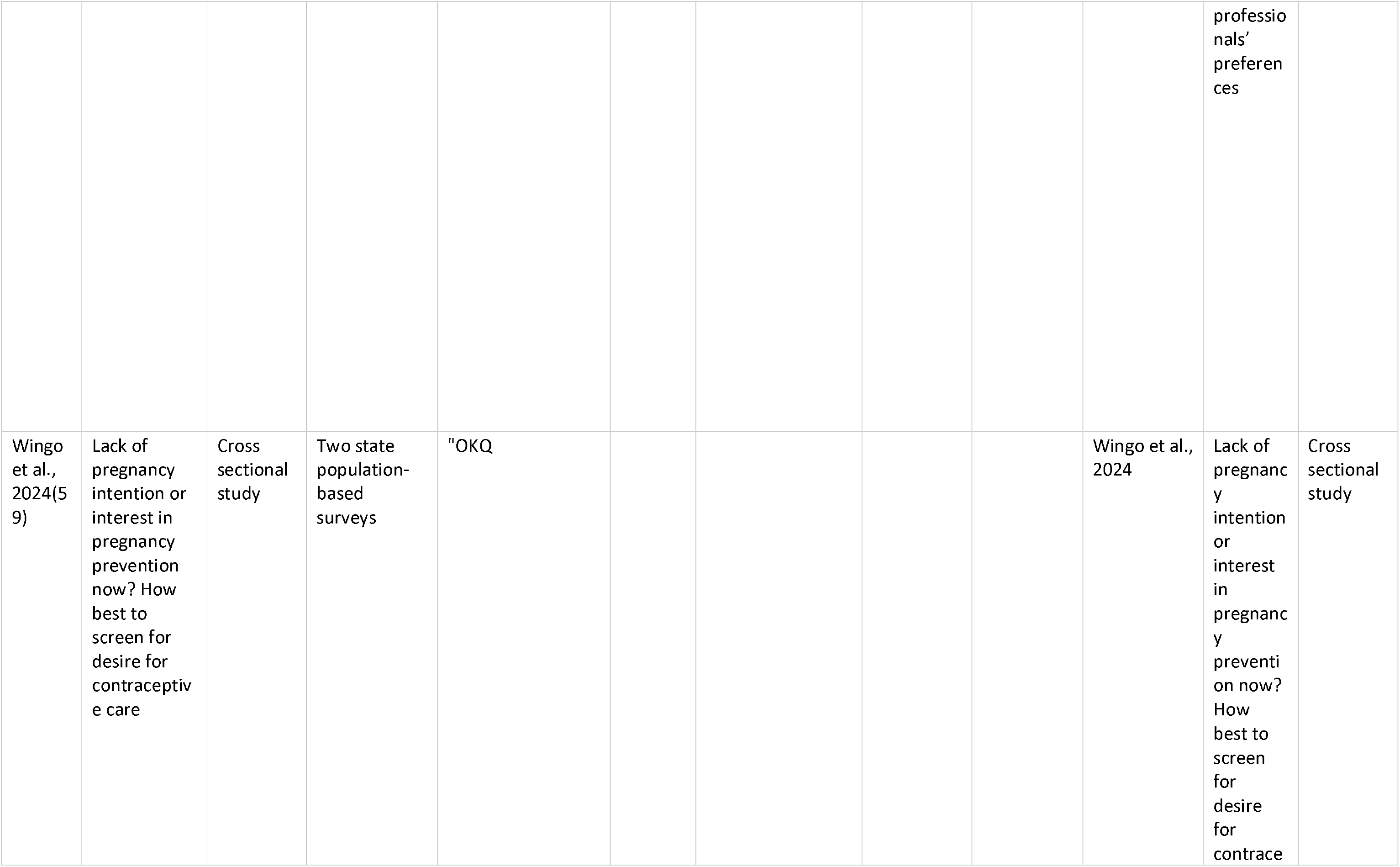
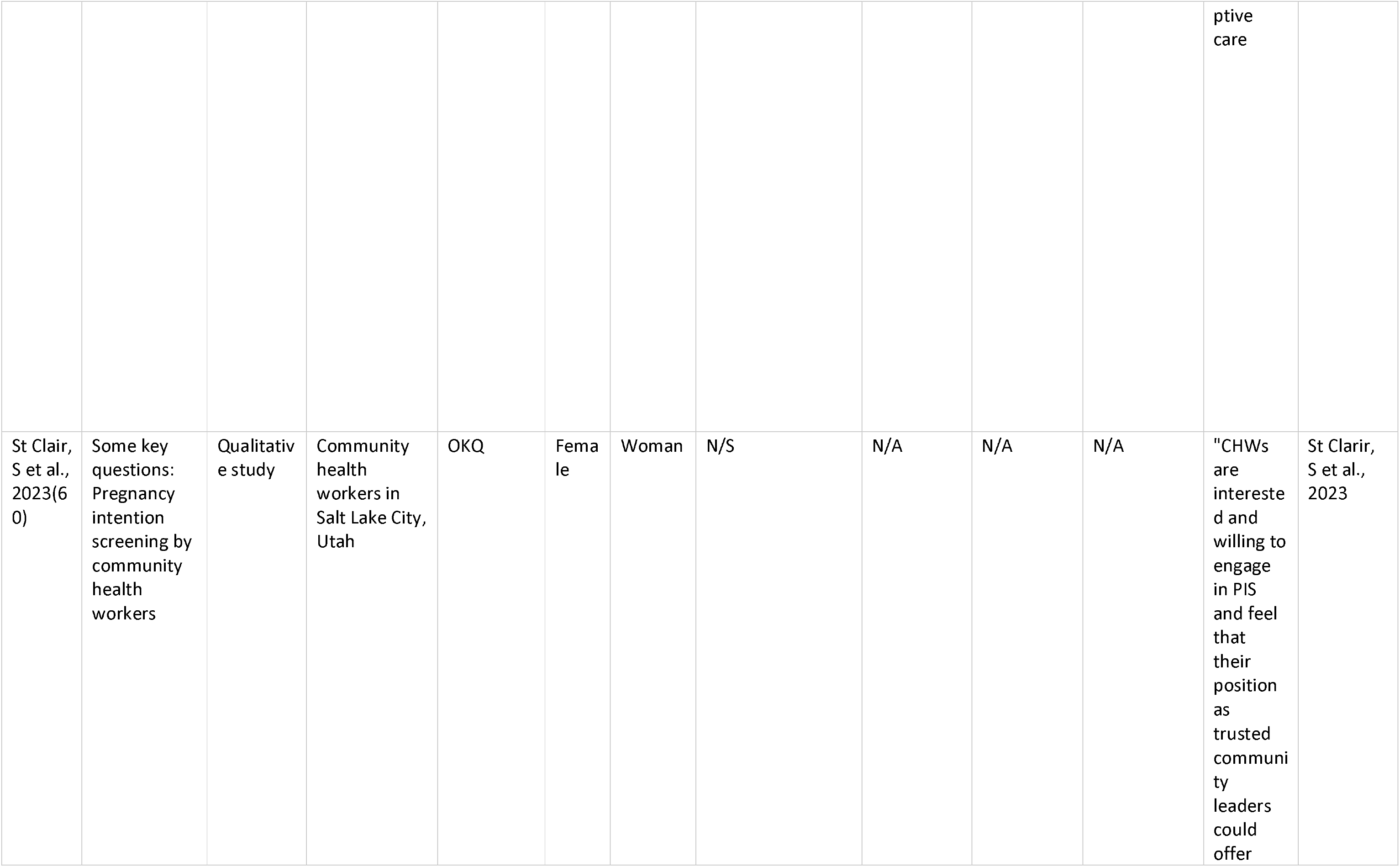
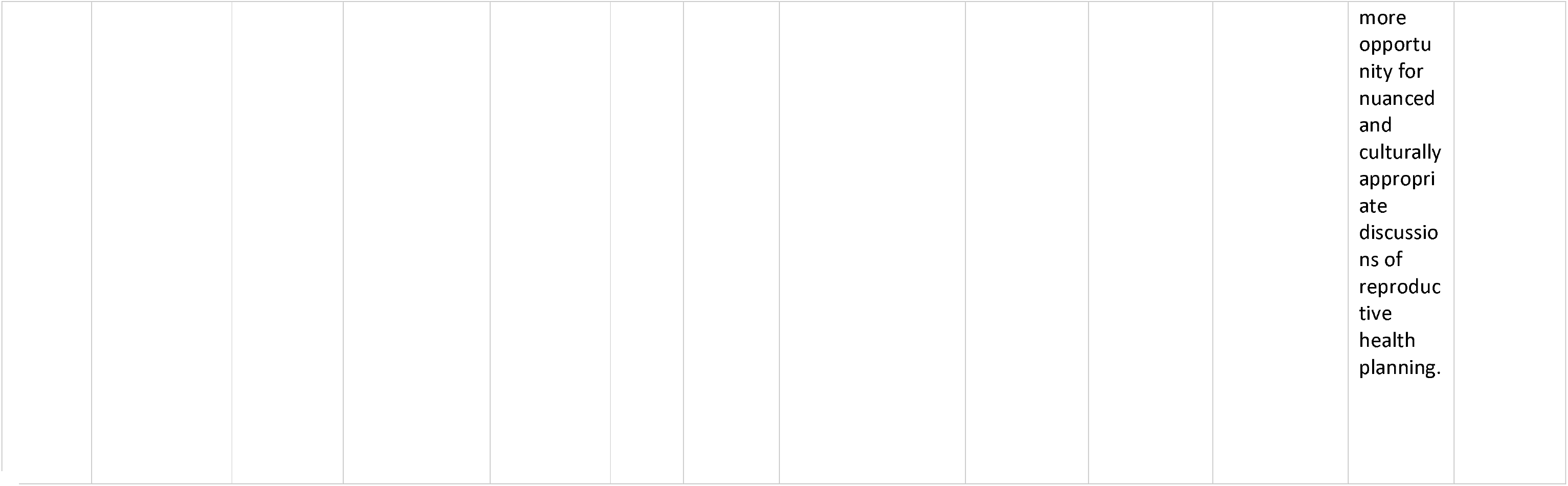
Characteristics of included studies.

**Table 2:**
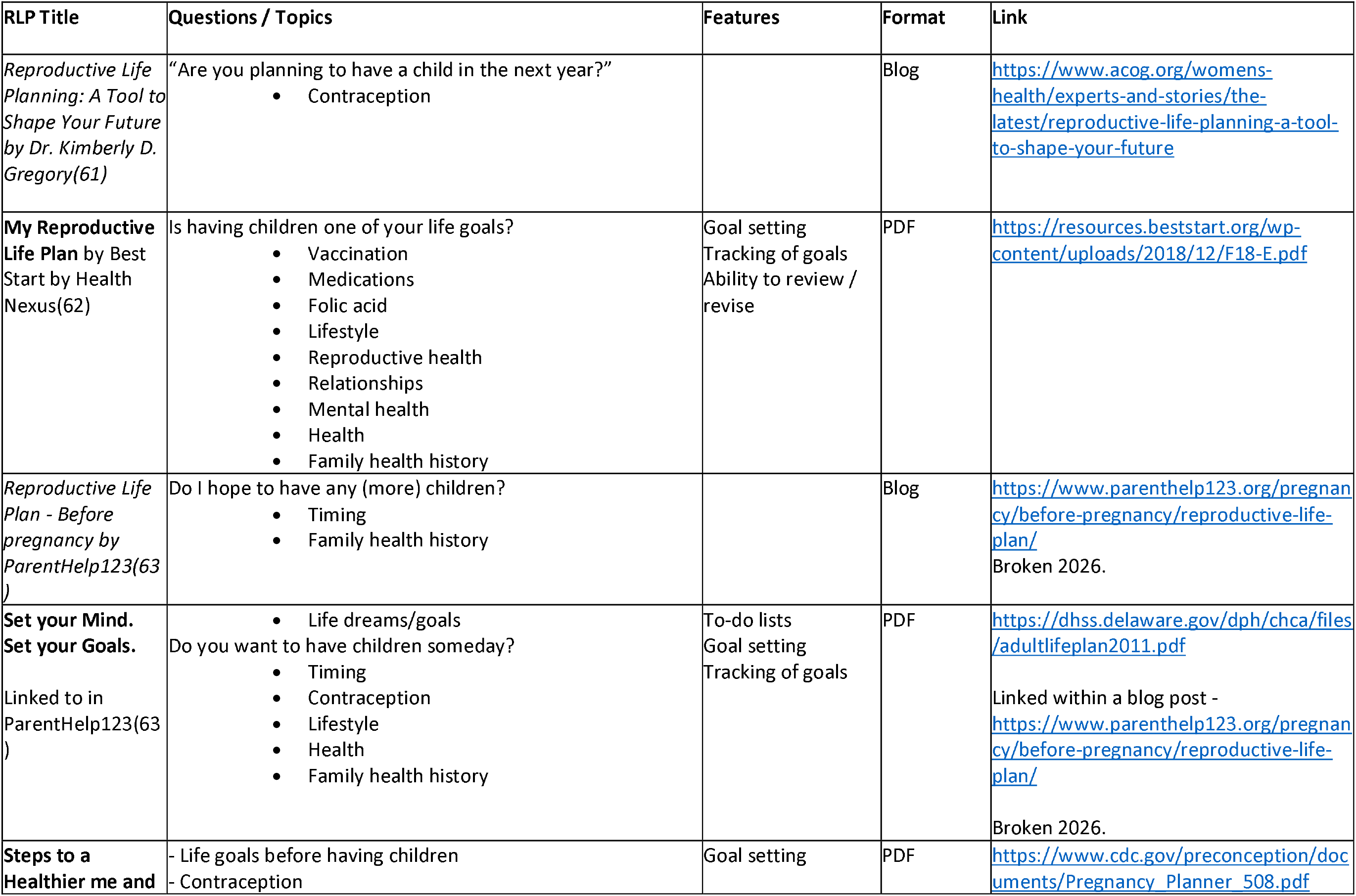
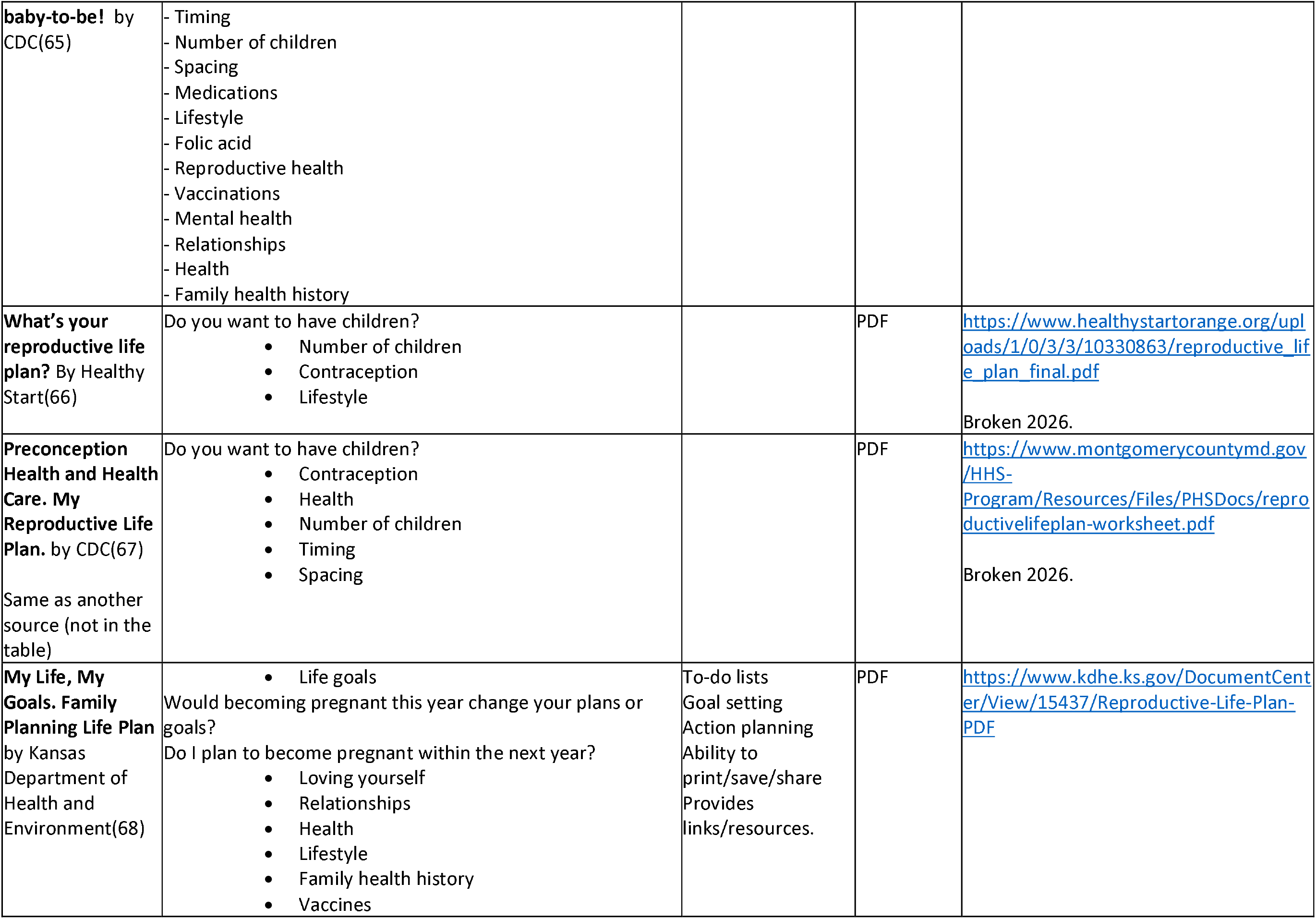
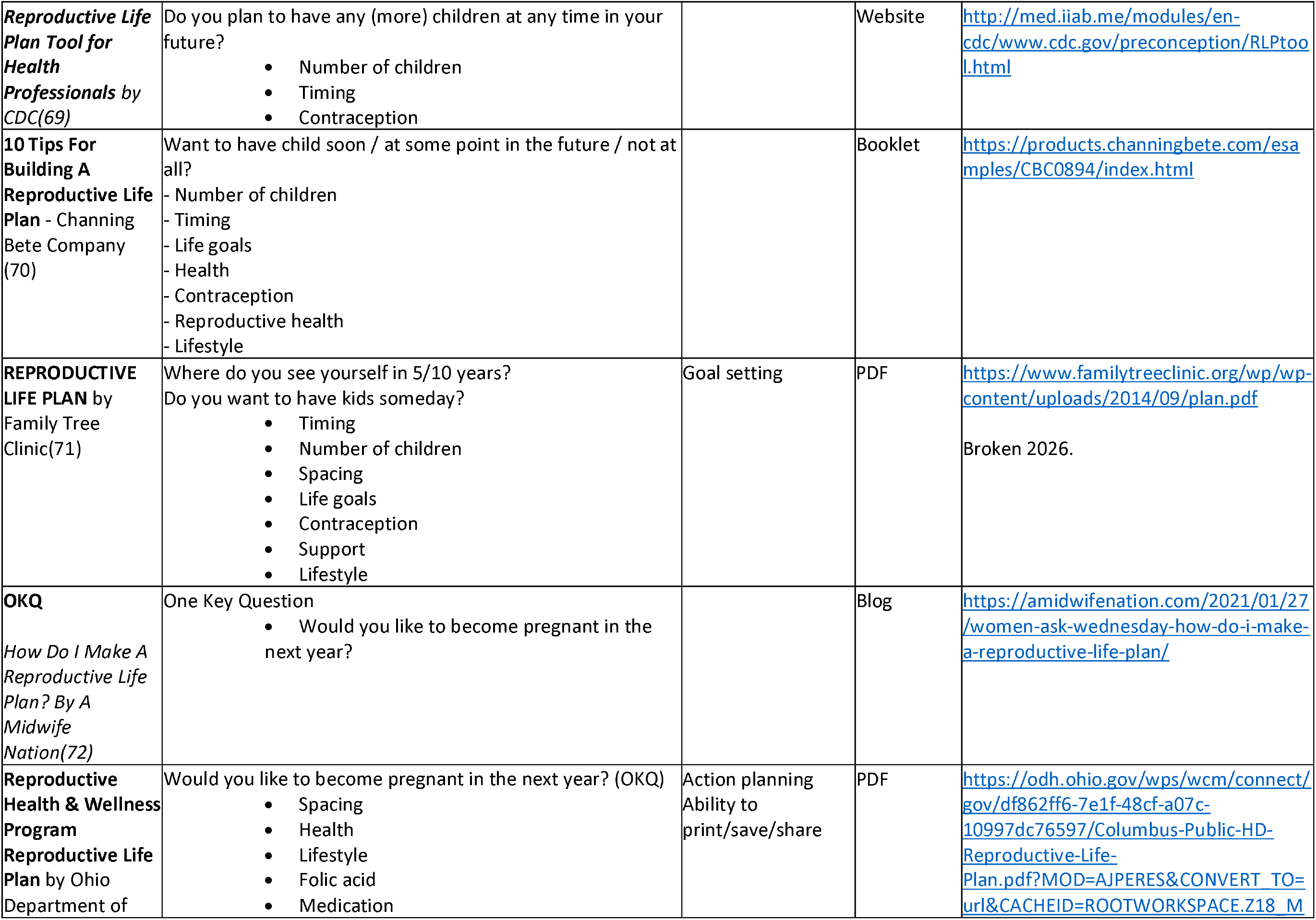
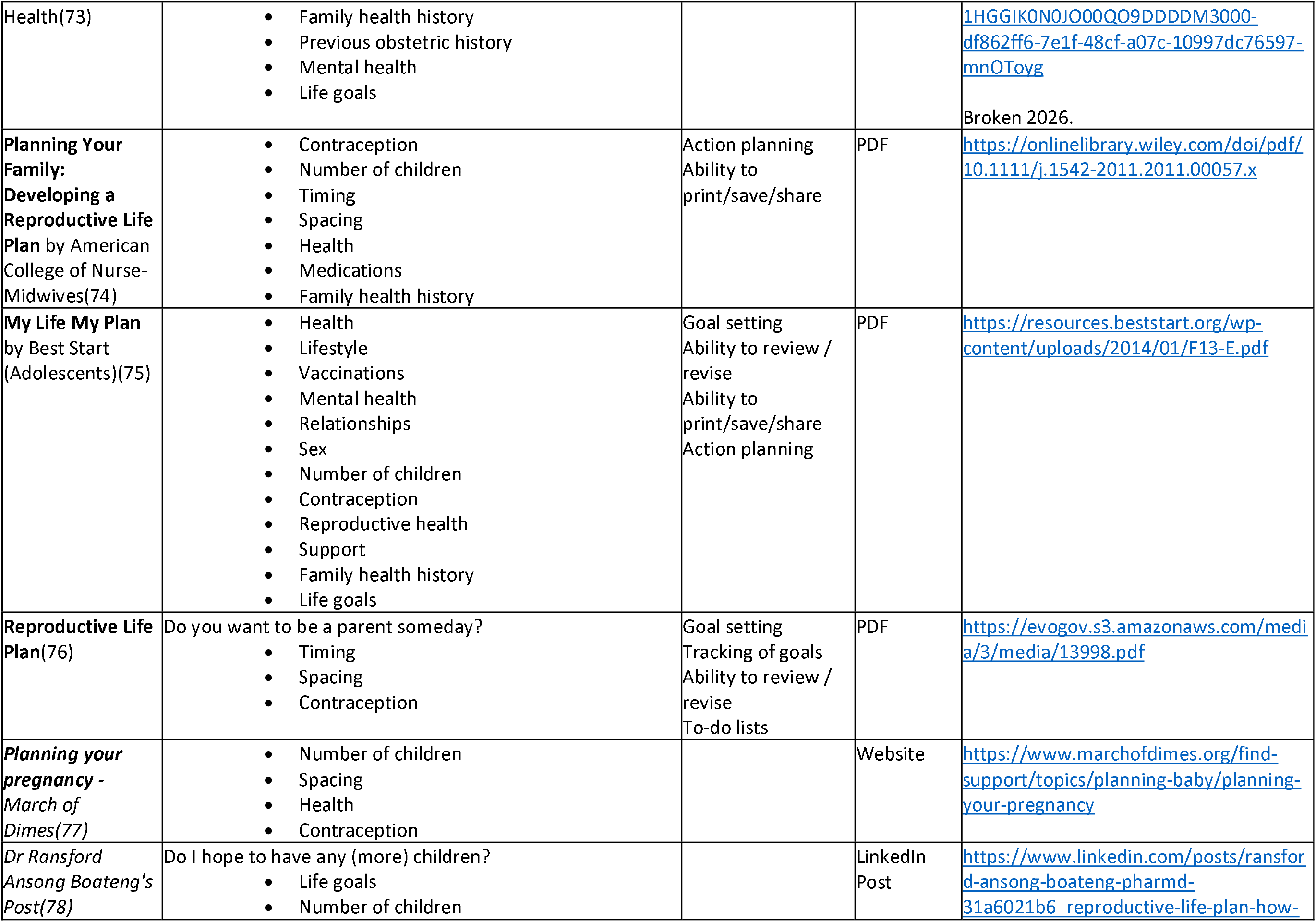
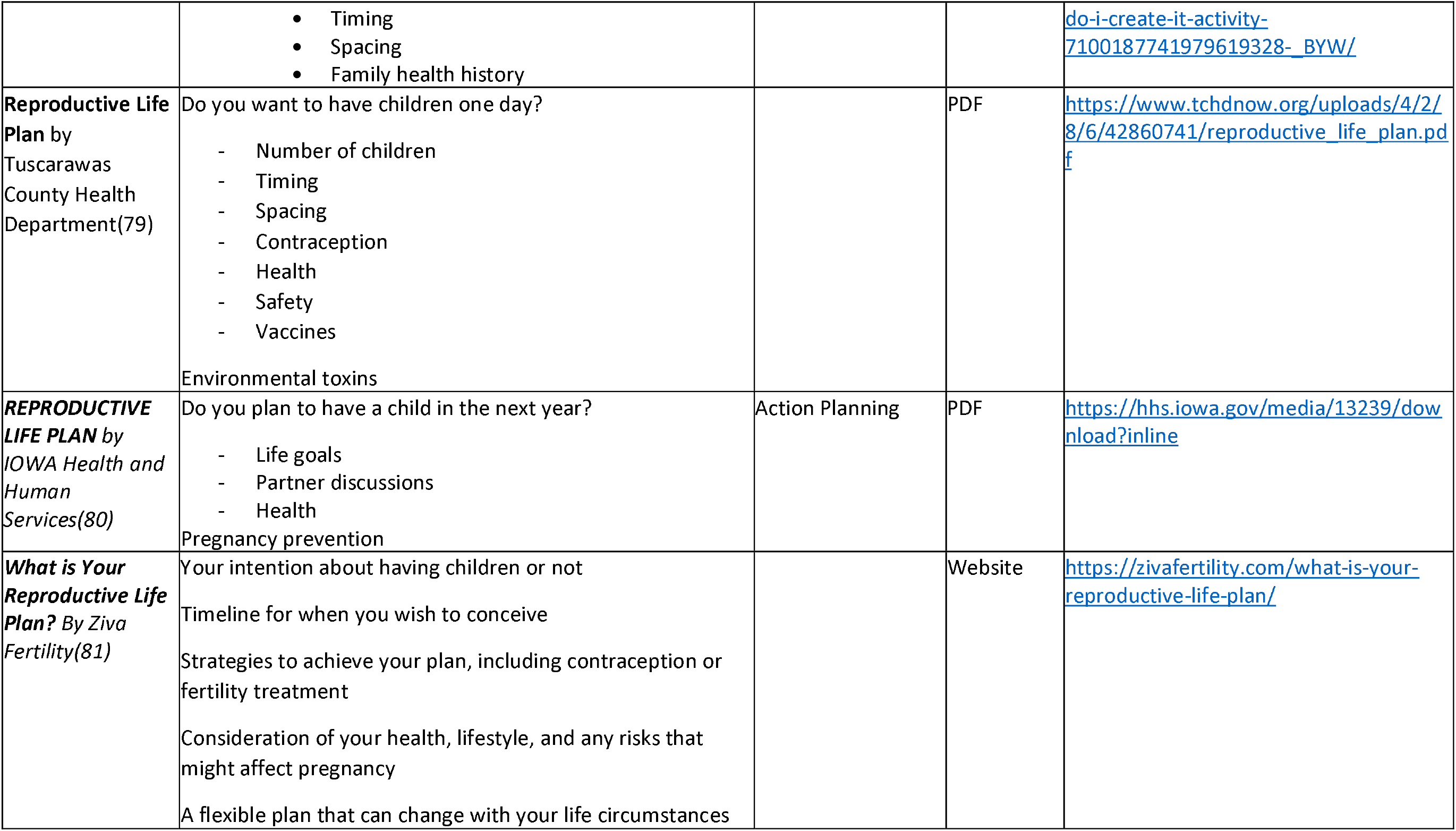
Details of included grey literature sources.

### Findings

We extracted and synthesised the evidence relating to the PIS/RLP’s target audience, format, content including any associated behaviour change theory, features, effectiveness and acceptability of the PIS/RLP. Findings were synthesised within each of these categories with differences between different RLPs, settings and formats considered.

### PIS/RLP

There were 25 different PIS/RLPs discussed in the 51 studies (Table 3).

**Table 3:** Details of Reproductive Life Plans & Pregnancy Intention Screening 1 Questions in Published Literature.

| RLP/PIS name | Questions | Format | Features | Paper | RLP/PIS |
| --- | --- | --- | --- | --- | --- |
| **MyNewOptions** | How many children do I have now?<br>How many more children do I want in the future?<br>Does my partner agree with my decision about if and when I want to have children in the future?<br>What am I doing now to prevent getting pregnant by accident?<br>Relationship, school, job/career Qs. | Digital (self-complete) | Interactive, individualised information, provides assistance, action planning, direct to nearest provider/pharmacists/GP, ability to review/revise and ability to print/share/save | Chuang et al., 2019(19) | RLP |
| **MyFamilyPlan** | Do you plan to have any (or any more) children at any time in your future?<br>Questions covering contraception, sexual health, nutrition, stions, substance use, family/genetic history, environmental exposures, medications, obstetric history, and chronic medical conditions. | Digital (self-complete) | Individualised information, to-do lists | Batra et al., 2018(43) | RLP |
| **MyPath** | Questions are not specified in the paper but are available in the online tool and are based on PATH.<br>Do you currently have any children?<br>Do you think you would like to have (more) children at some point?<br>What are your thoughts about pregnancy today? (options)<br>How important is it to you to prevent pregnancy now?<br>How happy would you feel if you got pregnant now?<br>How upset would you feel if you got pregnant now?<br>Then questions about menstrual cycle, information on health before pregnancy, and finding a method of birth control. | Digital (self-complete)<br>Linked to primary care | Interactive, individualised information, ability to print/save/share | Callegari et al., 2021(51) | RLP |
| **reproduktivlivsplan.se** | Do you wish to have children?<br>NOT SURE<br>- What are your thoughts about children?<br>- What are the advantages of having children? What are the disadvantages of having children?<br>- How do you plan to prevent getting pregnant until you are certain? | Digital (self-complete) | Ability to print/save/share | Ekstrand Ragnar et al., 2018(20) | RLP |
|  | - What parts of the puzzle of life affect your reproductive life plan?<br>YES<br>- Preconception advice<br>NO<br>- Contraception advice |  |  |  |  |
| Moos/CDC | Do you wish/plan to have children/more children at any time in the future/in your life?<br>NO<br>- How do you plan to prevent becoming pregnant (again)?<br>YES<br>How many (more) children do you wish to have?<br>When would you like to become pregnant (again)?<br>If within 0-6 months:<br>How do you prepare for pregnancy?<br>LATER:<br>How do you plan to preserve your fertility?<br>How do you plan to prevent becoming pregnant (again) until then?<br>If maybe or don't know:<br>What are your thoughts on having children?<br>From your point of view, what speaks in favour of or against having children?<br>How do you plan to prevent becoming pregnant (again) until you have decided?<br>What can I do to help you achieve your goals? | Digital (self-complete) with midwife. | Action planning, provides assistance, direct to nearest provider/pharmacists/GP, ability to review/revise, ability to print/save/share | Koo Andersson & Tyden, 2020(26) | RLP |
|  |  | Verbally (clinical) |  | Fooladi et al., 2018(22), Niemeyer Hultstrand et al., 2020(34), Skogsdal et al., 2019(36), Tydén et al., 2016(41) |  |
|  |  | Self-complete (written) |  | Engström et al., 2022(21) |  |
|  | Do you want children in your life?<br>DON'T KNOW<br>-What are your thoughts about children?<br>-What would you do if you became pregnant unintentionally?<br>-How do you plan to prevent becoming pregnant until you are sure?<br>- Which contraception method suits you?<br>YES, SOMETIME IN THE FUTURE, BUT NOT NOW<br>-How many children would you like?<br>-When would you like to have your first child?<br>-When would you like to have your last child? | Verbally (clinical) |  | Stern et al., 2015(39) | RLP |
|  | -Which contraception method suits you?<br>-What other parts of your life affect your RLP?<br>NO<br>-Which contraception method suits you?<br>-How can you protect yourself from STIs?<br>YES, I WANT TO BECOME PREGNANT WITHIN A YEAR<br>-How many children would you like?<br>-When would you like to have your last child? |  |  |  |  |
|  | Desire pregnancy in the future:<br>When do you want to get pregnant?<br>What do you need to change about your health to get ready for pregnancy?<br>Don't desire pregnancy in the future:<br>How will you prevent pregnancy?<br>Is there a possibility of changing your mind?<br>What if you become pregnant by accident? | Verbally<br>(clinical) |  | Callegari et al.,<br>2015(17) | RLP |
| RH-SAT | Want children? Don't want children? Aren't sure?<br>If you want (more) kids now or in the future...<br>-How many (more) kids do you hope to have?<br>-When do you hope to become pregnant or pregnant again?<br>-Information about health before pregnancy and a list of things to talk to your doctor about.<br>-Do you want to wait to get pregnant?<br>IF YOU DON'T WANT (MORE) KIDS RIGHT NOW...<br>-Do you want to wait to have kids later / want no (more) kids / want to learn about birth control but are not sure how you feel about having kids?<br>-What have you tried in the past to prevent getting pregnant (list of options)?<br>-Did you have any problems with any of the birth control methods you used in the past?<br>-What do you want to do to prevent getting pregnant right now?<br>-What help would you like from your doctor?<br>If you are not sure how you feel about having kids...<br>-How would you say you feel about pregnancy right now?<br>-You are ready to be pregnant / you are not ready to be pregnant / | Self-complete<br>(written) |  | Bello et al.,<br>2020(15) | RLP |
|  | you are not sure how you feel about pregnancy right now.<br>-Information about birth control / getting ready for pregnancy and a list of things you could talk to your doctor about.<br>-Are you interested in reading about birth control methods? |  |  |  |  |
| **Reproductive Life Plan Tool**<br>*(modified from CDC)* | As part of your child's care I ask about plans for future pregnancies because the timing of pregnancies has an impact on your child's health and your health. Do you plan to have any more children in the future?<br>NO/UNSURE<br>What are you using for birth control?<br>YES<br>When would you like to become pregnant again?<br>What are you using for birth control?<br>Do you plan to continue this form of birth control until you are ready to become pregnant? | Verbally (clinical) |  | Caskey et al., 2016(44) | RLP |
| **PATH** | Do you think you might like to have (more) children at some point?<br>When do you think that might be?<br>How important is it to you to prevent pregnancy (until then)? | Verbally (clinical) |  | Callegari et al., 2017(6), Goodsmith et al., 2023(25) | RLP |
| **Family Planning Quotient (FPQ) and Reproductive Life Index (RepLI) (FPQ/RepLI)** | FPQ<br>How many children do you have? (including adoptive/step)<br>How many children do you desire?<br>OKQ<br>Would you like to become pregnant in the next year?<br>RepLI<br>Asks questions about other pregnancy outcomes including whether live births were intended or unintended, adopted or step-children, miscarriages, ectopic or tubal pregnancies, elective abortions, stillbirths, and child deaths. The tool also incorporates other reproductive health indicators including menarche, sexual debut, contraceptive use, and history of sexually transmitted infections (STIs). *(Cancer diagnosis/treatment is also included, as FPQ/RepLI has been adapted for use with oncology)* | Verbally (clinical) |  | Madrigal et al., 2019a(29), Madrigal et al., 2019b(30) | FPQ - RLP<br>OKQ- PIS |
|  |  | Self-complete (written) |  | Baldwin et al., 2018(50) |  |
| **Healthy Start RLP** | Do you want to have (more) children one day?<br>YES:<br>What age would you like to have children? | Self-complete (written) – linked to |  | DiPietro Mager et al., 2018(14) | RLP |
|  | How many children would you like to have?<br>How far apart would you like your children to be?<br>NO/UNSURE:<br>Are you currently pregnant?<br>What is your plan to prevent pregnancy?<br>What will you do if you become pregnant?<br>Are you currently using a birth control method?<br>If yes, what method are you currently using?<br>If pregnant, method before pregnancy?<br>Personal Habits - tobacco use, alcohol, drugs.<br>Physical Activity and Nutrition - weight/Height/BMI, physically activity, diet.<br>Emotional Health - sad, nervous, anxious, worried, anger, physical/sexual abuse.<br>Reproductive Health - STIs, pregnancies/children, abortions/ miscarriages / stillbirth, preterm / low birth weight babies.<br>Health Problems<br>Current Medication<br>Vaccinations<br>Family History<br>Personal and Professional Goals<br>List of counselling and Referrals Provided | clinical encounter |  |  |  |
| No name | Do you have any children now?<br>Do you want to have (more) children?<br>How many (more) children would you like to have and when? | Verbally (clinical) |  | Nelson et al., 2016(33) | RLP |
| Health Assessment Questionnaire (HAQ) | (1) Are you sexually active? (2) Would you or your partner like to become pregnant in the next year? and (3) Which birth control are you using? | Self-complete (written) |  | Moon, B. et al (2025)(55) | RLP |
| No name | Are you pregnant?<br>Are you planning a pregnancy?<br>Are you using contraception?<br>What type of contraception are you using? | Verbally (clinical) |  | Murnion, B et al (2025)(56) | RLP |
| One Key Question (OKQ) | Would you like to become pregnant in the next year?<br>- Yes, No, I'm not sure, OK either way | Verbally (clinical) |  | Allen et al., 2017(42), Thorman et al., 2022(45), | PIS |
|  |  |  |  | Fitch et al., 2023(53), Goodsmith et al., 2023(25), Hipp et al., 2017(24), Kvach et al., 2017(28), Manze et al., 2020a(31), Manze et al., 2020b(32), Pryor et al., 2022(35), Stulberg et al., 2019(40), Villafana et al., 2022(52), St Clair et al., 2023(60). |  |
|  |  | Self-complete (written) |  | Baldwin et al., 2018(50), Ferketa et al., 2022(47), Gawron et al., 2021(46), Song et al., 2021(37), Wingo et al., 2024(59) |  |
| No name | Do you wish to have children? | Verbally (clinical) |  | Bodin et al., 2018(16) | PIS |
| No name | What are your thoughts on you (*or your partner*) becoming pregnant? | Verbally (clinical) |  | Jones et al., 2020(49), Manze et al., 2020(31) | PIS |
| No name | Would you or your partner(s) like to become pregnant in the next year? | Verbally (clinical) |  | Jones et al., 2020(49) | PIS |
| No name | How soon after this baby are you planning to become pregnant? Within the next year? | Verbally (clinical) |  | Villafana et al., 2022(52) | PIS |
| No name | Do you think you would like to become pregnant in the future? | Verbally (clinical) |  | Janiak, E et al (2024)(57) | PIS |
| **Desire to Avoid Pregnancy Scale**<br>*(modified single question)* | Using a five-point Likert scale (ranging from 'strongly disagree' to 'strongly agree' how much would you agree or disagree that:<br>It would be a good thing for you if you became pregnant in the next 3 months<br>It would be the end of the world for you to have a baby in the next year<br>You want to have a baby within the next year<br>Thinking about becoming pregnant in the next 3 months makes you feel excited<br>You would worry that having a baby in the next year would make it harder for you to achieve other things in your life | Verbally (clinical) |  | Hall J et al., 2024a,(9) Hall J et al., 2024b(58) | PIS |
| No name | Many of my patients are thinking about either getting pregnant or preventing a pregnancy. Where are you on this issue right now? | Verbally (clinical) |  | Jones et al., 2020(49), Manze et al., 2020a(31), Manze et al., 2020b(32) | PIS/Services Assessment |
| No name | Can I help you with any reproductive health services today, such as birth control or planning for a healthy pregnancy? | Verbally (clinical) |  | Jones et al., 2020 (49), Manze et al., 2020a (31), Manze at al., 2020b (32), Srinivasulu et al., 2022 (38) | PIS/Services Assessment |
| No name | Would you like your provider to help you with birth control or pregnancy planning today? | Verbally (clinical) |  | Shah et al., 2019(48), Srinivasulu et al., 2020(54) | PIS/Services Assessment |

Some PIS/RLPs were mentioned in multiple papers, e.g. One Key Question (OKQ) (n=17); CDC/Moos based RLPs (n=8); FPQ/RepLI (n=3); PATH (n=2) and Desire to Avoid Pregnancy (DAP) Scale (n=2), although they were not always asked in the same format.

The 21 grey literature sources discuss 20 different RLPs, though there were lots of commonalities and many were based on the CDC/Moos RLP(7).

### Target audience

In the published literature, the target audience for 46 of the 51 papers was cisgender females of reproductive age, although the age range varied. Tools used in these studies were specifically designed for individuals capable of becoming pregnant. Four papers were aimed at everyone(20, 26, 27, 49), with only one paper specifically focused on men(16). One study focused on women with cancer(29), one on women with chronic mental illness(25), one on women with Rheumatic Disease (55) and one on women receiving treatment for alcohol or other drug use (56), but most did not mention specific medical conditions.

In the grey literature, the target population was often unclear, however, 11 appeared to be targeting females and 10 appeared to be for everyone. Age was not specified in most cases, but two were specifically focused on adolescents/teens(71, 75) and another had an adolescent section(79).

### Format

The majority of PIS/RLPs were employed verbally during clinical encounters (18 RLPs in 39 papers)(5, 6, 16, 17, 22, 24, 25, 28–32, 34–36, 38–42, 44, 45, 48, 49, 52–54), six were self-completed (written) RLPs studied in eight papers(14, 15, 21, 37, 46, 50, 55) and six were digital RLPs(13, 19, 20, 26, 43, 51), although one was designed to be used/completed with a midwife(26) and one was linked to primary care(51). Several PIS/RLP were implemented in different formats, for example, the OKQ, FPQ/RepLI, DAP and Moos based RLPs were all asked (or could be asked) both verbally during clinical encounters and self-completed (written). All PIS (apart from OKQ) were only used in clinical encounters, while the RLPs were used in clinical encounters, self-completed and digital. While the method of recording PIS/RLP was not stated for most of the papers, the electronic medical record was the most frequently identified location (n=12).

Most of the RLPs in the grey literature were designed to be self-completed, however three were aimed to be used by clinicians within clinical encounters (69, 79, 80). 13 of the 217 RLPs in the grey literature were PDFs, seven were within blogs/websites and one was a booklet available to purchase (70).

### Content – questions

Most of the RLPs, whether in the published or the grey literature, started with a question like the options shown in Box 3. For those who answer yes to having children, most RLPs go on to ask how many children an individual would like, when they would like to have those children, and what method of contraception will be used to prevent pregnancy until the desired time. Some of the longer RLPs also asked about/mentioned preconception health e.g. asking about health behaviours, health problems or medications, however this was rare for RLPs in the published literature (n=6)(14, 19, 20, 43, 51, 57). For those that answer no, most RLPs ask how an individual will prevent pregnancy. For those who are unsure, most RLPs ask individuals what their thoughts on having children/getting pregnant are and how they will prevent pregnancy until they have decided what they want. Details of the specific topics covered by each RLP, the format of the RLP and its features can be found in Table 3.

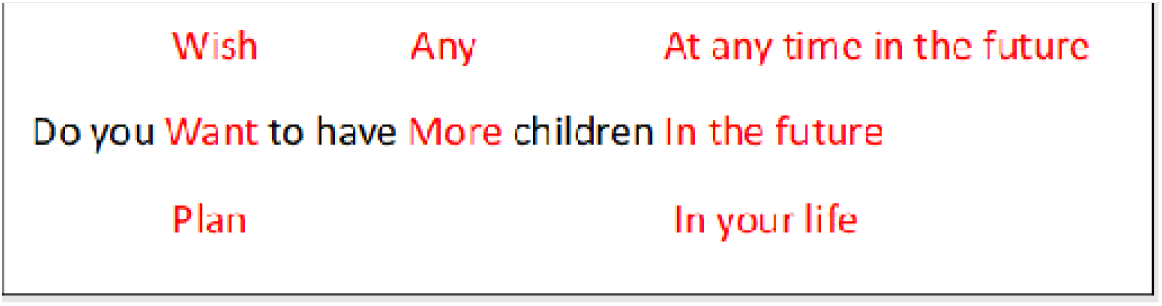

Box 3: Word tree of the most common wording of opening questions in RLPs

The RLPs in the grey literature tended to be longer, e.g. My Life My Plan was 12 pages long(75). This was because they usually also asked about other life goals, such as education and career aspirations, lifestyle, health (mental and physical) and family health history, relationships (e.g. partner’s thoughts, support, abuse) as well as questions about wider reproductive health (e.g. sexually transmitted infections) and previous obstetric experiences.

The PIS questions tended to ask an individual about their thoughts/feelings about becoming pregnant, rather than ‘having children’ and focused on a shorter time frame such as a year. Some had answer options, for example the OKQ has ‘Yes’, ‘No’, ‘I’m not sure’, ‘OK, either way’, but most were open-ended in line with the verbal format. Some questions were focused on the reproductive health services an individual wanted and were therefore only applicable to clinical encounters(31, 32, 38, 48, 49, 54). The exception to this was the DAP (9, 58), a 14-item research tool assessing the strength of an individual’s desire to avoid pregnancy. While a robust research tool its current length makes it impractical for routine implementation; a shorter version is understood to be under development offering the promise of a more valid way of screening for pregnancy intentions.

### Content – topics covered

Thirty-four papers discussed information/advice that was provided alongside the RLP; most provided information about health behaviours, lifestyle, fertility and contraception, with folic acid supplementation mentioned in both published and grey literature. For those that were used verbally during clinical encounters (especially the PIS) some stated the information/advice that should go alongside the RLP/PIS but it is impossible to know what discussions individual healthcare professionals actually had with their patients. From the grey literature, ‘My Life My Plan’, which is specifically targeted at teenagers, included topics relevant to this demographic, such as being ready for sex, relationships and teen pregnancy, as well as providing information (and links) specific to teenagers(75), while the Tuscarawas County Health Department Reproductive Life Plan also included a section for adolescents that included information about sexual coercion and open communication(79).

### Features

Five of the digital RLPs included features such as being interactive(19, 51), providing individualized information(19, 43, 51), creating to-do lists(43), and allowed the RLP to be revised(19, 26) or printed/saved/shared (19, 20, 26, 51)(19, 20, 26, 51). Other features included providing assistance such as directions to nearest provider/pharmacy/GP, or incorporated action planning into the RLP.

Of the 12 RLPs that were PDFs in the grey literature, nine had features including goal setting(62, 64, 65, 68, 71, 75, 76), tracking goals(62, 64, 76), to-do lists(64, 68, 76), action planning(68, 73–75, 80), ability to review/revise(62, 75, 76) or print/save/share(68, 73–75).

### Behaviour Change

Of the 51 included papers, 35 made no mention of any underpinning behaviour change theory and there was no explicit mention in the grey literature of behaviour change theory. Studies focused on implementation in a clinical setting often mentioned the importance of taking a patient-centred approach, ensuring shared decision-making and balanced counselling, and aspects of motivational interviewing, which is a recognised behaviour change technique. The web-based ‘My New Options’ RLP incorporated action planning(19), which encourages a person to think in advance how to implement the desired behaviour and how to address challenges, reducing the cognitive load when these situations are faced(82).

The development of three RLPs, all of which were digital, was said to be informed by specific behaviour change theories: Fishbein’s reasoned action approach to health promotion(43); the health promotion model(13); and self-determination theory(51, 83).

Batra et al(43) hypothesised that their RLP ‘MyFamilyPlan’ would increase preconception discussion with HCP via an increase in self-efficacy, a component of Fishbein’s theory(84). The intention was that the pre-consultation use of ‘MyFamilyPlan’, which provided actionable, specific preconception health recommendations and encouragement to discuss them with a provider, would empower patients to initiate discussions of reproductive health.

The health promotion model was utilised by Jagoda et al(13) as their conceptual framework. This model concentrates on three major categories: individual characteristics and experiences, behaviour-specific cognitions and affect; and the behavioural outcomes(85). It has previously been applied to reproductive health and showed that social support, perceived benefits and perceived barriers were the most important predictors of health promoting behaviour(86).

Finally, Callegari et al(51) implemented self-determination theory, a personality theory that focuses on the motivations behind decisions, in their web-based RLP ‘MyPath’(87). It focuses on intrinsic motivation i.e. engagement in the activity is valued rather than valuing the outcome. This theory suggests that there will be improved health behaviours and outcomes where health care meets individuals’ psychological needs for autonomy, competence, and relatedness (feeling understood and cared for by others). ‘MyPath’ was designed to build competence through knowledge and self-efficacy, autonomy by encouraging reflection on reproductive desires and goals, and relatedness by promoting patient-centred communication by clinicians.

### Effectiveness

28 out of 51 studies (54.9%) measured effectiveness as a study outcome, however there were several different ways that effectiveness was conceptualised. The most common effectiveness outcome was contraceptive use (14/28, 50.0%). One study described a significant increase in long-acting reversible contraceptive (LARC) use, but not pill or injection, following RLP implementation whilst controlling for ethnicity and education (aOR 1.635, 1.03, 2.61)(12), and another, using the MyNewOptions tool, noted no differences in contraception use between intervention and control arms(19). The remaining studies described either stable contraception use or did not report contraception use pre-intervention as a comparator.

Most of the nine studies(35, 45, 46, 48, 52, 54, 55, 57) reporting documentation of contraception or preconception counselling measured a significant increase after implementation of the RLP with intervention arm rates ranging from 58.4% in a study of the OKQ(54) to a maximum of 93.1% in a study of the ‘MyPath’ tool(51). Reproductive health knowledge also appeared to increase across the seven studies measuring this outcome. One study of the OKQ reported 77% of women had received new information and fertility awareness increased from a mean score of 4.6 to 5.5 out of 12 (p=0.004)(16).

Medication reviews and use of folic acid were less commonly measured outcomes of effectiveness, reported in only three out of 51 studies (5.9%). Results of the impact of the RLP on folic acid use were inconsistent across two studies: a study of the Moos/CDC tool showed a statistically significant increase in folic acid use prior to pregnancy after RLP implementation(22) whilst a study using the ‘MyFamilyPlan’ tool showed no difference between intervention and control(43). ‘The Healthy Start’ tool, implemented as part of a program serving urban low-income African-American women, was found to result in a medication review in 75% (437/580) of cases of which 8% (35/437) were identified as including potentially teratogenic medications(14).

Five of the six studies using digital tools evaluated effectiveness(13, 19, 26, 43, 51). Digital RLPs were shown consistently to increase reflection(13), discussion and advice provision(26, 43), and to sometimes increase self-efficacy(51) but there was limited evidence of the translation of this into contraception or preconception behaviour change(19, 43, 51). A cluster randomised controlled trial of 292 women, showed that participants completing the ‘MyFamilyPlan’ health education module prior to a well-woman visit were significantly more likely to report discussing reproductive health with their physicians (OR 1.97, 95%CI 1.22-3.19), though there was no impact on folate use before pregnancy(43). This tool was based on Fishbein’s reasoned action approach and aimed to improve self-efficacy to improve these outcomes, though they did not show an improvement in self-efficacy as measured in the study. Self-efficacy and knowledge were measured in the study of’ MyPath’ demonstrating a greater increase in both domains in the intervention arm versus the control arm, as well a significantly increased discussion of pregnancy and/or contraceptive needs (93.1% vs 67.9%, p=0.02)(51). There were non-significant increases in underpowered secondary outcomes of participants reporting the highest confidence that their contraceptive method was right for them the intervention versus control group (33.3% versus 7.4%, p = 0.11) and in the proportion of participants reporting the lowest level of decision conflict about their contraceptive decision (23.3% versus 7.1%, p=0.09).

### Acceptability to users

Acceptability to users was assessed in 28 studies, from the USA (n=19), Sweden (n=4), Australia (n=2)(52, 56) and one each in Iran(22), Eswatini(21) and UK(9). Users were overwhelmingly positive about being asked about reproductive life planning across countries, settings and tools, commonly appreciating the opportunity to discuss their reproductive health and stating that PIS/RLPs should be routine. One study that looked at men only(16) showed that most (76%) found being counselled about their RLP in sexual health clinics in Sweden to be a positive experience. Negative reactions were extremely rare: 3.1% of respondents in another Swedish study found the experience ‘fairly negative’ and none thought it was very negative(36); in the well-baby setting only three out of 55 declined to discuss the issues raised by the RLP (including one who had a tubal ligation)(43); and one participant said that mental health professionals should not ask RLP questions(25). In the UK study, women found it acceptable to be asked about their pregnancy preferences in in a range of primary care settings(9).

Qualitative findings showed that RLPs were a valuable prompt for women to think critically about pregnancy intentions and related actions(9, 17, 23) as they were hesitant to raise the topic themselves(15, 17, 23) and therefore preferred providers to raise the issue(17, 23). However, it was vital that providers did this, and gave the subsequent advice, in a non-judgemental, unbiased way that respected autonomy(17, 23, 32). This was particularly important for those with lower agency due to perceived judgement, for example where their desires may not conform with society’s expectations of who should become a mother and when, whether that related to teenage pregnancy(23) or being childless by choice(38).

The four studies of digital RLPs’ acceptability(19, 20, 43, 51), found them to be an acceptable format, including in one study that included men(20). Respondents described an interactive, web-based format as easy to use and ‘desirable’(19) with the majority reporting that they liked the online format (e.g. 75.3% in (44)). Comments related to specific tools included ‘user friendly’ with ‘interesting content’(20). High levels of completion (97.9% of intervention participants completely reviewed ‘MyFamilyPlan’(43)), recommendation (e.g. 93.3% would recommend ‘MyPath’ to other women Veterans(51)), and retention (follow up rates over two years of 94-95%(19)) are further evidence of acceptability. For this format it was important that it was seen as a trustworthy and reliable source(20), that struck the right balance between brevity, ease of understanding and comprehensive content, for example by having ‘click here to read more’ options(20, 43, 51) and that was inclusive i.e. not heteronormative(20).

Across different settings and populations, the FPQ/RepLI(29, 30), RH-SAT(15), PATH(25), FPQ(50) and OKQ(37, 50) were all shown to help participants talk to their provider about their reproductive goals, or to lead to an overall increase in satisfaction (from 81% to 97%, p=0.04) and communication (from 77% to 94%, p=0.06)(37). Where comparisons were made between tools, PATH(25), FPQ(50) and questions about reproductive service needs(32, 49) were preferred to OKQ.

### Acceptability to HCPs

HCP opinions were assessed in 18 studies, including nursing professionals (n=3), midwives (n=5), primary care healthcare professionals (n=9), obstetric and gynaecology doctors (n=2), psychiatrists (n=1), paediatricians (n=1) or support staff (e.g medical assistants, medical students, or administrative staff) (n=3), or a mentor mother (n=1). Data came from Sweden (n=4), the USA (n=11), Eswatini (n=1), Australia (n=1) and the UK (n=1). Overall, across the countries, professionals, formats and tools there was general support for the implementation of PIS/RLP as a good way of raising an important topic, that was generally a positive experience for the HCP(20, 26, 34, 39). HCP’s views on two online tools were positive(20, 26, 51), with most HCPs agreeing that women using MyPath prior to clinic attendance had helped users make informed decisions about pregnancy planning/timing (71.4%) and contraception (100%). There were no specific concerns with any of the approaches used, though more recently there has been growing evidence that, in the context of a clinical encounter, a question asking about service needs may be more acceptable to HCPs(30, 48), while another study found that HCPs favoured asking more open questions, such as how the person would feel about a pregnancy in the next year(58). Lack of training (30, 58) or time during consultations(39, 45, 47, 58) were raised, though some felt it was time-efficient(39, 46, 51) and while empirical evidence suggested an additional 1-5mins was the norm(44, 48, 53), even this could be unmanageable in a busy clinic. In relation to this, several studies showed support for self-complete options(26, 46, 51).

Ten papers considered the acceptability of the OKQ, either alone or in comparison with other RLPs: six were as part of a clinical consultation(30, 31, 45, 52, 53, 60); four were in a self-complete format(46, 47, 50); and all but one(52) were in primary care. Where OKQ was considered alone(45–47, 52, 53), providers generally found it easy to use, praising its simplicity(60), especially when related to the patient’s reason for presenting(53) and a helpful reminder to consider reproductive goals(45, 46) and get the conversation started(52, 53), though some felt overwhelmed by existing clinical reminders(46). However, when compared with other tools, Madrigal et al (30) found OKQ was the least-preferred option in comparison with FPQ/RepLI and a screening question for reproductive service needs (which was the preferred option). Wingo et al (59) found that OKQ did not capture the current pregnancy prevention desires of a sizeable minority of respondents. A screening question focused on service needs was highly acceptable to support staff in primary care in another USA study(48). Conversely, in Baldwin et al’s comparison of OKQ and FPQ(50), fewer providers agreed the FPQ tool helped to focus their counselling, though this was not statistically significant (FPQ 16/43, 37.2% versus OKQ 18/36, 50%; p=0.25).

### Awareness

Three studies considered awareness of RLPs: one each in Australia(18), the USA(27) and Iran(22). Most people were unaware of RLPs, whether or not they were currently seeking a pregnancy. A survey of women in Australia, for example, found 90.2% were unaware of RLP, with no statistically significant difference between ‘active planners’ (desire pregnancy in the next year) and ‘non-active planners’ (1-5 years)(18). There was slightly higher awareness (24%) amongst college students in the USA, but only 4.2% reported being asked about RLP by an HCP(27). Despite low awareness, most (62.9%) agreed developing an RLP was important and 68.7% wanted to receive information about RLP before attempting pregnancy, from either primary care or an obstetrician/gynaecologist. Awareness of RLPs amongst midwives in Sweden increased from 68% in 2012(41) to 96.2% in 2018(26).

## Discussion

### Findings and Interpretation

Our review has shown that the published and grey literature on RLP/PIS is dominated by the USA. While awareness of RLPs and their value is low amongst the public, there is consistent evidence of acceptability across countries, settings, tools, formats, users and providers. There was a great deal of consistency in the types of questions asked to help a person develop an RLP, with many RLPs based on the 2006 Moos RLP(7), corroborating what Edmunds et al’s concept analysis of RLPs found(8). However more recent tools have been developed to be more patient-centric, support reproductive autonomy and to be inclusive. There was also consistency in the topics covered, though those used outside a clinical encounter e.g. those in the grey literature, tended to be more holistic and less focused on reproductive health service needs, likely because they were not constrained by the time limits. However, their accessibility and longevity appear to be limited, with one-third (33%) of the grey literature RLPs extracted in 2023 no longer being available when reassessed in 2026. This high level of attrition highlights the transient nature of grey literature resources and raises concerns about their long-term sustainability, discoverability, and reliability as sources of patient information. Given the small number of digital RLPs, there is a scarcity of evidence on these tools, however, they are more likely to have underpinning theories and to have conducted more detailed effectiveness evaluations.

## Results In Context of What is Known

This review updates and extends the findings of previous reviews in the area. Hipp et al’s review of the peer-reviewed literature up to 2017 found that RLP approaches were generally seen as acceptable and clinically useful. Although they were well received in clinical practice there was limited evidence regarding their impact on knowledge, attitudes, or behavioural outcomes to demonstrate effectiveness(88). A narrative review published in 2022 considered the evidence for reproductive life planning in adolescents concluding that while it was acceptable in adult populations, there was minimal evidence regarding their potential benefits for adolescents(89). Despite this review being conducted nearly a decade later, we show that these evidence gaps remain.

Another 2022 review, focused on reproductive screening tools used in clinical settings in the United States, identified a range of approaches and formats for preconception, interconception, and pregnancy intention screening(90). We find ongoing diversity with, if anything, increasing numbers of tools that do not always build on previous learning or take robust approaches to development by integrating theory, following robust intervention development methods(91) or including end-users.

### Clinical Implications

The findings of this review suggest that asking about pregnancy intentions and supporting reproductive life planning is acceptable to both people of reproductive age and healthcare professionals across a range of settings. Despite relatively low levels of awareness among the public, there was consistent evidence that discussions about pregnancy intentions and use of RLPs were viewed positively when implemented. This supports the integration of pregnancy intention screening and reproductive life planning into routine healthcare encounters, particularly in primary care, sexual and reproductive health services, maternity care and other settings where reproductive health is discussed.

However, acceptability alone is unlikely to be sufficient to achieve meaningful **changes** in reproductive health outcomes. Implementation strategies should support person-centred, non-directive discussions that respect reproductive autonomy and reproductive justice principles. Healthcare professionals may require training and practical tools to facilitate these conversations, while implementation approaches should consider barriers such as time constraints, service accessibility and the broader social determinants that influence reproductive decision-making. The emergence of digital RLPs also offers opportunities to provide reproductive planning support beyond the clinical encounter and may help increase access to information and decision support for people who do not regularly access healthcare services.

### Research Implications

Despite finding 51 studies, there is limited data of effectiveness beyond changes in knowledge or recording of discussion of reproductive health in consultations. While important, these outcomes are a step away from the behaviour change outcomes on which we would wish to judge effectiveness, such as reduced unplanned pregnancies, increased use of folic acid before pregnancy or increased satisfaction with contraceptive decisions. Part of the gap is that many studies did not look at effectiveness in terms of behaviour change or other outcomes, and another part is due to the heterogeneity of outcomes measured. Even where effectiveness was considered the evidence is still limited and conflicting. There are several possible reasons for this. Firstly, very few tools have been developed with due consideration to relevant behaviour change theory. This omission may be why tools have shown limited impact beyond increased knowledge and discussion. Where behaviour change theory has been utilised, precursors, such as self-efficacy have been focused on, without perhaps considering the other steps in the theory of change required to lead to impact. Secondly, the choice of outcome measure could also be a factor, and considering patient-centred measures, such as satisfaction with contraception, rather than efficacy-based outcome measures such as LARC use which reflect health system targets, may be better aligned to the concept of patient-centred RLPs(92) and reproductive justice approaches. Thirdly there is potential selection bias in that those most likely to engage with an RLP might be the healthiest and least in need of change, resulting in a high baseline that is hard to affect. Fourthly, it is also important to consider that an individuals’ preferences and choices might be constrained by other factors, such as access to services and the (opportunity) cost of contraception which an RLP focusing on individual behaviour change may be unable to affect. Examination of the implementation context and facilitators and barriers in each setting and for each population group and tool is therefore also required, particularly in the context of worsening maternal mortality in globally. Finally, the digital RLPs developed may lack features such as personalisation, adaptive feedback and the ability to track behaviours which are known to facilitate engagement in mobile health apps(93–95).

Future research should move beyond assessing acceptability, knowledge gains and documentation of reproductive health discussions, and instead examine behavioural, reproductive and health outcomes. Greater consistency in outcome selection and reporting would facilitate comparison across studies and strengthen the evidence base. Research is also needed to better understand the implementation of RLPs across different settings and populations, including the barriers and facilitators that influence their use and effectiveness. Future intervention development should draw on established behaviour change theories and clearly articulated theories of change (e.g Normalisation Process Theory(96)), while digital RLPs should incorporate evidence-based engagement features such as personalisation and adaptive feedback. Given the absence of UK-based evidence identified in this review, further research is particularly needed to evaluate the acceptability, implementation and effectiveness of RLPs within UK healthcare settings to inform implementation.

### Strengths & Limitations

A key strength of this review is the inclusion of both published and grey literature, providing a comprehensive overview of current PIS and RLP approaches. Inclusion of grey literature enabled identification of patient-facing tools that may not be represented in academic publications, offering valuable insight into the resources available to people of reproductive age. Another strength is the breadth and depth of data extraction. In addition to describing the format and content of RLPs, we examined their acceptability, effectiveness and underlying behaviour change theory. This allowed a more comprehensive assessment of existing approaches and helped identify important gaps in intervention design and evaluation. Finally, the searches were updated in 2026, ensuring that this review reflects the most current evidence.

Due to our particular interest in digital tools, we focused on relatively recent literature. This could mean that we missed some RLPs, however given the consistency in the questions and topics covered in the identified RLPs this is unlikely to impact the overall findings of the review. We only searched in English, which would limit our ability to find studies or grey literature on RLPs in other languages. We excluded only one paper based on language (Hungarian) but did include an RLP in Swedish as the publications about it were in English.

Three papers did not include the questions that were asked as part of the RLP and only one author responded to our request for further information. Furthermore, some of the papers, particularly about the digital tools, only discussed the topics included within the RLPs. We were able to access some, but not all, meaning that we did not always have the specific wording of the questions used.

## Conclusions

The concept of developing an RLP is acceptable to people of reproductive age and health care professionals, but awareness is low and evidence of how to effectively implement PIS/RLP is inadequate, with none from the UK. New digital tools that are underpinned by behaviour change theories are emerging and though evidence of effectiveness is limited to date, it is promising. Digital RLPs that builds on the existing tools, integrate appropriate theory, learn from other digital health interventions, have an appropriate implementation strategy and that support person-centred care could facilitate reproductive choice and lead to the realisation of the potential that RLPs are theorised to deliver.

## Declaration of generative AI use

The authors report generative AI was not used in their research or preparation of this manuscript

## Data Availability Statement

The data supporting the findings of this scoping review are available within the article and its supplementary materials. All included sources are publicly available and are referenced in the manuscript.

